# Genomic architecture of Autism Spectrum Disorder from comprehensive whole-genome sequence annotation

**DOI:** 10.1101/2022.05.05.22274031

**Authors:** Brett Trost, Bhooma Thiruvahindrapuram, Ada J.S. Chan, Worrawat Engchuan, Edward J. Higginbotham, Jennifer L. Howe, Livia O. Loureiro, Miriam S. Reuter, Delnaz Roshandel, Joe Whitney, Mehdi Zarrei, Matthew Bookman, Cherith Somerville, Rulan Shaath, Mona Abdi, Elbay Aliyev, Rohan V. Patel, Thomas Nalpathamkalam, Giovanna Pellecchia, Omar Hamdan, Gaganjot Kaur, Zhuozhi Wang, Jeffrey R. MacDonald, John Wei, Wilson W.L. Sung, Sylvia Lamoureux, Ny Hoang, Thanuja Selvanayagam, Nicole Deflaux, Melissa Geng, Siavash Ghaffari, John Bates, Edwin J. Young, Qiliang Ding, Carole Shum, Lia D’abate, Clarissa A. Bradley, Annabel Rutherford, Vernie Aguda, Beverly Apresto, Nan Chen, Sachin Desai, Xiaoyan Du, Matthew L.Y. Fong, Sanjeev Pullenayegum, Kozue Samler, Ting Wang, Karen Ho, Tara Paton, Sergio L. Pereira, Jo-Anne Herbrick, Richard F. Wintle, Jonathan Fuerth, Juti Noppornpitak, Heather Ward, Patrick Magee, Ayman Al Baz, Usanthan Kajendirarajah, Sharvari Kapadia, Jim Vlasblom, Monica Valluri, Joseph Green, Vicki Seifer, Morgan Quirbach, Olivia Rennie, Elizabeth Kelley, Nina Masjedi, Catherine Lord, Michael J. Szego, Ma’n H. Zawati, Michael Lang, Lisa J. Strug, Christian R. Marshall, Gregory Costain, Kristina Calli, Alana Iaboni, Afiqah Yusuf, Patricia Ambrozewicz, Louise Gallagher, David G. Amaral, Jessica Brian, Mayada Elsabbagh, Stelios Georgiades, Daniel S. Messinger, Sally Ozonoff, Jonathan Sebat, Calvin Sjaarda, Isabel M. Smith, Peter Szatmari, Lonnie Zwaigenbaum, Azadeh Kushki, Thomas W. Frazier, Jacob A.S. Vorstman, Khalid A. Fakhro, Bridget A. Fernandez, M.E. Suzanne Lewis, Rosanna Weksberg, Marc Fiume, Ryan K.C. Yuen, Evdokia Anagnostou, Neal Sondheimer, David Glazer, Dean M. Hartley, Stephen W. Scherer

## Abstract

Fully understanding the genetic factors involved in Autism Spectrum Disorder (ASD) requires whole-genome sequencing (WGS), which theoretically allows the detection of all types of genetic variants. With the aim of generating an unprecedented resource for resolving the genomic architecture underlying ASD, we analyzed genome sequences and phenotypic data from 5,100 individuals with ASD and 6,212 additional parents and siblings (total n=11,312) in the Autism Speaks MSSNG Project, as well as additional individuals from other WGS cohorts. WGS data and autism phenotyping were based on high-quality short-read sequencing (>30x coverage) and clinically accepted diagnostic measures for ASD, respectively. For initial discovery of ASD-associated genes, we used exonic sequence-level variants from MSSNG as well as whole-exome sequencing-based ASD data from SPARK and the Autism Sequencing Consortium (>18,000 trios plus additional cases and controls), identifying 135 ASD-associated protein-coding genes with false discovery rate <10%. Combined with ASD-associated genes curated from the literature, this list was used to guide the interpretation of all other variant types in WGS data from MSSNG and the Simons Simplex Collection (SSC; n=9,205). We identified ASD-associated rare variants in 789/5,100 individuals with ASD from MSSNG (15%) and 421/2,419 from SSC (17%). Considering the genomic architecture, 57% of ASD-associated rare variants were nuclear sequence-level variants, 41% were nuclear structural variants (SVs) (mainly copy number variants, but also including inversions, large insertions, uniparental isodisomies, and tandem repeat expansions), and 2% were mitochondrial variants. Several of the ASD-associated SVs would have been difficult to detect without WGS, including an inversion disrupting *SCN2A* and a nuclear mitochondrial insertion impacting *SYNGAP1*. Polygenic risk scores did not differ between children with ASD in multiplex families versus simplex, and rare, damaging recessive events were significantly depleted in multiplex families, collectively suggesting that rare, dominant variation plays a predominant role in multiplex ASD. Our study provides a guidebook for exploring genotype-phenotype correlations in the 15-20% of ASD families who carry ASD-associated rare variants, as well as an entry point to the larger and more diverse studies that will be required to dissect the etiology in the >80% of the ASD population that remains idiopathic. All data resulting from this study are available to the medical genomics research community in an open but protected manner.

## Introduction

Autism Spectrum Disorder (ASD) is a neurodevelopmental condition whose core symptoms are social and communication difficulties, repetitive behaviors, and a restricted set of interests (American Psychiatric Association, 2013; Lord et al., 2020). In recent epidemiological studies, ASD has been observed in approximately 2% of individuals and is about four times more common in males than females (Maenner et al., 2020; Public Health Agency of Canada, 2021; Centers for Disease Control and Prevention, 2022). ASD is clinically heterogeneous, with some affected individuals exhibiting mild if any difficulties and others experiencing very distressing symptoms and a range of co-occurring conditions that affect both physical and mental health (Doshi-Velez et al., 2014). Twin studies have estimated the heritability of ASD to be 64-91% (Tick et al., 2016). Rare or *de novo* high-impact genetic variants, often with diagnostic value, are typically identified in <20% of individuals with ASD, often in those with more complex medical presentations (Tammimies et al., 2015; Fernandez and Scherer, 2017; Schaaf et al., 2020).

Previous ASD WGS studies, including our own, have examined the contribution of different classes of genetic variants (e.g., sequence-level, copy number, structural; see Supplementary Table 1 for a summary of major studies). However, these studies typically analyzed one or a small number of variant classes at a time and were thus unable to comprehensively delineate the genomic architecture of ASD.

In this study, we introduce a substantial update to the Autism Speaks MSSNG ASD whole-genome sequencing (WGS) resource (Figure 1). Compared with the previous release (Yuen et al., 2017), MSSNG now contains twice as many individuals with ASD (5,100 versus 2,626) and total individuals (11,312 versus 5,205, including parents and other family members; Table 1). Additionally, we describe a thorough annotation of the MSSNG genomic data, leveraging the power of WGS to explore the contribution to ASD susceptibility of variants that are inaccessible to genotyping methods such as chromosomal microarrays and whole-exome sequencing (Figure 1). We performed a comprehensive examination of many variant types in a single study, including common single nucleotide polymorphisms (SNPs), as well as rare and *de novo* single nucleotide variants (SNVs), short insertions/deletions (indels), mitochondrial DNA (mtDNA) variants, and structural variants (SVs, which include copy number variants (CNVs), inversions, larger insertions, uniparental disomies (UPDs), and tandem repeat expansions (TREs)). These analyses encompass both coding and non-coding regions of the genome and both dominant and recessive modes of inheritance. We also used high-coverage (>30x) WGS data of 9,205 individuals from the Simons Simplex Collection (SSC) (Fischbach and Lord, 2010) as a replication dataset and 2,504 unrelated population controls from the 1000 Genomes Project (1000G) (1000 Genomes Project Consortium et al., 2015), for a total of over 23,000 WGS samples analyzed (Supplementary Table 2). Variant detection and analysis of the SSC and 1000G data were performed using identical or near-identical pipelines as for MSSNG, allowing for true comparability across datasets. The current release of MSSNG also includes a completely redesigned online portal allowing the exploration of both genotype and phenotype data, integration with the Terra platform (https://terra.bio), and many other enhancements that facilitate clinical genomics research (Table 1).

**Figure 1:**
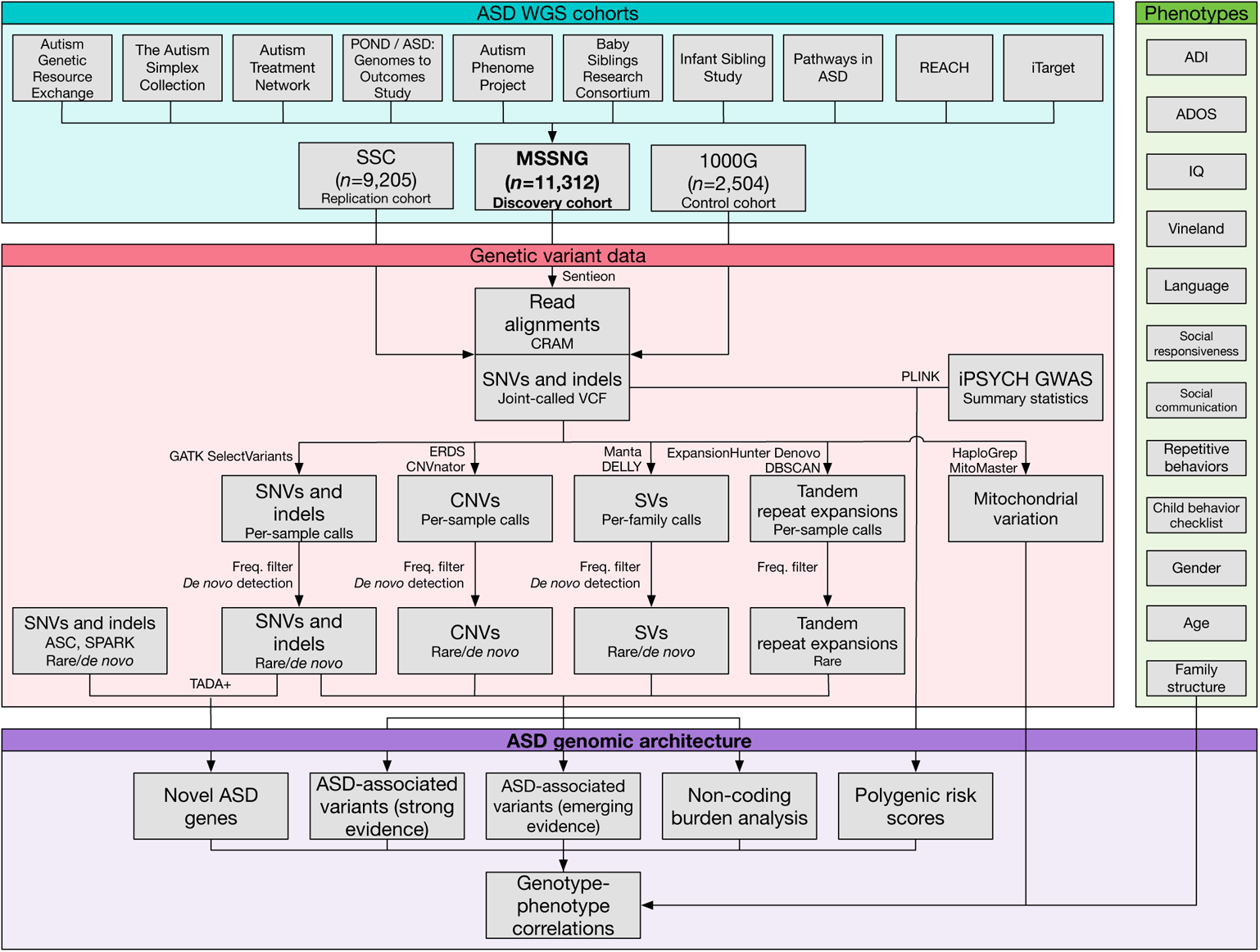
Data processing and analysis workflow used in this study. The “genetic variant data” section applies to all samples except n=1,738 samples from MSSNG sequenced using Complete Genomics technology, for which variants were detected using proprietary Complete Genomics software. Abbreviations: 1000G, 1000 Genomes Project; ADI, Autism Diagnostic Interview; ADOS, Autism Diagnostic Observation Schedule; ASC, Autism Sequencing Consortium; ASD, Autism Spectrum Disorder; CNV, copy number variant; GWAS, genome-wide association study; POND, Province of Ontario Neurodevelopmental Network; REACH, Relating genes to Adolescent and Child mental Health; SNV, single nucleotide variant; SPARK, Simons Foundation Powering Autism Research; SSC, Simons Simplex Collection; SV, structural variant; VCF, variant call format.

**Table 1:** Comparison between the previously-described version of the Autism Speaks MSSNG WGS resource (Yuen et al., 2017) and the current release. A few individuals were sequenced twice for quality-control purposes, which are reflected in the genome counts; the numbers of total individuals and individuals with ASD are 11,312 and 5,100, respectively.

|  | Yuen et al., 2017 | Current |
| --- | --- | --- |
| Participants |  |  |
| Total genomes | 5,205 | 11,359 |
| ASD genomes | 2,626 | 5,121 |
| Families | 2,066 | 4,258 |
| Genetic variant data |  |  |
| Reference genome | GRCh37/hg19 | GRCh38/hg38 |
| Single-nucleotide variants and indels | Single sample | Joint-called |
| Copy number variants | Not available | Available |
| Tandem repeat expansions | Not available | Available |
| Structural variants | Not available | Available |
| Polygenic risk scores | Not available | Available |
| Phenotype data |  |  |
| Composite phenotype measures | Not available | Available |
| Data access |  |  |
| Phenotype data explorer | Not available | Available |
| Terra integration | Not available | Available |
| Number of researchers approved for access |  |  |
| Researchers | 75 | 331 |
| Institutions | 17 | 67 |
| Countries | 4 | 20 |

## Results

### Overview of MSSNG

The Autism Speaks MSSNG resource (https://research.mss.ng) contains WGS data and phenotype information from individuals with ASD and their family members. We acknowledge that different perspectives exist within the ASD community regarding the preferred language to refer to individuals with ASD, or autistic people. In this manuscript, we respectfully use the first option, even though we recognize this may not be the terminology preferred by all (Vivanti, 2020; Bury et al., 2020; Botha et al., 2021). All individuals with ASD in MSSNG meet diagnostic criteria according to the Diagnostic and Statistical Manual of Mental Disorders (DSM) (American Psychiatric Association, 2013), also supported in many individuals by the Autism Diagnostic Interview-Revised (ADI-R) (Lord et al., 1994; Kim and Lord, 2012) and/or the Autism Diagnostic Observation Schedule (ADOS) (Lord et al., 1989, 2000). MSSNG aggregates data from several cohorts and studies (Supplementary Table 3); the largest contributor is the ASD: Genomes to Outcomes Study (Prasad et al., 2012), for which a major portion of participants are from the Province of Ontario Neurodevelopmental Network (POND; https://pond-network.ca/).

The latest release of MSSNG contains 11,359 samples from 11,312 unique individuals (47 samples were re-sequenced for quality control purposes), including 5,100 individuals with ASD (Supplementary Table 3). These include 4,073 affected males and 1,027 affected females, giving a male/female ratio of 4:1. Approximately 25% of the individuals in MSSNG are of non-European ancestry, including >2% each Admixed American, African, East Asian, and South Asian (Supplementary Figure 1). For 3,565 of the individuals with ASD, genome sequences for both parents are available. MSSNG contains many multiplex families, including 696 having two individuals with ASD and 88 with three or more. MSSNG also includes 263 unaffected siblings (Supplementary Table 3). Early MSSNG samples (n=1,738) were sequenced using Complete Genomics technology (Drmanac et al., 2010), while subsequent samples (n=9,621) were sequenced on Illumina platforms, mainly on the HiSeq X instrument (Supplementary Table 3). With few exceptions, all members of a given family were sequenced on the same platform.

Extensive phenotype data based on 121 different tests are available in MSSNG, with the exact data available for a given individual depending on the specific tests applied. These include ADI/ADOS diagnostic classification, co-morbidities, and IQ measures, as well as scores for the Vineland Adaptive Behavior Scale, Repetitive Behavior Scale, Social Responsiveness Scale, Social Communication Questionnaire, Child Behavior Checklist, and Aberrant Behavior Checklist (Supplementary Table 4). The MSSNG phenotype data also includes features related to syndromic autism, such as height, micro/macrocephaly, and dysmorphologies.

The organizational structure of MSSNG has been described previously (Yuen et al., 2017). Briefly, usage is controlled by a Data Access Committee, and qualified scientists can apply for access by submitting a proposal describing an ASD-related research question. As of January 2022, 331 researchers from 67 institutions in 20 countries have been approved for access, and at least 126 publications have used or acknowledged MSSNG resources (Supplementary Table 5). All MSSNG data are stored on the Google Cloud Platform (GCP); large flat files (e.g., FASTQ and CRAM files) are accessible via Cloud Storage buckets, while variant calls, variant annotations, sample metadata, and phenotype information are stored as BigQuery tables. The MSSNG web portal (https://research.mss.ng) allows variants to be queried based on sample, gene, or genomic region.

### Enhancements to MSSNG and online resources

Besides its substantial increase in sample size, the latest release of MSSNG contains numerous additional improvements (Table 1). Sequence reads are aligned to the latest version of the human reference genome (GRCh38/hg38), and SNVs and indels from samples sequenced on Illumina platforms are joint-called for improved accuracy. Also available are several additional types of variant calls, all of which are accessible via both the MSSNG portal and BigQuery. These include CNVs (deletions and duplications >=1 kb detected using a previously-described read depth-based pipeline (Trost et al., 2018)); SVs (deletions, duplications, insertions, and inversions >=50 bp identified using a novel pipeline described in this paper); tandem repeats (detected as part of a previous study (Trost et al., 2020)), and polygenic risk scores (PRS) (calculated as described in this study).

Numerous enhancements have been made to the MSSNG web portal. The interface for querying variants has been redesigned to provide a more intuitive user experience and to accommodate the additional variant types that can now be queried (for example, CNV queries; Supplementary Figure 2). The Read Viewer, implemented using an Integrative Genomics Viewer (IGV) plugin (Robinson et al., 2011), includes tracks showing CNV and SV calls for all members of the family being examined. The portal now features a Phenotype Data Explorer allowing users to analyze phenotypic data at the level of specific individuals, subsets of the data (for example, stratified by sex), or the entire dataset (Supplementary Figure 3). For users requiring specific or customized data, the Phenotype Data Explorer also allows structured query language (SQL) queries to be run within the platform itself.

Because the number of measures is large, exploring the phenotypic data can be challenging for users who lack extensive knowledge of the various tests. This difficulty is compounded by the fact that the tests (or versions thereof) used in each cohort comprising MSSNG vary, and because the tests have evolved over time. To address these challenges, we have developed several composite phenotype measures, each of which encapsulates several phenotypic data points into one easy-to-understand measure. For example, the adaptive behaviour standard score uses the most recent of four possible scores for adaptive behavior. A more complex composite measure is the global ability composite estimate, which is calculated based on several measures related to IQ, general verbal and nonverbal ability, and motor and language skills. Other composite measures include the socialization standard score, full-scale IQ, and measures for common co-occurring conditions (attention deficit hyperactivity disorder (ADHD), anxiety, gastrointestinal problems, and epilepsy).

### Discovery of ASD-associated genes using TADA+

Previously, the Autism Sequencing Consortium (ASC) developed TADA+, an enhanced version of the transmission and *de novo* association (TADA) test (He et al., 2013) that adds information about genic constraint (pLI) and predicted missense deleteriousness (MPC score) (Samocha et al., 2017). The TADA+ model was applied to whole-exome sequencing (WES) data from 6,430 trios, 5,556 cases, and 8,809 controls, identifying 102 ASD-associated genes with false discovery rate (FDR) <0.1 (Satterstrom et al., 2020). To increase power for discovering ASD-associated genes, here we incorporated *de novo* variants from 12,375 additional trios from the MSSNG (WGS) and Simons Foundation Powering Autism Research (SPARK) (WES) (SPARK Consortium, 2018; Feliciano et al., 2019) cohorts (Supplementary Tables 6-7). The distributions of the number of exonic *de novo* variants per child were similar for ASC, MSSNG, and SPARK (Figure 2 (A)). After incorporating the additional trios, the TADA+ model gave 135 ASD-associated genes with FDR <0.1 (Supplementary Table 8). Of these, 67 were identified in the previous TADA+ analysis (Satterstrom et al., 2020), 68 were new, and 35 genes from the previous analysis no longer met the FDR threshold (Figure 2 (B)). Many of the new genes (27/68; 40%) are not currently in the SFARI Gene database (Banerjee-Basu and Packer, 2010), providing a rich source of new ASD-associated genes for hypothesis generation and functional studies (Figure 2 (C)). All 30 genes with the lowest FDR from the previous study were retained, including well-established ASD-linked genes such as *CHD8*, *SYNGAP1*, *SCN2A*, and *SHANK3* (Figure 2 (D)).

**Figure 2:**
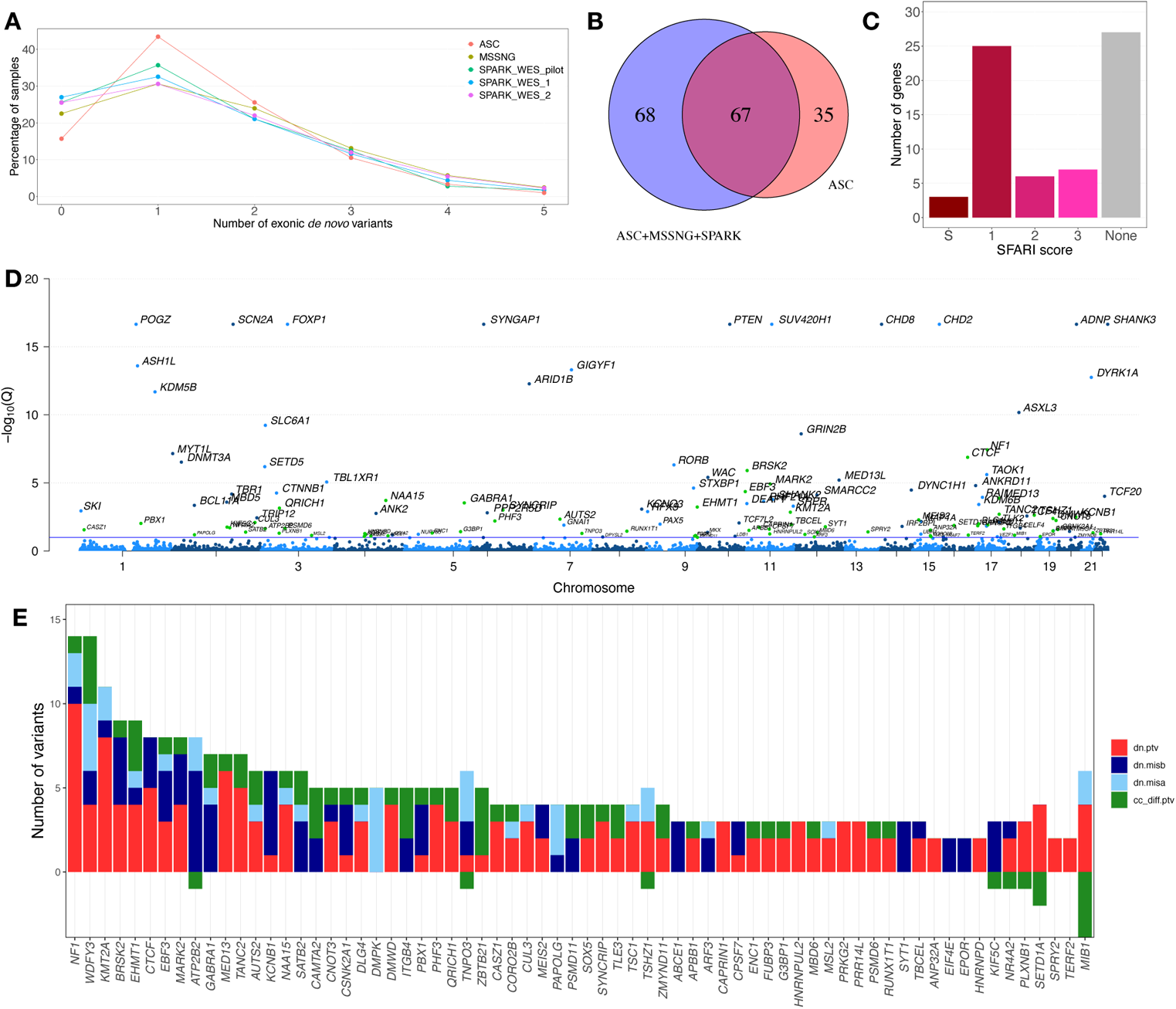
Identification of ASD-associated genes using TADA+. (A) Distribution of the number of exonic *de novo* variants per individual in the ASC, MSSNG, and SPARK datasets. (B) Number of ASD-associated genes discovered only in the previous TADA+ analysis (“ASC”) (Satterstrom et al., 2020), only in the current analysis (“ASC+MSSNG+SPARK”), or both. (C) Distribution of SFARI Gene scores for the 68 genes not identified in the previous TADA+ analysis. (D) Manhattan plot showing the false discovery rates of the 135 ASD-associated genes. Light or dark blue dots represent genes also identified in the previous TADA+ analysis, while green dots represent newly discovered ASD-associated genes. For visualization purposes, genes with FDR calculated to be zero were assigned a ceiling value of 10^!,%^. The blue horizontal line represents FDR=0.1. (E) Evidence supporting the 68 ASD-associated genes not identified in the previous TADA+ analysis. dn.ptv, *de novo* protein-truncating variants; dn.misb, *de novo* missense variants predicted to be highly damaging (MPC score ≥ 2); dn.misa, *de novo* missense variants predicted to be moderately damaging (1 ≤ MPC score < 2); cc_diff.ptv, number of protein-truncating variants in cases minus the number in controls (with negative values indicating more variants in controls than cases). The y-axis is truncated at −3; the cc_diff.ptv value for *MIB1* is −9.

For most of the new ASD-associated genes, the evidence constituted a mix of *de novo* protein-truncating variants (PTVs), *de novo* missense variants, and excess PTVs in cases compared with controls (Figure 2 (E)). However, for some genes the evidence consisted exclusively of PTVs (e.g., *MED13*, *TANC2*, *DMWD*, *CAPRIN1*) or exclusively of *de novo* missense variants (e.g., *ATP2B2*, *DMPK*, *PAPOLG*, *ABCE1*). These biases may provide insight into molecular mechanisms—for example, haploinsufficiency for PTV-biased genes and gain-of-function or dominant-negative mechanisms for missense-biased genes. Most of the newly identified genes had high pLI scores (Supplementary Figure 4), suggesting a general bias toward haploinsufficiency as molecular mechanism.

One gene identified by our updated TADA+ analysis that was of particular interest was *DMPK*. RNA toxicity stemming from TREs in the 3′ untranslated region of *DMPK* cause myotonic dystrophy type I (DM1) (Sznajder and Swanson, 2019), and individuals with DM1 have a higher incidence of ASD (Ekström et al., 2008; Lagrue et al., 2019). In addition, we recently identified *DMPK* as one of the top candidate genes for association of TREs with ASD (Trost et al., 2020). Here, five *de novo* missense variants were detected in *DMPK* (Supplementary Table 8), suggesting that missense variants may represent another mechanism by which this gene contributes to ASD susceptibility. None of the five individuals were from MSSNG, so we were unable to determine if they had features of DM1.

Another interesting novel ASD-candidate gene was *GABRA1*, for which we detected two PTVs in ASD cases and five *de novo* missense variants (Supplementary Table 8). *GABRA1* encodes the most abundant α subunit of the GABA_A_ receptor, which mediates fast inhibitory neurotransmission (Zhu et al., 2018). Synaptic protein dysregulation and other alterations in the GABAergic system, including in the GABA_A_ receptors, have previously been implicated in ASD (Fatemi et al., 2009; Coghlan et al., 2012; Di et al., 2020). Mouse models also support an association between ASD and *GABRA1*: mice treated with valproic acid, one of the few suspected environmental susceptibility factors for ASD (Christensen et al., 2013), exhibit ASD-like phenotypes and significantly decreased *GABRA1* expression (Chau et al., 2017). *GABRA1* is also a target for potential therapeutics, with several programs exploring potential novel medications both in ASD and related rare disorders.

Our analysis uncovered several new ASD-associated genes located within CNV regions known to be associated with both ASD and neurodevelopmental disorders more generally. For example, *PRKG2*, *HNRNPD*, and *HNRDPL* are three of the five genes located within the critical region for 4q21 microdeletion syndrome, which is associated with growth restriction, intellectual disability, and absent or delayed speech (Bonnet et al., 2010; Hu et al., 2017). Another example is *TSHZ1*, which has been most strongly associated with a non-NDD phenotype (congenital aural atresia), but is also found within the 18q deletion region, which is characterized by intellectual disability, various dysmorphologies, and increased susceptibility for ASD (O’Donnell et al., 2010). Other TADA+ genes within ASD-associated CNV regions included *CASZ1* (1p36), *TBCEL* (11q23.3), *PSMD11* (17q11.2), *RUNX1T1* (8q21.3), *ABCE1* (4q31.21-q31.22), and *PAPOLG* (2p16.1-p15). The identification of these genes may provide insight into which gene(s) within these CNV regions contribute to their associated neurodevelopmental phenotypes.

One of the highest-confidence genes identified in the original TADA+ analysis to “drop out” was *NRXN1* (previous FDR=8.1 × 10^−4^). In the ASC data, the evidence for *NRXN1* was three *de novo* PTVs, one *de novo* missense variant, and one PTV in a case (versus none in controls). However, no *de novo* PTVs or missense variants were found in *NRXN1* in MSSNG or SPARK, resulting in a new FDR (0.19) that did not meet the cutoff. This does not mean that *NRXN1* is irrelevant to ASD; rather, it reflects the limitation that TADA+ considers only sequence-level variants. Previous evidence for the involvement of *NRXN1* in ASD and other neurodevelopmental disorders has largely been derived from deletions (Autism Genome Project Consortium et al., 2007; Lowther et al., 2017). *NRXN1* was also identified in our recent study on TREs in ASD (Trost et al., 2020), underscoring the idea that the totality of evidence associated with a given gene must be considered, including that from different types of genetic variants. Indeed, according to the EAGLE ASD-gene curation protocol (Schaaf et al., 2020), *NRXN1* is a definitive ASD-linked gene.

Of the 135 TADA+ genes, 107 (79%) were involved in gene networks. Testing for biological process and pathway enrichment, we stratified genes into several groups, including transcription regulation, synaptic processes, RNA processing, signalling pathways, epigenetic gene regulation, and proteasome components (Figure 3). A substantial portion of the ASD-associated genes in the RNA processing (14/21; 67%) and epigenetic gene regulation (6/8; 75%) categories were newly identified, suggesting that they may be promising candidates for new biological processes involved in ASD etiology.

**Figure 3:**
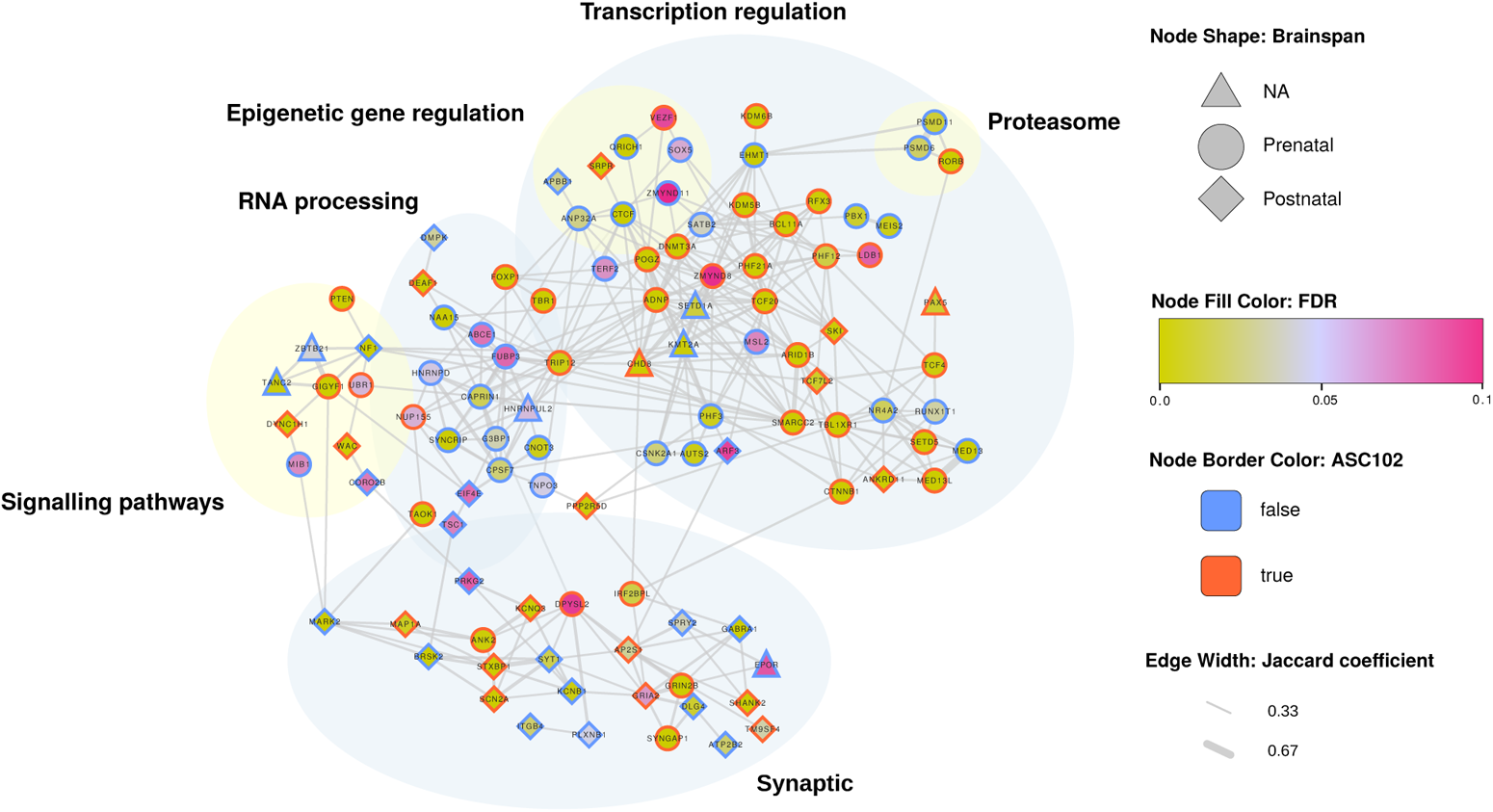
Network diagram of TADA+ genes. Only genes connected to gene networks are shown. Nodes represent genes, and edges represent physical (protein-protein interaction) and pathway interactions between gene pairs. Node shape represents BrainSpan pre/postnatal gene expression (Kang et al., 2011; Miller et al., 2014), and node fill color represents TADA+ FDR (see legend). Red node borders represent genes also identified in the previous ASD TADA+ analysis, while blue borders represent newly identified ASD-associated genes.

Detecting new ASD-associated genes also enhances our ability to identify potential links with other distinct or phenotypically overlapping disorders. While the overlaps between ASD and conditions such as ADHD, obsessive-compulsive disorder (OCD), intellectual disability (ID), and epilepsy have been well-established (Viscidi et al., 2013; Kushki et al., 2019; Thurm et al., 2019), potential links between ASD and other disorders are less clear. Interestingly, several of our novel ASD-associated genes have tentative links with neurodegenerative disorders, such as Alzheimer’s disease (AD)—*NR4A*2 (Jesús et al., 2021; Moon et al., 2019), *ATP2B2* (Brendel et al., 2014), *EIF4E* (Li et al., 2004; Ghosh et al., 2020), *EBF3* (Rao et al., 2018), *MARK2* (Segu et al., 2008; Gu et al., 2013; Zhou et al., 2019), *EPOR* (Brettschneider et al., 2006; Hernández et al., 2017), *G3BP1* (Martin et al., 2013; Silva et al., 2019), *PLXNB1* (Yu et al., 2018; Zhao et al., 2021), *APBB1* (McLoughlin and Miller, 2008; Zeidán-Chuliá et al., 2014; Probst et al., 2020), and *ANP32A* (Chai et al., 2017, 2018; Podvin et al., 2020). The link between ASD and neurodegenerative disease may also extend to X-linked genes; for example, variants in *RAB39B* have been linked to both (Mata et al., 2015; Woodbury-Smith et al., 2017). Although highly distinct both phenotypically and genetically, ASD and neurodegenerative diseases have common clinical features, including impairments of language, executive function, and motor skills (Khan et al., 2016); they also share aspects of their molecular pathomechanism (e.g., synaptic deregulation), with several genes having been proposed to contribute to both (Jęśko et al., 2020). Further evidence for genetic links includes early suggestions of an increased likelihood that individuals with ASD will later be diagnosed with AD-like disorders (Plana-Ripoll et al., 2019) and increased incidence of dementia (Vivanti et al., 2021). However, a major challenge in evaluating these associations is the paucity of longitudinal studies in adults with autism (Lord et al., 2022).

Finally, we enumerated the X-linked genes with the greatest number of hemizygous PTVs in males with ASD from MSSNG, SSC, and SPARK (Supplementary Table 9). The most well-represented gene was *DMD*, which has previously been linked to ASD in addition to its association with Duchenne muscular dystrophy (Wu et al., 2005). We also detected multiple hemizygous PTVs in established X-linked ASD genes, including those of the neuroligin family (*NLGN3*, *NLGN4X*).

### Identification of recessive genes and variants

To identify recessive genes and variants, we searched for homozygous and compound heterozygous PTVs and damaging missense (DMis) variants. A total of 151 genes harbored at least one biallelic PTV event or at least two biallelic DMis events, and 50 had damaging biallelic events in at least three unrelated individuals with ASD. The gnomAD pRec score, which represents the probability of intolerance to biallelic loss of function, was higher in these genes than other genes (Mann-Whitney test: p=0.0004). Five of these genes (*SYNE1, FAT1, VPS13B, DNAH3,* and *ITGB4*) have previously been associated with ASD (Krumm et al., 2015; Wang et al., 2020). *ITGB4* was also identified in the TADA+ analysis, suggesting that it may confer ASD susceptibility in a dose-dependent manner.

The two genes with the largest number of damaging biallelic events were *SYNE1* and *NEB*. *SYNE1* had one biallelic PTV and nine biallelic DMis events (two homozygous and eight compound heterozygous), while *NEB* had nine biallelic DMis events (one homozygous and eight compound heterozygous). Rare recessive variants in *SYNE1* have previously been described in individuals with ASD and ID (Yu et al., 2013; Riazuddin et al., 2017; Tuncay et al., 2022), whereas damaging biallelic variants in *NEB* have been reported to cause nemaline myopathy-2 (OMIM:256030), as well as being observed in patients exhibiting ID, epilepsy, and other conditions (Al-Mubarak et al., 2017).

In 25 multiplex families from MSSNG, two siblings with ASD shared the same rare biallelic deleterious event. These comprised a total of 26 events (9 homozygous and 17 compound heterozygous) in 25 distinct genes (one family had two events in the same gene), including *ACACB, ADGRV1, LRBA, NOL10, SORBS1,* and *PEPD*. In one multiplex family, the two individuals with ASD shared a compound heterozygous genotype in *LRIG1* (p.T152R and p.D1001N). *LRIG1* encodes the transmembrane protein “leucine rich repeats and immunoglobulin like domains 1” (Lrig1), which plays a role in receptor tyrosine kinase signaling and is required for proper nervous system development and plasticity (Alsina et al., 2012). One study showed that Lrig1-deficient mice exhibit hippocampal dendritic abnormalities that correlate with deficits in social behaviours similar to ASD phenotypes (Alsina et al., 2016).

A total of 26 individuals with ASD had damaging biallelic events (6 homozygous and 20 compound heterozygous) of established medical importance in ClinVar (pathogenic or likely pathogenic). These were found in 23 genes, including *ABCA3*, *OTUD6B*, *GAA*, *EIF3F*, *ATP7B*, *MYO7A*, and *CAD*, all of which are associated with known neurodevelopmental phenotypes such as ID, as well as metabolic diseases (Wilson disease and glycogen storage disease), deafness, and other syndromic conditions.

### Mitochondrial sequence variation and haplogroups

We evaluated pathogenic variants, haplogroups, and heteroplasmy (variants present in only some copies of an individual’s mtDNA genome) within mtDNA to study their association with ASD. We identified 23 instances of known pathogenic variants with >5% heteroplasmy in individuals with ASD from MSSNG and SSC (Supplementary Table 10). Of these, 17 were *de novo*, defined as when maternal heteroplasmy was either undetectable or <5%. The highest *de novo* heteroplasmy value was a 46.7% load of the m.13513G>A variant associated with Leigh Disease. The frequency of newly observed pathogenic heteroplasmies >5% in individuals with ASD (17/6,044) was significantly greater than the frequency of pathogenic heteroplasmies in mothers only (0/5,320) and fathers only (4/5,295) (χ^2^ test: p=6 × 10^!#^). Two pathogenic heteroplasmies were identified in individuals without maternal sequencing data, so inheritance status could not be determined. We also identified four families for which a pathogenic variant was present at >5% heteroplasmy in both a mother and her child with ASD. In one of these families, the child had a 48.8% load of the m.3243A>G variant associated with mitochondrial encephalopathy, lactic acidosis and stroke-like episodes (MELAS), which was increased from the maternal load of 12.2%. Both mother and child had clinical symptoms consistent with MELAS. The average intergenerational change in heteroplasmy for pathogenic variants was an 11.1% shift toward the pathogenic allele in children with ASD. We also evaluated mtDNA variants causing homoplasmic disorders generally affecting vision and hearing only, but we found no association with ASD (χ^2^ test: vision p=0.99; hearing p=0.40) (Supplementary Table 11).

It was previously reported that certain macrohaplogroups, such as I, J, K, T, A and M, were associated with higher ASD susceptibility (Chalkia et al., 2017). We retested this association by comparing proband haplogroups with those of their fathers (n=4,821 proband-father duos), which had the advantage that each case-control pair was identical at half of the nuclear loci but had no mitochondrial relationship. Contrary to previous findings, we observed no significant difference in haplogroup distribution between cases and controls (corrected p>0.05 for all haplogroups; Supplementary Table 12).

Next, we evaluated the inheritance of heteroplasmy at all mtDNA positions. A prior study suggested increased transmission of heteroplasmy to children with ASD compared with control siblings (Wang et al., 2016). However, we found no significant difference between the shifts from mother to affected child and the shifts from mother to unaffected sibling in either MSSNG (affected: 0.0026 ± 0.046; unaffected: 0.0043 ± 0.058; p=0.18) or SSC (affected: 0.0012 ± 0.042; unaffected: 0.0014 ± 0.048; p=0.65) (Supplementary Figure 5). This is inconsistent with the hypothesis that heteroplasmy is generally increased in children with ASD compared with unaffected siblings.

### Identification of structural variants

We used two pipelines to detect SVs: a previously-described read depth-based workflow (Trost et al., 2018), which detects CNVs >=1 kb, and a newly-developed workflow employing split read and paired-end mapping-based algorithms (Methods), which detects deletions, duplications, insertions, and inversions >=50 bp. We then identified rare (<0.1% population frequency) SVs falling into five categories: chromosomal abnormalities (e.g., aneuploidies); CNVs overlapping genomic disorder regions; large and gene-rich CNVs not overlapping known genomic disorder regions; uniparental disomies (UPDs); and smaller SVs disrupting genes implicated in ASD and other neurodevelopment disorders (including the 135 TADA+ genes). SVs were interpreted according to American College of Medical Genetics (ACMG) and Clinical Genome Resource (ClinGen) guidelines (Richards et al., 2015; Riggs et al., 2020). All SVs falling under the categories of “likely pathogenic” or “pathogenic” (LP/P) are described here as pathogenic. All pathogenic SVs were verified by BAM confirmation (Trost et al., 2018), overlap with microarray findings, and/or Sanger sequencing. For *de novo* SVs, we also verified the variant’s absence in the parental genomes.

The majority of the pathogenic SVs (96%) were deletions or duplications >=1 kb detected by our read depth pipeline; we identified comparatively few pathogenic deletions or duplications <1 kb, large insertions (that would not be detected by standard small-variant pipelines), and inversions. Pathogenic SVs were detected in 6% of individuals with ASD. Maternally inherited pathogenic SVs (n=93) were significantly more frequent than paternally inherited (n=68) (one-sided binomial test: p=0.03), possibly reflecting a female protective effect (Robinson et al., 2013).

UPD occurs when meiotic errors result in a child inheriting genetic material for a particular chromosome from only one parent. Isodisomic UPD (isoUPD) results in runs of homozygosity spanning some (partial isoUPD) or all (complete isoUPD) of a chromosome, potentially unmasking recessive variants. We identified two partial and three complete isoUPDs in individuals with ASD in MSSNG and SSC, with no isoUPDs detected in unaffected siblings. None were on chromosomes associated with imprinted disorders/domains (6, 7, 11, 14 and 15). It is difficult to determine whether this UPD rate represents an enrichment compared with the general population due to the lack of platform-matched data; however, non-significant enrichment was observed when comparing to a previous population study based on 23andMe data (Nakka et al., 2019) (ASD: 5/15,038 child-parent duos [0.033%]; general population: 129/916,712 duos [0.014%]; Fisher’s Exact Test: odds ratio (OR)=2.4, p=0.07). No heterodisomies were detected.

To date, few pathogenic inversions and large insertions have been found in ASD, possibly because their detection typically requires WGS. Here, we highlight two examples of such variants. The first is a 71 bp *de novo* frameshift insertion in *SYNGAP1* (Figure 4 (A)), which affects all known splice variants of the gene. A BLAT (Kent, 2002) search of the inserted sequence revealed that it comprised 65 bp of mtDNA from *MT-CO3* (cytochrome C oxidase III) plus a 6 bp microduplication (Figure 4 (B,C)). Although pathogenic mtDNA insertions have been described previously (e.g., (Borensztajn et al., 2002; Turner et al., 2003; Goldin et al., 2004; Millar et al., 2010), to our knowledge this is the first reported in ASD. We also detected a likely pathogenic 13,472 bp inversion of unknown inheritance (parental samples unavailable) affecting *SCN2A* (chr2:g.[165,376,995-165,390,457inv;165,390,458_165,390,464delTTTTAGAinsGT]). The presence of the inversion was supported both by Sanger sequencing, which confirmed that the 5’ end in the reference was at the 3’ end in this individual and vice-versa (Figure 4 (D)), and by IGV visualization, which confirmed the presence of anomalously mapped read pairs consistent with the signature for inversions (Figure 4 (E)). These examples serve to illustrate the ability of WGS to detect pathogenic variants that are difficult or impossible to ascertain using other technologies.

**Figure 4:**
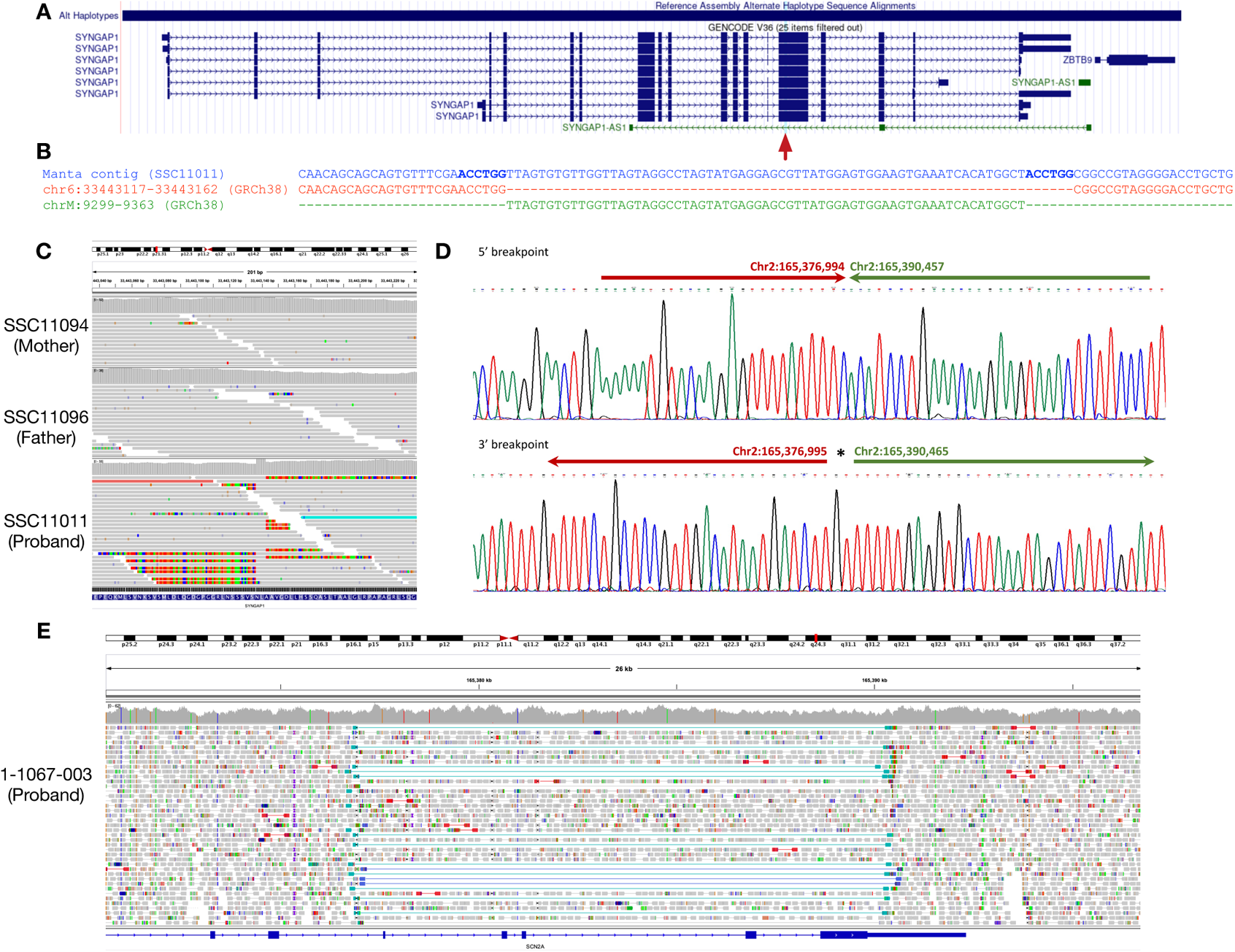
Examples of pathogenic structural variants detected in individuals with ASD in MSSNG and SSC. (A-C), 71 bp *de novo* frameshift insertion in *SYNGAP1* comprising 65 bp of mitochondrial sequence and a 6 bp microduplication in SSC individual SSC11011. (A) Schematic of the *SYNGAP1* gene, with the location of the insertion indicated by the red arrow. (B) Multiple sequence alignment of the insertion-containing contig sequence assembled by Manta, the relevant portion of the GRCh38 chromosome 6 reference sequence, and the portion of the mitochondrial reference sequence inserted into chromosome 6 in this individual. The 6 bp microduplication is indicated in bold. (C) Integrative Genomics Viewer visualization of the insertion. The colored portions of the reads represent mismatched bases, allowing the precise breakpoint to be determined. The increase in read depth reflects the 6 bp microduplication. No evidence for the insertion is present in either parent, confirming its *de novo* status. (D-E), 13.4 kb inversion overlapping *SCN2A* in MSSNG individual 1-1067-003. (D) Sequence trace showing the 5’ and 3’ breakpoints of the inversion. (E) Integrative Genomics Viewer visualization of the inversion, showing its relationship to *SCN2A*. The dark and light blue lines indicate anomalously mapped read pairs exhibiting the signature for an inversion.

Our SV data also afforded the opportunity to re-evaluate previous hypotheses concerning particular ASD-linked genes and variants. For example, deletions of exon 2 of the X-linked gene *TMLHE* have previously been proposed as a low-penetrance contributor to ASD susceptibility in males (Celestino-Soper et al., 2012). *TMLHE* codes for trimethyllysine hydroxylase epsilon, an enzyme comprising part of the pathway for the biosynthesis of the cellular energy production metabolite carnitine. To re-evaluate its association with ASD, we identified males from our three WGS datasets with deletions spanning exon 2 of *TMLHE* (Supplementary Figure 6; Supplementary Tables 13-14). Consistent with the previous report (Celestino-Soper et al., 2012), we detected an enrichment of *TMLHE* exon 2 deletions in male-male sibling pairs from multiplex ASD families compared with male controls (unaffected siblings, 1000G, and unaffected fathers in MSSNG and SSC), although in our study this difference was not significant (Fisher’s Exact Test: OR=2.5, p=0.22). Unlike the original study, we also detected a near-significant overall enrichment of such deletions in cases compared with controls (OR=1.8, p=0.12). While still tentative, these data suggest that further study is warranted into *TMLHE* as a low-penetrance susceptibility factor for ASD in males.

To harness the power of WGS to aid the interpretation of copy number duplications, we attempted to resolve the breakpoint junctions of duplications impacting a broad list of ASD candidate genes (Supplementary Table 15) in a subset of individuals with ASD in MSSNG. Because this process is currently manual in nature, we did not attempt to be exhaustive or to choose only duplications that would be definitively pathogenic if the overlapping gene was disrupted. Rather, we identified duplications impacting ASD candidate genes and show how their impact on gene structure and function can be evaluated by visualizing reads in IGV (Methods). By fine-mapping the breakpoints of 375 duplications identified by read depth-based algorithms in 332 individuals, we identified 248 unique duplications ranging in size from 4,254 bp to 6,522 kbp (Supplementary Table 16). We found that most duplications remained at or near their locus of origin, including tandem duplications (192/248; 77.4%), likely non-allelic homologous recombination (NAHR)-mediated events (6/248; 2.4%), and complex duplications involving sequence transposition <100 kb from their locus of origin or inverted triplications without sequence transposition (14/248; 5.6%). A subset of complex duplications involved large-scale sequence transpositions >100 kb (14/248; 5.6%) or interchromosomal transpositions (2/248; 0.8%). The structures of a minority of duplications (20/258; 8.1%) remained unresolved using short-read sequencing data. By examining read depth changes in IGV, we confirmed gene dosage increases of multiple ASD candidate genes. Several duplications of uncertain clinical significance increased gene dosage of established neurodevelopmental risk genes, including *de novo* tandem duplications encompassing *USP7* (OMIM: 602519) and *MEF2C* (OMIM: 600662). Both genes have emerging evidence for triplosensitivity and warrant further study to determine their clinical significance (Sanders et al., 2011; Novara et al., 2013; Uehara et al., 2016).

### Genomic architecture of rare coding variants in ASD

In this section, we synthesize the data presented above to give both a high-level view (burden analysis) and a detailed view (enumeration of specific genes and regions) of the genomic architecture of rare coding variants in ASD. We begin by comparing the burden of different variant types in individuals with ASD from either MSSNG or SSC with unaffected siblings from SSC. *De novo* PTVs in constrained genes (gnomAD observed/expected (o/e) <0.35) were significantly enriched in individuals with ASD (logistic regression: MSSNG OR=3.4, FDR=3 × 10^-6^; SSC OR=4.4, FDR=1.6 × 10^-7^), as were *de novo* damaging missense variants (MSSNG OR=1.7, FDR=0.001; SSC OR=1.8, FDR=0.0004) (Figure 5 (A)). We observed no significant enrichment in inherited PTVs in constrained genes. Inherited PTVs were enriched in the 135 TADA+ genes, although this trend was significant only for individuals with ASD in MSSNG (OR=1.6, FDR=0.03). No enrichment of inherited damaging missense variants was observed. Damaging recessive events were significantly enriched in individuals with ASD, and this enrichment was elevated when limited to genes with gnomAD pRec >0.9 (Figure 5 (A)). Substantial and significant enrichment was observed for nearly all categories of pathogenic SVs, including chromosomal abnormalities, genomic disorders, large and gene-rich CNVs, and SVs impacting ASD-linked genes (Supplementary Table 17). Finally, we previously reported that individuals with ASD have a higher burden of TREs in genic regions than unaffected siblings (Trost et al., 2020).

**Figure 5:**
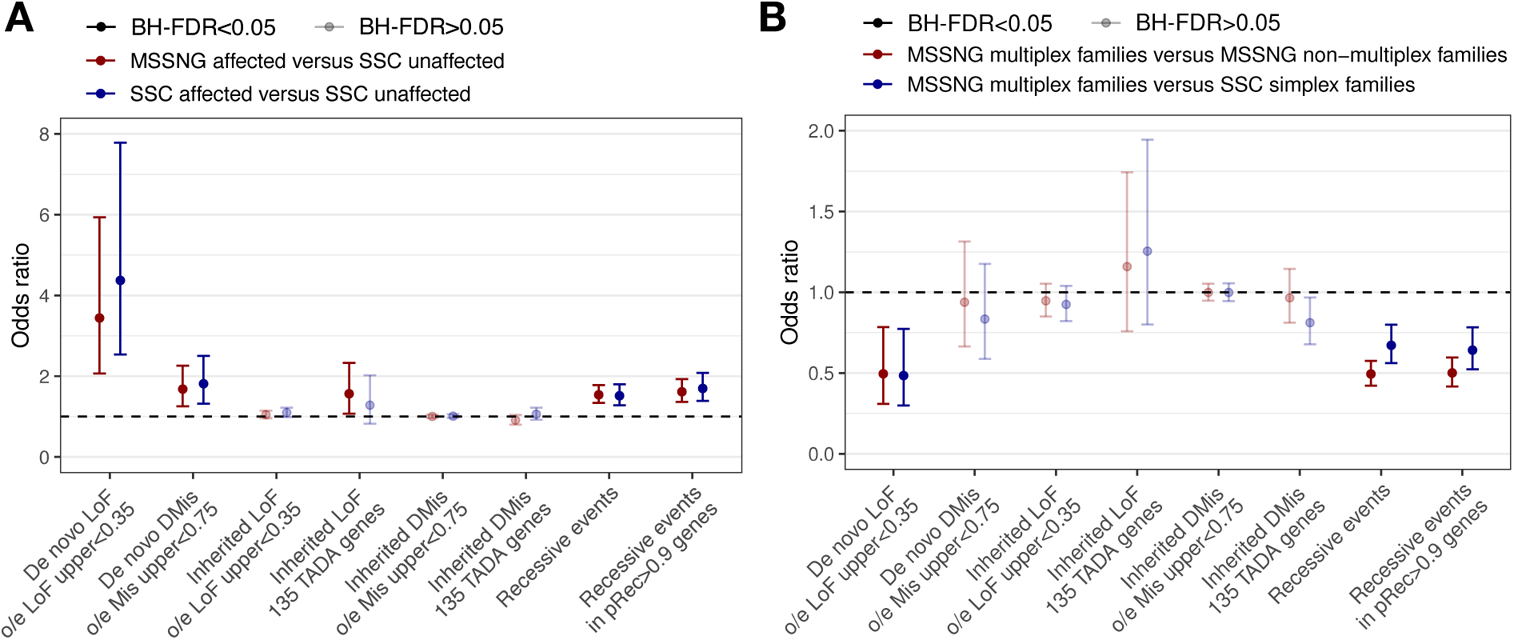
Burden analysis of sequence-level rare coding variants. (A) Individuals with ASD versus siblings without ASD. (B) Individuals with ASD from multiplex families from MSSNG versus those from either simplex families from SSC (defined as families having exactly one individual with ASD and at least one sibling without ASD) or non-multiplex families from MSSNG (defined as families having exactly one individual with ASD and no siblings).

To further explore genomic architecture, we compared the burden in multiplex families (where inherited variation would be expected to play a larger role) versus simplex families (*de novo* variation). As expected, *de novo* PTVs in constrained genes were significantly depleted in multiplex families (MSSNG multiplex versus MSSNG non-multiplex: OR=0.5, FDR=0.01; MSSNG multiplex versus SSC simplex: OR=0.5, FDR=0.01) (Figure 5 (B)). We also observed a non-significant enrichment of inherited PTVs in the 135 TADA+ genes in multiplex families. In some cases in which individuals with ASD from multiplex families had ASD-associated rare variants, multiple individuals had the same variant, and in other cases, they had different variants, consistent with previous reports (Yuen et al., 2015); examples of both are given in Supplementary Figure 7).

Overall, one or more ASD-associated rare variants were detected in approximately 16% of individuals with ASD, with the yield nearly identical in MSSNG and SSC (Figure 6). Of these, the largest contributor was dominant sequence-level variants (41%), followed by pathogenic SVs (28%). The large contribution of these variant types explains why the overall yield is similar to previous reports (Tammimies et al., 2015), as the additional variant types examined here appear to make a comparatively smaller contribution to ASD susceptibility. We observed no significant difference between ancestry groups in the overall yield of ASD-associated rare variants (Supplementary Figure 8 (A) and Supplementary Table 18). We also tested whether any ancestry group had a significantly different rate of any particular type of rare variant. After adjusting for multiple tests, one significant difference was detected: an enrichment of TREs in individuals of African descent (Fisher’s Exact Test: OR=5.0, FDR=0.0004) (Supplementary Figure 8 (B) and Supplementary Table 19).

**Figure 6:**
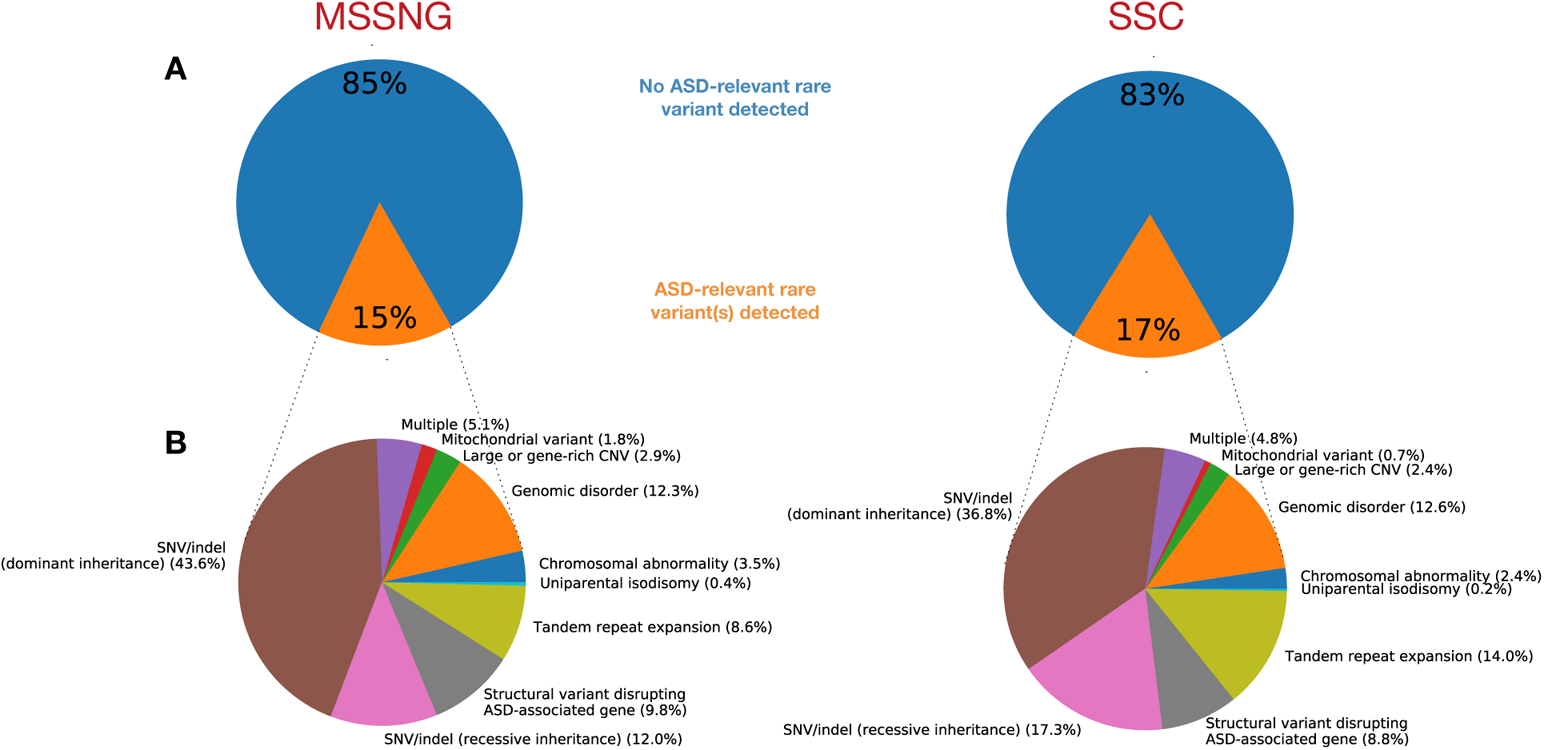
Summary of the genomic architecture of rare variants in ASD. (A) Percentage of individuals with ASD with or without an identified ASD-associated rare variant. (B) Percentage of ASD-associated rare variants in each category. “Multiple” indicates individuals with ASD-associated variants in multiple categories.

To examine genotype-phenotype associations, as well as to illustrate the use of MSSNG’s new composite phenotype measures, we determined the distributions of four measures—adaptive behavior standard score, full-scale IQ, global ability composite estimate, and socialization standard score—in individuals with ASD from MSSNG having each type of ASD-associated rare variant, as well as in those with no such variant. With few exceptions, all categories of ASD-associated rare variants were associated with lower scores across all four composite measures (Figure 7). Using a logistic regression model to compare each rare variant type to no variant and adjusting for sex and ADMIXTURE population components, dominant SNVs/indels were significantly associated with lower scores across all four phenotype measures, and TREs were significantly associated with lower scores for all measures except the global ability composite estimate. Some variant types, such as large and gene-rich CNVs, were consistently associated with lower scores, but were not significant, likely due to lack of power.

**Figure 7:**
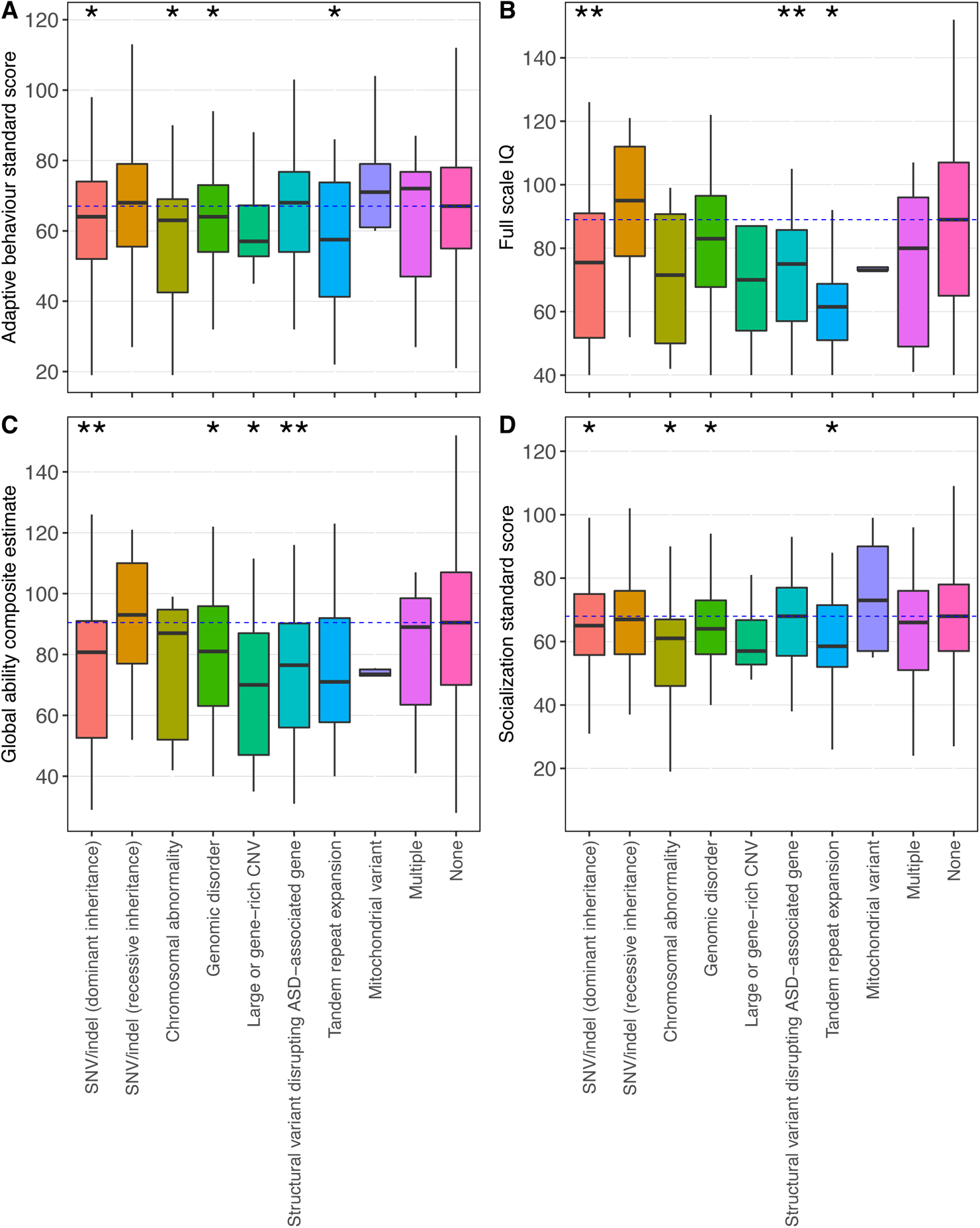
Composite phenotype measure distributions for individuals with ASD having each type of ASD-associated rare variant, or no ASD-associated rare variant (“None”). (A) Adaptive behavior standard score (n=2,787); (B) full-scale IQ (n=1,279); (C) global ability composite estimate (n=1,782); (D) socialization standard score (n=2,765). Asterisks indicate that a given type of rare variant was significantly associated with the phenotype measure (single asterisk: nominally significant with p<0.05; double asterisk: significant after multiple testing correction). The dotted blue line indicates the median of the “None” group.

To give a fine-grained view of the genomic architecture of rare coding variants in ASD, we enumerated the genes and genomic regions impacted (Figure 8 and Supplementary Tables 20-28). Of the genes that were either in the TADA+ list or were ranked definitive in the EAGLE curation process (Schaaf et al., 2020), those most frequently affected by dominant PTVs and damaging missense variants (both *de novo* and inherited) in individuals with ASD included *PTEN*, *KDM5B*, *MIB1*, *CHD8*, *NRXN1*, and *NF1*. Genes commonly impacted by damaging recessive events included *SYNE1*, *NEB*, *OBSCN*, *FAT1*, *DNAH8*, *and DNAH3*. In addition to eight individuals with trisomy 21, all in MSSNG due to SSC inclusion criteria (Fischbach and Lord, 2010), and many instances of sex chromosome aneuploidies, a number of other chromosomal abnormalities were observed, including several translocations. The most frequently detected genomic disorder syndromes included 15q13.3 deletions, along with CNVs (predominantly duplications) at 16p11.2, 1q21.1, 15q11-q13, and 22q11.21. A variety of large and gene-rich CNVs not overlapping with canonical genomic disorder regions were identified, including two each at 8p23.3-p23.1, 5p15.31-p15.2, and 2q23.3-q241. The ASD-associated genes most affected by SVs included *NRXN1*, *PTCHD1-AS*, *AUTS2*, *SCN2A*, *MBD5*, and *DMD*. Top genes for TREs were described previously (Trost et al., 2020) and are recapitulated in Figure 8. Finally, the mtDNA genes most frequently affected by pathogenic variants were *MT-TL1* (mainly causing MELAS) and *MT-ND6* (Leigh Disease).

**Figure 8:**
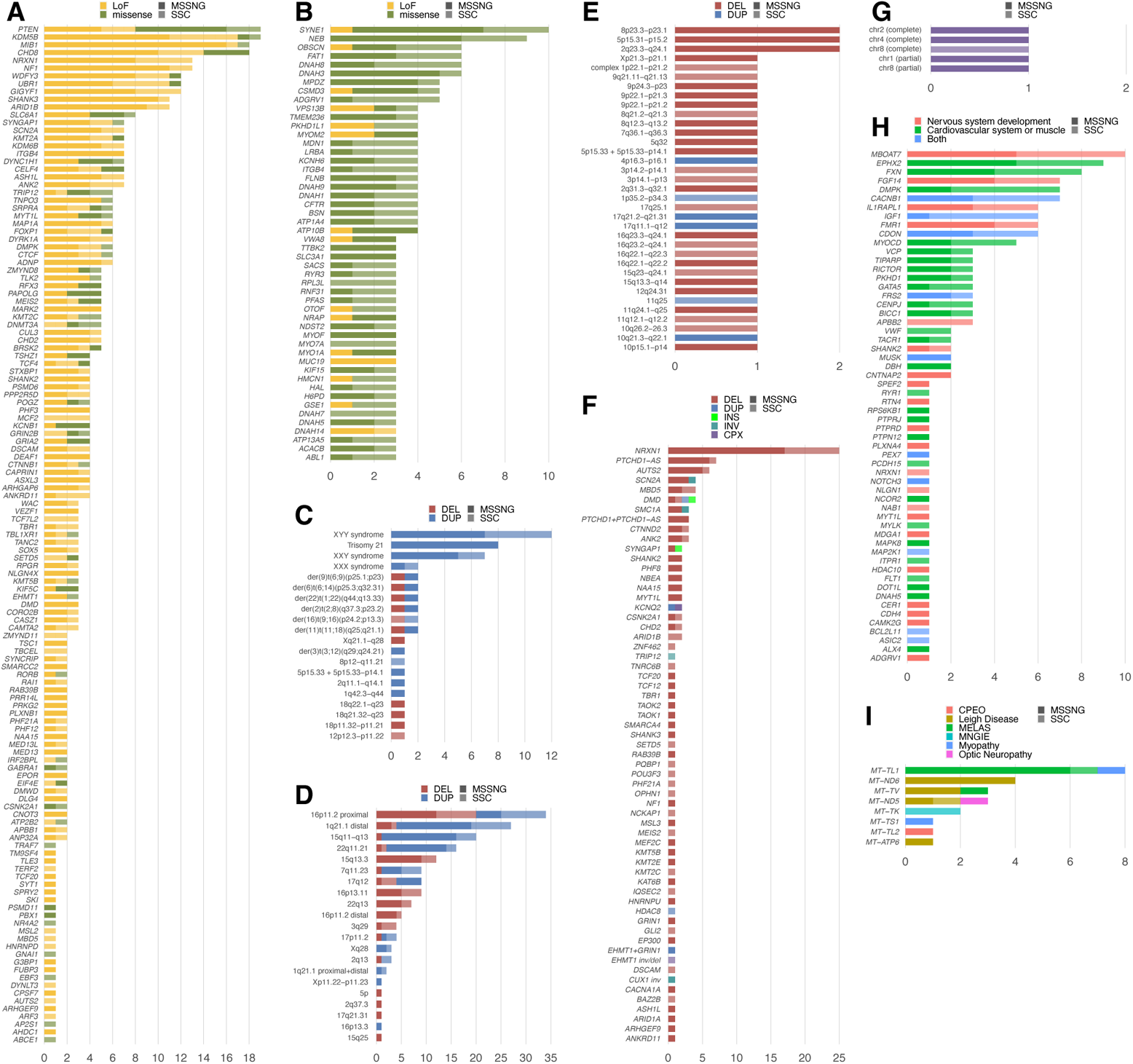
Detailed view of the genomic architecture of rare variants in ASD. Dark versions of a given color represent individuals from MSSNG, while light versions of that same color represent individuals from SSC. (A) *De novo* or rare inherited (gnomAD allele frequency <10^-4^) protein truncating variants (PTVs) and *de novo* damaging missense variants in genes identified by the TADA+ analysis and/or evaluated as definitive by the EAGLE ASD gene curation process (Schaaf et al., 2020). (B) Recessive (homozygous or compound heterozygous) events. (C) Chromosomal abnormalities. (D) Genomic disorders. (E) Large, gene-rich CNVs other than genomic disorders. (F) Structural variants disrupting the 135 TADA+ genes or other genes with strong evidence for a role in ASD or neurodevelopmental disorders. (G) Uniparental isodisomies. (H) Tandem repeat expansions in enriched gene sets identified previously (Trost et al., 2020). (I) Pathogenic mitochondrial variants. CPEO, chronic progressive external ophthalmoplegia; MELAS, mitochondrial encephalomyopathy, lactic acidosis, and stroke-like episodes; MNGIE, mitochondrial neurogastrointestinal encephalomyopathy.

### Non-coding transmission and burden analysis

To investigate the role of non-coding variation, we annotated rare (<0.1% population frequency) or singleton (private to a particular family) sequence-level variants according to whether they impact various regulatory elements, including enhancers, transcription factor binding sites, promoters, topologically associating domains (TADs), and many others (Supplementary Table 29). We first performed transmission bias tests, which compare the number of variants impacting a given non-coding element that were transmitted from a parent to a child with ASD with the corresponding number of non-transmitted variants. The hypothesis underlying transmission bias tests is that variants in elements related to ASD susceptibility will be over-transmitted to individuals with ASD. Genome-wide, the numbers of transmitted and non-transmitted rare variants were nearly equal for most parents in both MSSNG and SSC, regardless of predicted ancestry or sequencing platform, suggesting that the data were suitable for performing transmission bias tests (Supplementary Figure 9). In MSSNG, variants were over-transmitted to individuals with ASD for several types of non-coding elements (FDR <5%), including 5’-untranslated regions (OR=1.03); GeneHancer enhancers (all enhancers (OR=1.01), as well as those intolerant to loss of function (OR=1.01)); transcription factor binding sites from Remap (OR=1.01); and promoters for which the variant is predicted to negatively affect binding (OR=1.02) (Supplementary Figure 10). In SSC, over-transmission was observed in individuals with ASD for two non-coding annotations: transcription factor binding sites targeting the 135 TADA+ genes (OR=1.2); and variants damaging promoters of loss of function intolerant genes (OR=1.2). No annotations exhibited significant over-transmission in SSC unaffected siblings. Similar results were obtained when performing transmission tests using singletons (Supplementary Figure 11).

We next performed burden tests, in which the number of variants in a non-coding element were compared between individuals with ASD and their unaffected siblings. This comparison was done only in SSC due to the small number of unaffected siblings in MSSNG. After correcting for multiple tests, no annotation classes were significantly enriched in individuals with ASD for rare variants (Supplementary Figure 12), singletons (Supplementary Figure 13), or *de novo* variants (Supplementary Figure 14).

### Polygenic risk scores

Much of our knowledge of the genetic architecture of ASD, in particular specific ASD-associated genes and genomic loci, is derived from studies of rare and *de novo* gene-disrupting variants. However, common variation also accounts for part of the heritability of ASD (Gaugler et al., 2014; Klei et al., 2021). Here, we used summary statistics from a recent ASD genome-wide association study (GWAS) of European individuals (Grove et al., 2019) to calculate PRS for individuals of European ancestry in MSSNG, SSC, and 1000G (Supplementary Table 30).

To assess technical reproducibility, we compared PRS in 10 monozygotic twin pairs from MSSNG. Relative to the overall PRS range (−18.6 to 21.4; Supplementary Figure 15 (A)), all between-twin differences in PRS were small (mean=0.31; max=0.81; Figure 9 (A)), and the intra-class correlation coefficient was perfect (R=1). PRS distributions were similar in MSSNG, SSC, and 1000G (Figure 9 (B)). Using individuals with ASD from MSSNG and SSC as cases and individuals from 1000G as controls, higher PRS was associated with higher ASD susceptibility (OR=1.03, p=2.5 × 10^!&^; Nagelkerke’s R^2^=0.0039). After adjusting for sex, individuals with ASD in both MSSNG and SSC had higher mean PRS than unaffected siblings in SSC (logistic regression: adjusted p=1.8 × 10^-2^ and 2.9 × 10^-3^, respectively; Figure 9 (C)). Comparisons with unaffected siblings in MSSNG showed the same trend as unaffected siblings in SSC but were not statistically significant, probably due to insufficient power (samples sizes n=142 versus 1,519). No significant difference in PRS was observed between individuals with ASD in MSSNG versus SSC. We also performed a polygenic transmission disequilibrium test (pTDT), finding that PRS was significantly over-transmitted in individuals with ASD in both SSC (consistent with previous results based on an earlier ASD GWAS (Weiner et al., 2017)) and MSSNG, but not in unaffected siblings (Figure 9 (D)). Stratifying by sex, PRS was significantly over-transmitted in males with ASD in both MSSNG and SSC as well as females in SSC (Supplementary Figure 15 (B)).

**Figure 9:**
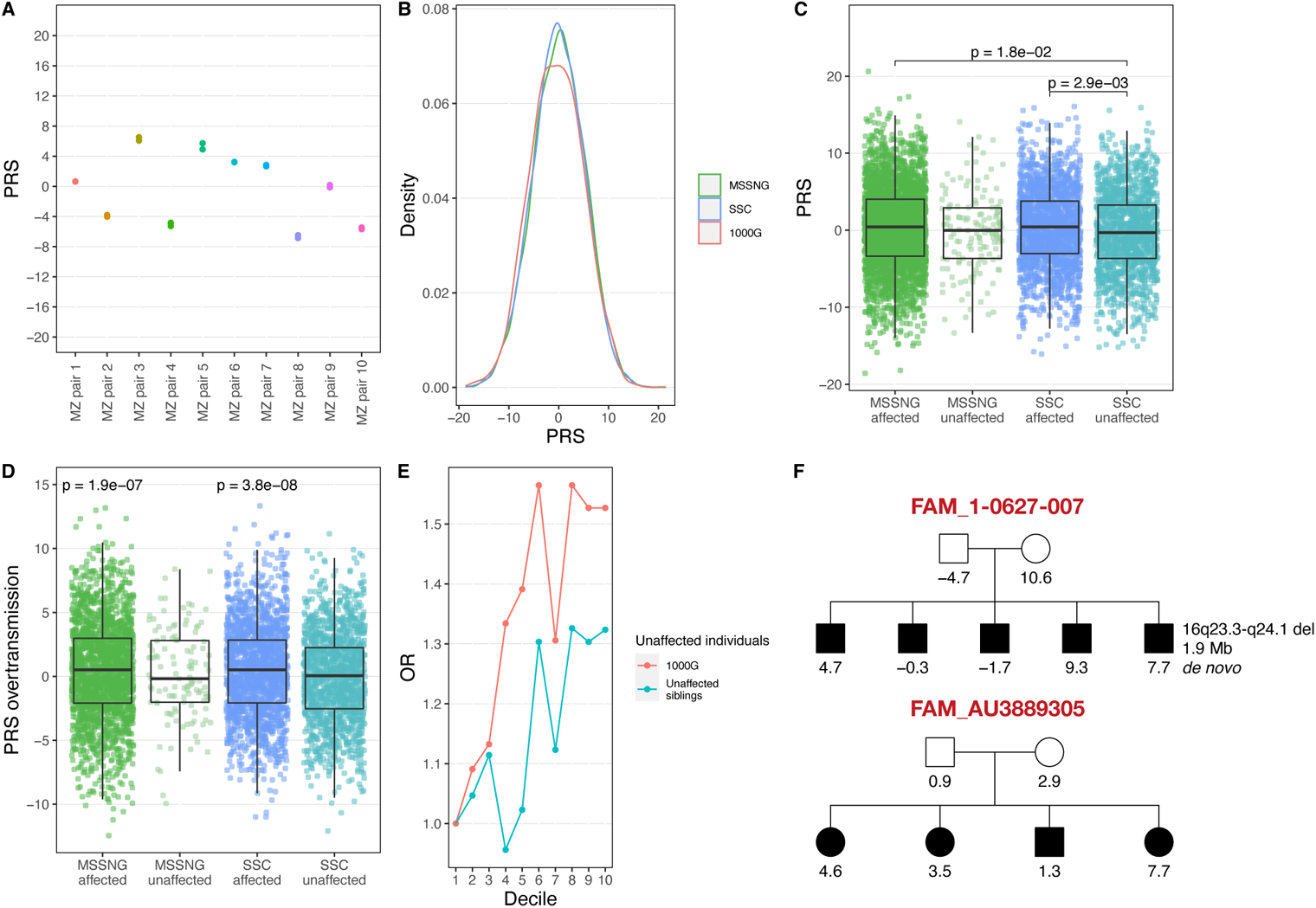
Polygenic risk score (PRS) analysis. (A) Reproducibility in ten pairs of monozygotic (MZ) twins from MSSNG. The PRS of each individual is shown, along with lines depicting the difference in score for each twin pair. (B) PRS distributions in MSSNG, SSC, and 1000G. (C) Comparison between individuals with ASD and their unaffected siblings, stratified by dataset (MSSNG or SSC). Statistically significant differences in means between pairs of groups are indicated; other differences are not significant. (D) Over-transmission of polygenic risk from parents to children in individuals with ASD and their unaffected siblings, stratified by dataset. Values represent the difference between a child’s PRS and the mean of his or her parents. If polygenic risk is over-transmitted in a given group, then the mean should be greater than 0. Statistically significant differences from 0 are indicated; means for other groups were not significantly different from 0. (E) Scatterplot showing the odds ratio (OR) of individuals with ASD to unaffected siblings in each PRS decile (decile 1 = bottom 10%, decile 10 = top 10%), relative to the ratio in decile 1, in MSSNG and SSC combined. (F) Pedigrees indicating the PRS for each individual in two large multiplex families in MSSNG.

As the same trends were observed in MSSNG and SSC, we combined the two datasets to explore patterns in the extremes of the PRS distribution. Specifically, we partitioned the children with ASD and unaffected individuals into PRS deciles and then computed ORs relative to the lowest decile. This was done using two control sets: unaffected siblings from MSSNG and SSC, and individuals from 1000G only. The ORs for individuals in the highest PRS decile were 1.32 and 1.53, respectively, relative to the lowest decile (Figure 9 (E)). MSSNG contains several large multiplex families, affording the unique opportunity to explore PRS at the family level. Here we highlight two families, one with five children with ASD (FAM_1-0627-007) and the other with four (FAM_AU3889305) (Figure 9 (F)). Other than one child in FAM_1-0627-007 with a 1.9 Mb *de novo* deletion at 16q23.3-q24.1, no ASD-associated rare variants were detected in any of the children. In both families, the children have a wide range of PRS: between −1.7 and 9.3 in FAM_1-0627-007 and between 1.3 and 7.7 in FAM_AU3889305. Scores of 7.7 and 9.3 are in the top 93% and 96%, respectively, of the three datasets. In a hypothetical scenario in which these two children were the only children in their respective families, it may have been tempting to attribute their ASD, at least in part, to high polygenic risk. Given the additional context afforded by the lower PRS in the other children with ASD from these families, however, there does not appear to be a strong basis for associating the ASD in these families with polygenic risk. It has been suggested that PRS has, or will have, clinical or predictive utility for many different conditions (Lambert et al., 2019; Lewis and Vassos, 2020). However, this simple example, along with the low percentage of variance explained by PRS in ASD (at least with current data), reinforces the notion that these scores should be interpreted with caution and for ASD are currently unlikely to be informative diagnostically at the level of individuals or families.

We leveraged the family structures in MSSNG and SSC to explore additional trends related to polygenic risk. In SSC sibling pairs (one with ASD and the other unaffected) whose PRS scores differed by more than one standard deviation, the affected sibling had the higher score 58% of the time (binomial test: p=6.8 × 10^−4^), and similarly for a difference of two standard deviations (65%; p=0.01). These observations mirror previous studies comparing polygenic risk in siblings in the context of other disorders (Lello et al., 2020). We detected no significant difference in mean PRS in individuals with ASD from multiplex families compared with simplex families (Supplementary Figure 15 (C)), suggesting that the ASD in multiplex families may be more likely to be attributable to rare, high-impact variants. No significant difference was observed between PRS in mothers of children with ASD versus fathers (Supplementary Figure 15 (D)). We also hypothesized that individuals in 1000G may have a lower mean PRS than parents in MSSNG and SSC, but no significant difference was observed (Supplementary Figure 15 (E)).

Finally, we investigated the association between PRS and the same four composite phenotype measures investigated in the context of rare variants. No significant association was observed between PRS and any of the four measures (Supplementary Figure 16).

### Reproducible and extensible ASD research using Terra and the cloud

Online data and code repositories have many advantages, including data persistence, format standardization, searchability, and version control. However, they also have several drawbacks; in particular, they are poorly suited to very large files (for instance, FASTQ or CRAM) due to the time, expense, and logistics required for researchers to download and store local copies of the data. There are also many issues associated with online code deposition (Perkel, 2021), as the code often fails to reproduce the published results or cannot be executed at all (Pimentel et al., 2019). Cloud computing represents an appealing solution to these issues. Having data storage, compute capabilities, and code in the same place eliminates the wastefulness inherent in each research group storing its own copy of the data and removes the disconnect between data and code, enhancing reproducibility and the potential for extensibility. However, working with and sharing cloud-based data can be technically challenging.

Co-developed by the Broad Institute of MIT and Harvard, Microsoft, and Verily, Terra (https://terra.bio) is a scalable, secure platform for biomedical researchers to access cloud-based data, run analysis tools, and collaborate. It currently supports GCP, with support for other cloud platforms in development. The fundamental unit of organization in Terra is the “workspace”, each of which can contain multiple notebooks (code interleaved with its output) and workflows (chains of programs linked together, typically for more computationally intensive operations).

With the MSSNG data already stored in GCP, Terra is a natural fit for exploring the potential for cloud-based ASD research. To illustrate its use, we have created two Terra workspaces. The first, called terra-mssng-getting-started (https://app.terra.bio/#workspaces/mssng-db6/terra-mssng-getting-started), includes eight notebooks written in Python or R that help researchers get started accessing MSSNG data via Terra, including tutorials illustrating how to query variants, access metadata, and visualize variants in IGV. It also contains two example workflows that illustrate how to run ExpansionHunter (Dolzhenko et al., 2019) and ExpansionHunter Denovo (Dolzhenko et al., 2019) on MSSNG data. The second workspace, called terra-mssng-paper (https://app.terra.bio/#workspaces/mssng-internal-test/terra-mssng-paper), illustrates how Terra can be used to reproduce and extend data analysis. Specifically, it includes two notebooks containing code that generates the figures associated with the MSSNG-specific portions of the non-coding analysis (Supplementary Figures 9-10) and PRS analysis (Figure 9; Supplementary Figures 15-16) from this paper. Researchers can easily inspect the underlying data, run the code on those data themselves, and modify or extend the code by cloning the workspace. All available notebooks and workflows are described in more detail in Supplementary Table 31.

## Discussion

Building upon the latest release of the Autism Speaks MSSNG WGS resource, we have generated a massive and rich set of genetic data, including sequence-level variants, structural variants, tandem repeats, mitochondrial variants, and polygenic risk scores. To meet the needs of users with different research questions or different levels of technical expertise, all of this information, along with extensive phenotype data, are accessible via several interfaces. The MSSNG web portal (https://research.mss.ng) is suitable for users without programming experience or who are interested in extracting variants from a handful of genes or regions. For phenotype information, the Data Explorer component of the MSSNG portal allows users to examine the data at several levels of detail, ranging from sample-level data to dataset-level summaries. MSSNG’s Terra integration is aimed at users with more complex research questions, with the provided “getting started” notebooks allowing even users with little programming experience to quickly begin using Terra to analyze and share data. Finally, advanced users can access MSSNG data via Google BigQuery tables or via flat files stored in Cloud Storage buckets. Together, these interfaces make the MSSNG data accessible and useful to the broadest possible range of ASD researchers.

Importantly, the governance of this autism genomics study explicitly places its participants at the forefront of all decision-making. Clinicians who see families enrolled are heavily involved in the research study, and input is regularly sought from a Participant Advisory Committee. Annual meetings are held with hundreds of participating families discussing new developments in the project, consents are regularly updated with participant input, and the most appropriate methods for communicating and interpreting genomic findings are contemplated by multidisciplinary teams (Hoang et al., 2018; Schaaf et al., 2020; Vorstman and Scherer, 2021). Findings that meet standard clinical reporting criteria are regularly returned to families and accompanied by genetic counselling; hundreds of examples are found in Supplementary Table 5. We are also attempting to capture ancestral genetic diversity and family structure diversity (simplex/multiplex) in MSSNG, including in our own analyses, which will continue to be critical in delineating the role of both rare and common genetic variation (and combinations thereof) in ASD (Weiner et al., 2017; Leblond et al., 2019; D’Abate et al., 2019). In fact, one of our most interesting findings pertaining to ASD genomic architecture was that rare, dominant variation appears to play a predominant role in multiplex ASD, which was evidenced by the depletion of rare, damaging recessive events and lack of enrichment for polygenic risk in multiplex families compared with simplex.

By examining many different types of genetic variants, we detected rare ASD-associated variants in approximately 17% of individuals with ASD. However, it should be emphasized that the current study still falls short of examining every possible type of genetic contribution. For instance, in several published and ongoing studies, the MSSNG data are being examined for somatic (Freed and Pevsner, 2016; Dou et al., 2017; Krupp et al., 2017; Lim et al., 2017; D’Gama, 2021) and epigenetic (Choufani et al., 2015; Yuen et al., 2017; Siu et al., 2019; Chater-Diehl et al., 2021; Rots et al., 2021; Siu et al., 2021) contributors, which may account for 3-5% and 1% of ASD cases, respectively. Epigenetic contributions can be associated with specific genomic variants, environmental exposures, and/or (stochastic) variation in developmental processes. As another example, we examined recessive events only in the context of sequence-level variants; further analysis would be required to uncover the impact of other types of recessive events, such as a large deletion on one allele and a damaging sequence-level variant on the other allele.

It should also be emphasized that the different variant types examined in this study are not equivalent in terms of their penetrance and degree of confidence in their association with ASD. For example, whereas high-impact variants such as pathogenic SVs and rare PTVs in ASD-associated genes can often provide a genetic diagnosis for a given individual, other types of variants, such as common variants and rare non-coding variants, typically cannot. The mitochondrial variants we highlighted are well-known to be pathogenic, but their degree of association with and penetrance for ASD (and at what level of heteroplasmy) are not yet clear. The degree of confidence in the damaging sequence-level variants affecting genes identified in the TADA+ analysis vary, with PTVs in some genes (e.g., *SYNGAP1*, *CHD8*) being the likely cause of the individual’s ASD phenotype, and damaging variants in genes with higher FDRs having more uncertainty. Finally, our recently described TRE analysis (Trost et al., 2020) showed that genes involved in nervous system development and cardiovascular system or muscle are associated with ASD, but we do not yet have the statistical power to determine which of those individual genes increases ASD susceptibility.

With increasing emphasis being placed on genotype-phenotype correlation in precision medicine applications for ASD, individual-level understanding will be key to making earlier diagnoses, which may improve outcomes by allowing treatment programs to begin earlier (Bradshaw et al., 2015), as well as to leveraging information about the detailed circuitry of particular disrupted pathways and processes (Paulsen et al., 2022), which may eventually inform pharmacological interventions (Baribeau and Anagnostou, 2022). This study provides a significant new data resource and analysis roadmap as genome sequencing begins to take its place in mainstream medicine (Costain et al., 2021).

## Methods

### MSSNG whole-genome sequencing and variant detection

DNA quality assessment, quantitation, library preparation, and sequencing were performed as described previously (Yuen et al., 2017). Nearly all new samples in this MSSNG release were sequenced on the Illumina HiSeq X platform, giving 150 bp paired-end reads.

Alignment of reads against the GRCh38/hg38 reference sequence was performed using a pipeline conforming to the Centers for Common Disease Genomics (CCDG) functional equivalence standard (Regier et al., 2018). Sentieon v201808.01 (Kendig et al., 2019), which includes an optimized implementation of BWA (Li and Durbin, 2009), was used to perform alignment, base quality score recalibration, and marking of duplicate reads, producing alignment files in CRAM format. The Sentieon implementation of the Genome Analysis Toolkit (GATK) (McKenna et al., 2010) was used to generate genomic VCF (gVCF) files for each sample. The gVCF files were combined to produce joint-genotyped VCF files (one per chromosome) containing variant calls from all samples. The gVCF files were divided into shards of approximately 50 Mb each to facilitate parallelization of the joint genotyping step. Variant metrics were generated using the CollectVariantCallingMetrics function of Picard. Workflows for read alignment and small variant detection are available as workflow description language (WDL) files on Dockstore/GitHub (https://dockstore.org/search?organization=DNAstack).

### Replication and population control datasets

The Simons Simplex Collection (SSC) (Fischbach and Lord, 2010), which includes WGS data from individuals with ASD and their family members, was used as a replication dataset. Alignment files and small variant calls for 9,205 SSC samples were downloaded from SFARI Base (https://base.sfari.org). Samples from the 1000 Genomes Project (1000G) (1000 Genomes Project Consortium et al., 2015) were used as population control data. Alignment files for high-coverage sequencing data from 2,504 unrelated 1000G samples (Byrska-Bishop et al., 2021) were downloaded from Amazon Web Services, and small variant calls were downloaded from the European Bioinformatics Institute (http://ftp.1000genomes.ebi.ac.uk/vol1/ftp/data_collections/1000G_2504_high_coverage/workin <u>g/20190425_NYGC_GATK</u>). Other than read alignment and small variant detection, all other analyses of SSC and 1000G data (CNV detection, SV detection, mtDNA variant detection, PRS calculations, and variant annotations) were performed using the same pipelines as used for MSSNG.

### Small variant annotation, filtering, and *de novo* detection

Small variants were annotated using a custom ANNOVAR-based pipeline (Wang et al., 2010). To be labeled as high quality, calls were required to have FILTER=“PASS” and depth (DP) >=10. Other criteria based on the genotype quality (GQ) and allelic fraction (AF) also had to be satisfied depending on the variant type. For heterozygous SNVs, also required were GQ ≥ 99 and 0.3 ≥ AF > 0.8. For heterozygous indels, also required were GQ ≥90 and 0.3 ≥ AF > 0.8. For homozygous SNPs and indels, also required were GQ ≥25 and AF ≥0.8. *De novo* variants were detected using DeNovoGear (Ramu et al., 2013) as previously described (Yuen et al., 2017).

### Ancestry determination

We extracted the genotypes for 57,984 positions from a previously published list (https://www.tcag.ca/tools/1000genomes.html) and retained ∼43,000 positions for ADMIXTURE analysis (Alexander et al., 2009) after removing variants with missing calls (genotyping rate <99%) in MSSNG, SSC, and 1000G. We refined population clusters by applying K-means clustering with K=7. Ancestry labels of 1000G samples were reassigned based on the population majority in the refined clusters. We then trained a random forest classifier using the 1000G samples with the re-assigned labels. Ancestry labels in MSSNG and SSC were assigned using separate random forest classifiers. The ancestry assignment for the MSSNG samples was a consensus of the ADMIXTURE analysis and Google’s genomic ancestry inference using deep learning (https://cloud.google.com/blog/products/gcp/genomic-ancestry-inference-with-deep-learning). Self-reported ancestry was used in cases where ADMIXTURE and the deep learning method were discrepant; if no self-reported ancestry was available, the individual was tagged as OTH (other).

### ASD gene list generation

We generated a new ASD gene list by adding more trio data to a previous study by the Autism Sequencing Consortium (ASC) (Satterstrom et al., 2020), which used an enhanced version of the original TADA approach (He et al., 2013) to discover ASD-associated genes based on case-control data and *de novo* variants in trios. Specifically, we added *de novo* variants detected in individuals with ASD in trios from both the MSSNG and Simons Foundation Powering Autism Research (SPARK) (SPARK Consortium, 2018; Feliciano et al., 2019) cohorts.

Because variant quality score recalibration was not applied to the SPARK variant calls, we applied the hard filters recommended by the GATK development team (https://gatk.broadinstitute.org/hc/en-us/articles/360035890471-Hard-filtering-germline-short-variants): Qual By Depth (QD) <2; Fisher Strand (FS) >60; Strand Odds Ratio (SOR) >3; RMS Mapping Quality (MQ) <40; Mapping Quality Rank Sum Test (MQRankSum) <-12.5; and Read Position Rank Sum Test (ReadPosRankSum) <-8. We considered recurrent *de novo* variants more likely to be false, so we manually inspected all recurrent *de novo* variants in IGV to verify their correctness and removed those deemed to be false. We also verified a randomly-selected subset of non-recurrent *de novo* variants in IGV to verify their correctness and *de novo* status.

ASC variant coordinates were converted from GRCh37 to GRCh38 using the NCBI liftover tool. Sample overlap between MSSNG and SPARK was determined using PLINK (Purcell et al., 2007). Because we only had access to *de novo* variants for ASC, sample overlap between ASC and MSSNG and between ASC and SPARK could not be determined using PLINK. Thus, we conservatively removed any sample in MSSNG or SPARK that shared a *de novo* variant with an ASC sample. Variant calls from MSSNG and SPARK were annotated with Missense badness, PolyPhen-2, and constraint (MPC) scores (Samocha et al., 2017) using dbNSFP (Liu et al., 2016). Genes with FDR <0.1 were considered to be ASD-associated.

ASD-associated X-linked genes were detected by identifying rare, hemizygous PTVs in genes with pLI > 0.65 in males with ASD, as described previously (Yuen et al., 2017). Whereas the TADA+ analysis of autosomal genes was performed using data from MSSNG, ASC, and SPARK, for identifying X-linked genes we used MSSNG, SSC, and SPARK (inherited variants from ASC were not available, but SSC was added because sample overlap between SSC and ASC was no longer a factor).

### Gene network diagram

A network diagram representing the ASD-associated genes identified in the TADA+ analysis was generated as previously described (Yuen et al., 2017). Briefly, for each gene we used GeneMANIA (Warde-Farley et al., 2010) to identify the 200 gene neighbors with the closest association based on protein-protein interactions and biological pathways. The network was visualized using Cytoscape (Shannon et al., 2003).

### Identification of recessive events

We identified homozygous or compound heterozygous PTVs and damaging missense (DMis) variants in autosomes that were present in individuals with ASD and were absent from unaffected siblings and controls. Homozygous variants were identified in all individuals with ASD, while compound heterozygous events were identified only in individuals with ASD having both parents sequenced so that phase could be determined. Compound heterozygous events in which one of the variants was a PTV and one was a DMis variant were classified as PTV events.

Variants were filtered with the following criteria: (1) “high quality” as defined above; (2) minor allele frequency <1%. DMis variants were defined as those predicted to be damaging by at least three out of six prediction tools (SIFT (Ng and Henikoff, 2003), PolyPhen2_HDIV (Adzhubei et al., 2010), PolyPhen2_HVAR, MutationTaster (Schwarz et al., 2010), MutationAssessor (Reva et al., 2011), and likelihood ratio test (LRT) (Chun and Fay, 2009)). Biallelically constrained genes were defined as those with fewer than two homozygous PTVs and fewer than 5 homozygous DMis variants in gnomAD. (Compound heterozygous events could not be counted in gnomAD due to the lack of phasing information).

### Detection of copy number variants

For Illumina data, CNVs >=1 kb in size were detected using a pipeline involving the algorithms ERDS (Zhu et al., 2012) and CNVnator (Abyzov et al., 2011) as previously described (Trost et al., 2018). CNV frequencies were calculated separately for three groups of samples—MSSNG parents sequenced by HiSeq X, MSSNG parents sequenced by HiSeq 2000/2500, and 1000 Genomes Project individuals. To avoid reliance on an external ASD cohort, we did not compute SSC parent frequencies. Samples with anomalous CNV counts (more than 3 standard deviations higher than the mean) were not included in frequency calculations. Rare CNVs were defined as those with less than 1% frequency according to both ERDS and CNVnator in each of the three groups, as well as in MSSNG parents sequenced by Complete Genomics. High-quality CNVs were defined as those that were detected by both ERDS and CNVnator with at least 50% reciprocal overlap and for which less than 70% of the CNV overlapped assembly gaps, centromeres, and segmental duplications. “High-quality rare” (HQR) CNVs were defined as those that were both high-quality and rare. HQR CNVs were used for subsequent analysis. *De novo* CNVs were defined as HQR CNVs that were not detected by either ERDS or CNVnator in either parent. Further sample-level quality control was performed after identifying HQR and *de novo* CNVs. Samples with HQR counts more than three standard deviations higher than the mean were tagged as outliers. Due to the smaller number of individuals of non-European ancestry, this was done in European individuals only. The *de novo* CNV counts appeared to have a Poisson distribution, so we applied the Anscombe transformation to normalize the distribution. Samples with transformed counts more than three standard deviations higher than the mean (in any ancestry group) were tagged as outliers.

For Complete Genomics data, CNVs were detected using a proprietary pipeline provided by the company. Frequencies were calculated and rare CNVs were identified in the same way as for Illumina samples. All Complete Genomics CNVs were considered high-quality for the purposes of identifying HQR CNVs. *De novo* CNVs were defined as those present in the child but not present with at least 50% reciprocal overlap in either parent. Subsequent sample-level quality control was performed using the same method as for Illumina samples.

### Detection of structural variants

Unlike for CNVs, we did not have an existing workflow for the detection of SVs from Illumina sequencing data. Thus, we developed a novel workflow specifically for this study by identifying promising candidate algorithms, evaluating the concordance and accuracy of those algorithms, selecting the most accurate combination of algorithms, and then determining effective filtering criteria to attain high sensitivity and specificity.

Dozens of algorithms for detecting SVs from short-read WGS data have been developed (Kosugi et al., 2019). We chose an initial set of algorithms for further evaluation based on these criteria: all algorithms were required to predict most types of SVs (deletions, duplications, insertions, and inversions) and to have high accuracy based on a previous evaluation (Kosugi et al., 2019); at least some had to be under active development; and collectively use a variety of detection strategies (e.g., split reads, anomalous paired-end mapping, local assembly). The algorithms selected for further evaluation were DELLY (Rausch et al., 2012), GRIDSS (Cameron et al., 2017), LUMPY (Layer et al., 2014), Manta (Chen et al., 2016), SoftSV (Bartenhagen and Dugas, 2016), SvABA (Wala et al., 2018), and Wham (Kronenberg et al., 2015) (Supplementary Table 32).

We evaluated the accuracy of each algorithm using two different methods. The first method involved running the algorithms on WGS data from the HuRef (Levy et al., 2007), NA12878 (1000 Genomes Project Consortium et al., 2015), and HG002 (Zook et al., 2020) genomes, and then comparing the SVs detected by each caller to SV benchmarks detected by orthogonal technologies (i.e., other than Illumina short-read sequencing). The HuRef benchmark was as previously described (Trost et al., 2018), but adding PBSV (https://github.com/PacificBiosciences/pbsv) and Sniffles (Sedlazeck et al., 2018) calls made using in-house 100x Pacific Biosciences long-read data. The NA12878 and HG002 benchmarks were derived from previous studies (Kosugi et al., 2019; Zook et al., 2020). The second method involved performing BAM-validation (Trost et al., 2018) on randomly selected calls from each algorithm in order to more fully evaluate specificity. Based on our evaluation, we found that Manta had the best combination of sensitivity and specificity. Although it made more false positives than Manta, we found that DELLY calls that overlapped with Manta calls were useful for giving added confidence to the Manta calls, and were also useful for detecting inversions. Thus, we selected Manta and DELLY as the basis of our SV-detection workflow.

After algorithm selection, we determined whether the number of anomalously mapped paired-end reads and split reads supporting a given variant call could be used as filters to distinguish true calls from false ones. We found that neither variable was useful for this purpose (Supplementary Figures 17-18). Next, for each variant type, we evaluated the sensitivity and specificity of various caller and stringency combinations (Manta PASS, DELLY PASS, Manta any, or DELLY any), and developed criteria that optimized sensitivity while maintaining good specificity (Supplementary Table 33).

Prior to calculating SV frequencies, we computed the number of SVs detected for each sample in 15 categories based on a combination of SV type (five categories: deletion, duplication, insertion, inversion, or overlapping deletion/duplication) and algorithm (three categories: detected by DELLY, Manta, or both DELLY and Manta). To reduce the impact of false positives, samples for which the call count was more than three standard deviations higher than the mean in two or more categories were excluded from the frequency calculations. As with CNVs, frequencies were calculated separately for MSSNG HiSeq X parents, MSSNG HiSeq 2000/2500 parents, and 1000 Genomes Project individuals. Frequencies were calculated separately for DELLY and Manta. Rare CNVs were defined as those with less than 1% frequency in each of the three groups according to both Manta and DELLY, as well as in Complete Genomics MSSNG parents. SVs were defined as high-quality when they satisfied the filtering criteria derived as described above. HQR SVs were defined as for CNVs. HQR SVs were deemed to be *de novo* if they were not detected by either DELLY or Manta with 90% reciprocal overlap in either parent. For further quality control (QC) tagging, HQR and *de novo* counts were subjected to the Anscombe transformation, and samples were tagged as failing QC if they were an outlier for at least one of the five variant types (SVs detected by DELLY alone, Manta alone, or both Manta and DELLY were aggregated for each SV type prior to detecting outliers). QC based on HQR was performed only in samples of European ancestry, as other ancestry groups had fewer samples for comparison and generally had more rare SVs due to the European bias of the reference genome.

SVs in Complete Genomics samples were detected using the company’s proprietary pipeline. Samples having an SV count that was an outlier in at least one SV category (deletion, distal-duplication, distal-duplication-inversion, interchromosomal, inversion, probable-inversion, and tandem-duplication) were excluded from the frequency calculation. Complete Genomics outliers were determined separately for different versions of the variant-detection software.

### Annotation of copy number and structural variants

CNVs and SVs were annotated using a custom R script (v3.6.1) employing the GenomicRanges and data.table libraries. Gene annotations, genomic features, phenotype ontologies, and disease information were downloaded from various sources (Supplementary Table 34).

### Resolution of duplication structures

To investigate the potential to use WGS data to resolve the structures of duplications and understand their impact on genes, we identified rare duplications in a subset of individuals with ASD in MSSNG that overlapped exons from a broad list of ASD candidate genes (Supplementary Table 15). Known recurrent genomic disorders (e.g., 15q11-q13 duplications) were excluded. Duplication structures were analyzed by visualizing CRAM files using IGV (Robinson et al., 2011) and were classified as tandem or complex through manual inspection of paired-end reads and split reads at the duplication breakpoint junctions. Duplications having breakpoints mapping within homologous segmental duplications or LINE elements were considered likely NAHR events. Breakpoint junctions that could not be classified as tandem, complex, or likely NAHR events were considered unresolved. The sequence-level breakpoint coordinates of tandem and complex duplications were determined through analysis of split read sequence in IGV and using BLAT (Kent, 2002). The last nucleotide of a split read before a deviation from the reference sequence was considered the sequence-level breakpoint of the duplication. Breakpoints that could not be resolved to the nucleotide level using split reads were estimated from the location of paired-end reads and read depth changes. The ERDS coordinates were used as an approximation of the true duplication breakpoints for likely NAHR and unresolved duplications. Duplications with identical breakpoints that were found in multiple related or unrelated individuals were deemed to represent a single unique event. The impact of each duplication on the ASD candidate gene(s) was annotated manually using the “NCBI RefSeq genes, curated subset” track of the UCSC Genome Browser (Speir et al., 2016). A duplication was considered to increase gene dosage when all RefSeq isoforms were fully contained within the duplication. The reading frame of intragenic duplications and fusion genes created at the breakpoints of a duplication were assessed using the UCSC Genome Browser.

### Detection of uniparental disomies

Whole-chromosome and large segmental uniparental disomies (UPDs) were identified using SNPs and CNVs. For a subset of common SNPs, Mendelian errors were calculated using PLINK (Purcell et al., 2007), and samples with excessive errors on a given chromosome were identified. Candidate regions were compared with CNV calls to exclude regions overlapping deletions. Putative isodisomies were further verified by checking that the SNP genotypes were mostly homozygous. Log R ratio (LRR) and B-allele frequency (BAF) plots were generated for the entire family to visualize UPDs.

### Calculation of polygenic risk scores

We calculated PRS in individuals of European ancestry from the MSSNG, SSC, and 1000G cohorts using PLINK v1.9 (Purcell et al., 2007). For MSSNG, PRS was calculated only for individuals sequenced on Illumina platforms. We used summary statistics from a previously-published ASD meta-GWAS, which included the iPSYCH and Psychiatric Genomics Consortium (PGC) cohorts (Grove et al., 2019). Due to sample overlap between PGC and both MSSNG and SSC, we used summary statistics derived only from iPSYCH (13,076 cases and 22,664 controls).

We included only SNPs with minor allele frequency >0.05 (iPSYCH controls) and imputation quality score (INFO) >0.9. To avoid potential strand conflicts, complementary SNPs were excluded. We also excluded SNPs with FILTER != PASS and those within the broad MHC region (chr6:25,000,000-35,000,000). Data from MSSNG, SSC and 1000G were merged prior to clumping, and only SNPs common to all three cohorts were retained. Clumping was performed with an r^2^ threshold and radius of 0.1 and 500 kb, respectively. PRS values were generated by including SNPs with p-value ≤ 0.1, weighting by the additive scale effect (log_10_ OR) of each variant, and then summing over the variants. Scores were centred to a mean of zero.

### Non-coding annotations

We used Ensembl Variant Effect Predictor (VEP) v102 (McLaren et al., 2016) and the October 2019 release of ANNOVAR (Wang et al., 2010) to perform non-coding variant annotations based on information from several databases. The variant data from each ASD WGS cohort were converted into ANNOVAR- and VEP-compatible format, with multi-allelic variants split and indels normalized to ensure correct matching of variants. Selected files from the non-coding variant databases (see below) were also converted to ANNOVAR-compatible format.

Regulatory features from Ensembl Regulatory Build (http://useast.ensembl.org/info/genome/funcgen/regulatory_build.html) (Zerbino et al., 2015) were added using VEP. Annotations related to promoters, enhancers, and their target genes were obtained from the GeneHancer database (geneHancerInteractionsDoubleElite, UCSC update January 2019) (Fishilevich et al., 2017). Transcription factor binding sites and other regulatory elements were derived from the ReMap2020 non-redundant peak file (http://remap.univ-amu.fr) (Chèneby et al., 2020). Long non-coding RNA annotations were obtained from LNCipedia v5.2 (https://lncipedia.org) (Volders et al., 2013). Small non-coding RNA annotations were acquired from DASHR v2.0 (https://dashr2.lisanwanglab.org) (Kuksa et al., 2019). Retrotransposon insertion polymorphism annotations were obtained from dbRIP (https://dbrip.brocku.ca) (Wang et al., 2006).

Topologically associating domain (TAD) boundaries were derived from a previous publication (Dixon et al., 2012), and five annotations were generated using the TAD boundaries. The first three annotations had possible values of 0, 1, or 2, depending on whether neither TAD flanking the boundary contained genes from a given gene list (0), one of the two TADs did (1), or both did (2). The three gene lists used were the ASD-associated genes from our TADA+ analysis (135 genes), genes associated more generally with neurodevelopmental disorders (1,250 genes), or genes intolerant to loss of function (pLI >0.9) (2,867 genes) (Lek et al., 2016). The 1,250 NDD-associated genes were derived from the literature (excluding those having autosomal recessive inheritance) (Pinto et al., 2014; Sanders et al., 2015; Satterstrom et al., 2020), SFARI genes (score 1-3 plus syndromic), genes with ClinGen haploinsufficiency or triplosensitivity scores of 2 or 3 (Riggs et al., 2012), and the Geisinger Developmental Brain Disorder Gene Database (https://dbd.geisingeradmi.org). The remaining two annotations were based on the “brain expression specificity” of the left and right TAD—that is, the fraction of genes in each TAD that are expressed in the brain based on Illumina Body Map 2.0 RNA-seq data. Specifically, let *B_L_* and *B_R_* be the proportion of genes that are brain-expressed in the left and right TAD, respectively. Then the first annotation represented the difference in brain expression specificity (|*B_L_* − *B_R_*|) and the second the sum of brain expression specificity (*B_L_* − *B_R_*).

### Non-coding transmission bias tests

We defined the odds ratio for over-transmission of non-coding variants with annotation *A* as OR = (*A_T_/A_NT_*) / (*O_T_/O_NT_*), where *A_T_* is the number of transmitted non-coding variants with annotation *A*, *A_NT_* is the corresponding number of non-transmitted variants, and *O_T_* and *O_NT_* are the total number of transmitted and non-transmitted non-coding variants, respectively, for all annotations other than *A*. To reduce noise, only variants with PhastCons score >0 were considered. To avoid the case where the expected transmission rate is not equal to 50% (for example, when both parents were heterozygous for a given variant, or one or both parents were homozygous), we included only variants for which one parent was heterozygous and the other did not have the variant. Complete Genomics samples were not included in the non-coding transmission tests because their genome-wide transmission rate differed from the expected 50%.

### Burden tests

We used logistic regression to compare individuals with ASD with unaffected siblings. The covariates used were sex, presence or absence of an ASD-associated rare variant (for non-coding analysis only), population structure variables from ADMIXTURE (Alexander et al., 2009), and the total number of variants of the type being tested. FDRs were determined using the Benjamini-Hochberg method.

### Mitochondrial Analysis

For samples sequenced on Illumina platforms, reads aligning to the mitochondrial genome were extracted and realigned to the revised Cambridge Reference Sequence (NC_012920) using BWA v0.7.8. Pileups were generated with SAMtools mpileup v1.1 (Li, 2011), requiring the program to include duplicate reads and retaining all positions in the output. Custom scripts were developed to parse the mpileup output to determine the most frequently occurring non-reference base at each position. The heteroplasmic fractions were calculated and VCF files were generated. For samples sequenced by Complete Genomics, mitochondrial variants detected by the proprietary software were extracted. For both platforms, FASTA files replacing mitochondrial reference bases with alternative bases at sites where the heteroplasmic fraction was greater than or equal to 0.5 were also generated and haplogroups were predicted using HaploGrep v2.1.1 (Weissensteiner et al., 2016). The VCF files were annotated using ANNOVAR-based custom scripts with annotations from MitoMaster (Lott et al., 2013) (April 2019) and Ensembl v96.

### Composite phenotype measures

Composite scores across several domains were calculated to facilitate the use of varied behavioral data across cohorts. These measures are not meant to replace the detailed and strategic work that a clinical psychologist or a computational scientist may choose to do with these data, but rather to provide reasonable ways of summarizing cognitive/behavioral data across domains of interest, especially for researchers who have less familiarity with the underlying tests. These measures represent a work in progress, and we welcome feedback on their design and utility as well as ideas for additional composite measures. Below, we describe the composite measures currently available in MSSNG.

#### Adaptive behaviour standard score

Two measures capture this domain within MSSNG: Adaptive Behavior Assessment System (ABAS) and Vineland Adaptive Behavior Scales Survey Interview Form (Vineland). The computed score is the global adaptive composite (GAC) score from the ABAS or the adaptive behavior standard score from the Vineland. If more than one instrument is available, the most recent score is used. There is active work being done to create a reliable correction strategy that will allow the use of scores from both instruments to be used as a continuous measure (Dupuis et al., 2021).

#### Socialization standard score

This score is also calculated from the ABAS or Vineland, but the social composite score of the ABAS or the socialization standard score of the Vineland are used. If more than one instrument is available, the most recent score is used.

#### Full-scale IQ

While there are several IQ instruments in MSSNG, we have decided to include only Wechsler scales and Stanford-Binet under this category. Full Scale IQs (FSIQs) are prioritized over abbreviated IQs and Wechsler scales are prioritized over Stanford-Binet. Among FSIQs, the algorithm selects the most recent one if they are more than two years apart.

#### Global ability composite estimate

This score is intended to estimate global abilities when gold standard full IQ measures are not available. If a full-scale IQ is available as per above, then this category is populated by that value. Otherwise, the following measures are used to populate this variable, in descending order of preference. The first choices are the Mullen Scales of Early Learning composite scores, followed by Merrill-Palmer (only when all three subdomain scores are available). Subsequently, nonverbal IQ can be used: Leiter International Performance Scale, followed by Raven’s Progressive Matrices. Lastly, verbal standard scores from language assessments can be used in the following order: Clinical Evaluation of Language Fundamentals (CELF), Oral and Written Language Scales (OWLS), Preschool Language Scale (PLS), Expressive Vocabulary Test (EVT), and the Peabody Picture Vocabulary Test (PPVT). This is the category with the most noise and involving a clinical psychologist is highly recommended, depending on the research aim. Within each category, the most recent test is selected.

#### Co-occurring conditions (ADHD, anxiety, seizures, and gastrointestinal conditions)

The available data across cohorts is extremely variable and may include everything from single items (e.g., parental report of a condition) to standardized instruments (e.g., Child Behavior Checklist (CBCL), Symptoms and Normal Behaviors (SWAN) score, Conners Comprehensive Behavior Rating Scales, Revised Children’s Anxiety and Depression Scale (RCADS), and Spence Children’s Anxiety Scale). In all categories, a non-stringent approach was taken so that if any available scores across any measures reflect clinical concern, then the co-occurring condition variable is labeled as true (e.g., ADHD=true). If none of the available measures suggest clinical concern, then the co-occurring condition variable is labeled as false (e.g., ADHD=false). These measures should not be considered evidence of diagnoses, but rather simply of clinical concern.

## Data Availability

Access to the data contained in MSSNG and SSC can be obtained by completing data access agreements at https://research.mss.ng and https://www.sfari.org/resource/sfari-base, respectively. The 1000G WGS data are publicly available via Amazon Web Services (https://docs.opendata.aws/1000genomes/readme.html).

## Data Availability

Access to the data contained in MSSNG and SSC can be obtained by completing data access agreements at https://research.mss.ng and https://www.sfari.org/resource/sfari-base, respectively. The 1000G WGS data are publicly available via Amazon Web Services
(https://docs.opendata.aws/1000genomes/readme.html).

https://base.sfari.org

https://docs.opendata.aws/1000genomes/readme.html

## Acknowledgements

The authors wish to acknowledge the resources of MSSNG (www.mss.ng), Autism Speaks, and The Centre for Applied Genomics at The Hospital for Sick Children, Toronto, Canada. We also thank the participating families for their time and contributions to this database, as well as the generosity of the donors who supported this program. We thank Anders Børglum and Jakob Grove for providing the iPSYCH-only summary statistics. We are grateful to all of the families at the participating Simons Simplex Collection and SPARK sites, as well as the principal investigators (A. Beaudet, R. Bernier, J. Constantino, E. Cook, E. Fombonne, D. Geschwind, R. Goin-Kochel, E. Hanson, D. Grice, A. Klin, D. Ledbetter, C. Lord, C. Martin, D. Martin, R. Maxim, J. Miles, O. Ousley, K. Pelphrey, B. Peterson, J. Piggot, C. Saulnier, M. State, W. Stone, J. Sutcliffe, C. Walsh, Z. Warren, E. Wijsman). We appreciate obtaining access to genetic and phenotypic data on SFARI Base. Approved researchers can obtain the SSC and SPARK datasets by applying at https://base.sfari.org. This work was supported by the University of Toronto McLaughlin Centre, Genome Canada/Ontario Genomics, Genome BC, the Government of Ontario, the Canadian Institutes of Health Research (CIHR), the Canada Foundation for Innovation (CFI), Autism Speaks, Autism Speaks Canada, Brain Canada, Kids Brain Health Network, Qatar National Research Fund (NPRP10-0202-170320), Ontario Brain Institute and SickKids Foundation. We thank Roche for the use of PacBio sequencing data from the HuRef genome. B.T. has been supported by the CIHR Banting Postdoctoral Fellowship and the Canadian Open Neuroscience Platform (CONP) Research Scholar Award. L.O.L. held the Lap-Chee Tsui Fellowship for Research Excellence. S.G. has been supported by the CONP Research Scholar Award. M.H.Z. acknowledges the generous support of the Fonds de recherche du Québec–Santé Junior 1 Research Scholar programme. J.S. and the REACH project were supported by grants from the National Institutes of Health (MH113715, MH119746, 1MH109501) and the Simons Foundation Autism Research initiative (SFARI 606768). L.Z. holds the Stollery Children’s Hospital Foundation Chair in Autism. M.E.S.L. holds a BC Children’s Hospital Research Institute Investigator Grant Award. S.W.S. holds the Northbridge Chair in Pediatric Research at The Hospital for Sick Children and the University of Toronto.

## Ethics approval

Approval for use of the AGRE data was obtained from WCG IRB (https://www.wcgirb.com). Approval for other cohorts was obtained from the Research Ethics or Institutional Review Board of each recruiting site, including Montreal Children’s Hospital—McGill University Health Centre, McMaster University—Hamilton Integrated, Memorial University—Eastern Health, Holland Bloorview Kids Rehabilitation Hospital, Queen’s University, University of Alberta, University of British Columbia, IWK Health Centre, University of California Davis, University of California San Diego, University of Miami, and The Hospital for Sick Children.

## Inclusion and diversity statement

We worked to ensure sex balance in the recruitment of human subjects (the male:female ratio for individuals with ASD in MSSNG closely mirrors the well-established 4:1 sex bias in ASD). We worked to ensure ethnic or other types of diversity in the recruitment of human subjects. We worked to ensure that the study questionnaires were prepared in an inclusive way. The author list of this paper includes contributors from the location where the research was conducted who participated in the data collection, design, analysis, and/or interpretation of the work.

