## Supplementary Information and Figures for "Genomic architecture of Autism Spectrum Disorder from comprehensive whole-genome sequence annotation"

### Supplementary Results

#### Sample quality control

To assess the quality and comparability of the WGS samples used in this study, we stratified the samples into categories based on dataset (MSSNG, SSC, or 1000G), sequencing platform (Complete Genomics or Illumina HiSeq 2000/2500/X), and DNA library preparation method (PCR-based or PCR-free) and then examined the distributions of SNV (Supplementary Figure 19) and indel (Supplementary Figure 20) counts for various classes of variants (all, rare, rare exonic, rare damaging missense (DMis), rare protein-truncating variants (PTVs), or *de novo*). Only high-quality variants (as defined in Methods) were included. All categories had similar distributions, except that Complete Genomics samples differed from the others in terms of indel counts, in particular with fewer indels overall but more *de novo* indels (Supplementary Figure 20).

#### Tandem repeat data quality control

Because samples were added to MSSNG after our study on tandem repeat expansions in ASD (Trost et al., 2020), we ran ExpansionHunter Denovo v0.7.0 (EHdn) (Dolzhenko et al., 2020) on the additional samples, and then repeated QC measures using the full dataset. Each cohort comprising MSSNG had similar distributions of the number of tandem repeat loci detected by EHdn (Supplementary Figure 21 (A)). As we observed previously (Trost et al., 2020), samples sequenced on the Illumina HiSeq 2000 platform had substantially higher call counts than those sequenced on the HiSeq X or HiSeq 2500 platforms (Supplementary Figure 21 (A)). Thus, HiSeq 2000 samples were excluded. Samples sequenced using PCR-based DNA library preparation generally had higher call counts than those using PCR-free library preparation (Supplementary Figure 21 (B)); however, there was substantial overlap in the PCR-based versus PCR-free distributions, so PCR-based samples were retained.

Subsequently, we tagged samples that were outliers in terms of call counts or principal components (PCs). After removing HiSeq 2000 samples, samples having call counts greater than three standard deviations higher than the mean were tagged as failing QC. For QC based on PCs, the EHdn output was converted to a matrix, where the value of a given cell was the size reported by EHdn for a given sample-locus combination (loci not called by EHdn in a given sample were given a value of 0). Principal component analysis was then performed with that matrix as input using the R function *prcomp*. A handful of samples were extreme outliers in terms of some PCs (Supplementary Figure 22), hindering outlier detection for the remaining samples. Thus, two rounds of outlier detection were performed: first removing samples with PC outside mean +- 3SD, recomputing the mean and standard deviation, and then again tagging samples outside mean +- 3SD. Any sample that was an outlier in either round of outlier detection for any of the first five PCs was deemed to have failed QC.

#### Supplementary figures

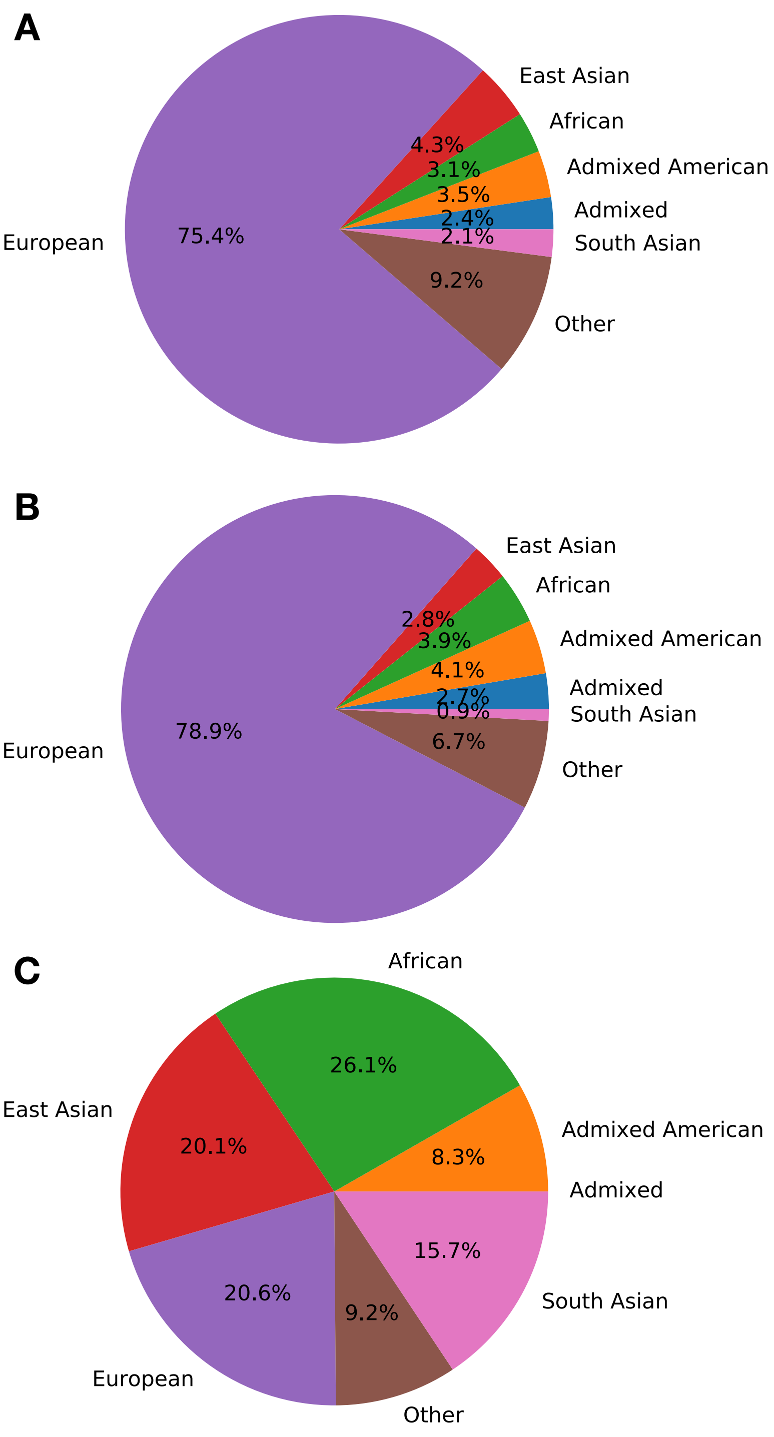


**Supplementary Figure 1:** Genotype-computed ancestry of individuals from (A) MSSNG, (B) Simons Simplex Collection, and (C) 1000 Genomes Project.

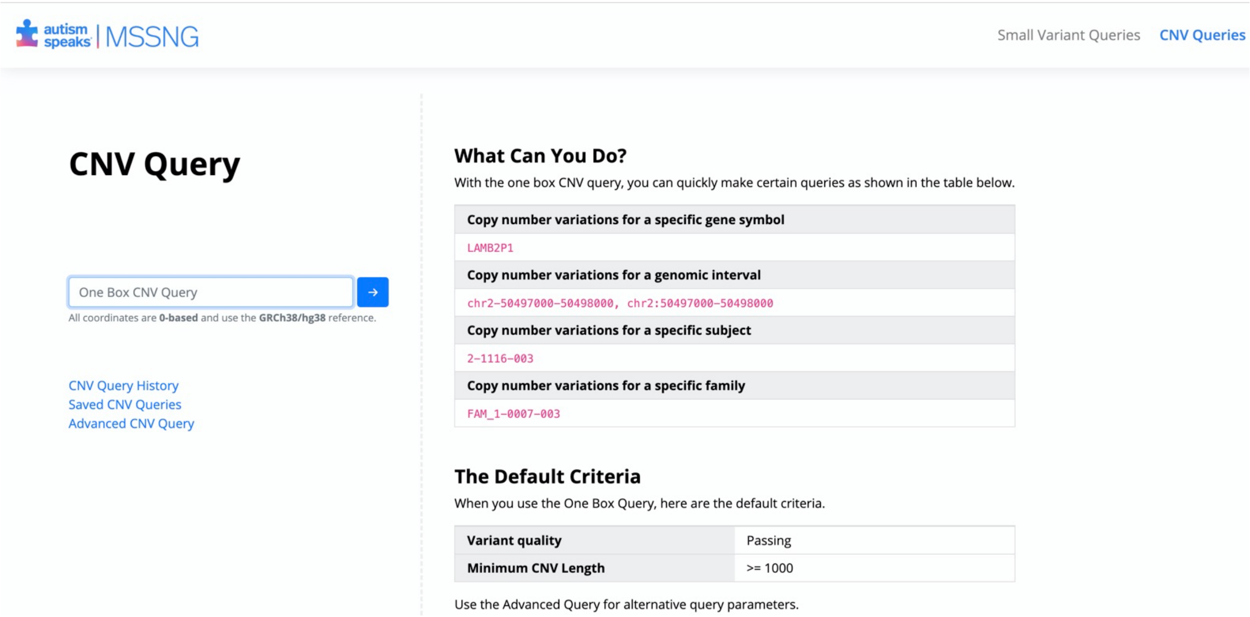


**Supplementary Figure 2:** Screenshot of the interface for querying copy number variants in the MSSNG portal.

A

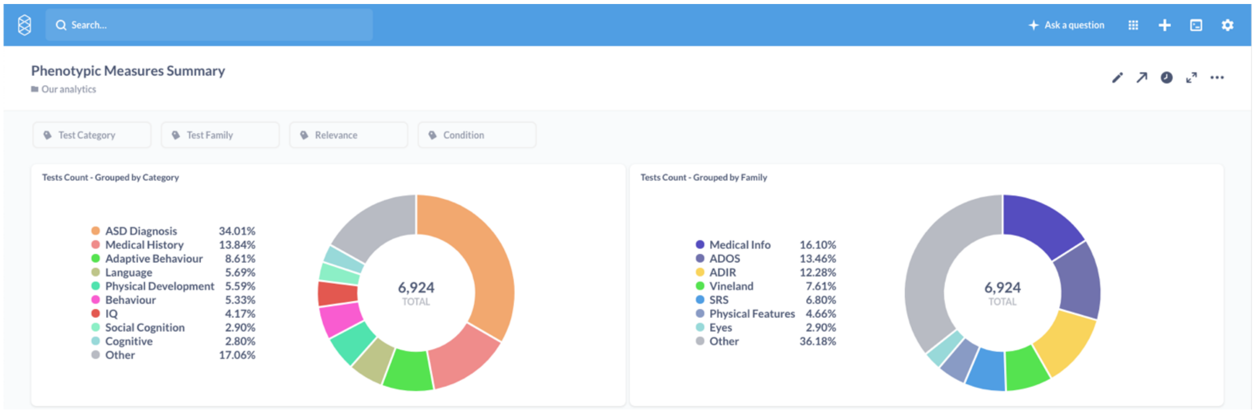
Search...
Ask a question
Phenotypic Measures Summary
Our analytics
Test Category
Test Family
Relevance
Condition
Tests Count - Grouped by Category
ASD Diagnosis 34.01%
Medical History 13.84%
Adaptive Behaviour 8.61%
Language 5.69%
Physical Development 5.59%
Behaviour 5.33%
IQ 4.17%
Social Cognition 2.90%
Cognitive 2.80%
Other 17.06%
6,924
TOTAL
Tests Count - Grouped by Family
Medical Info 16.10%
ADOS 13.46%
ADIR 12.28%
Vineland 7.61%
SRS 6.80%
Physical Features 4.66%
Eyes 2.90%
Other 36.18%
6,924
TOTAL

B

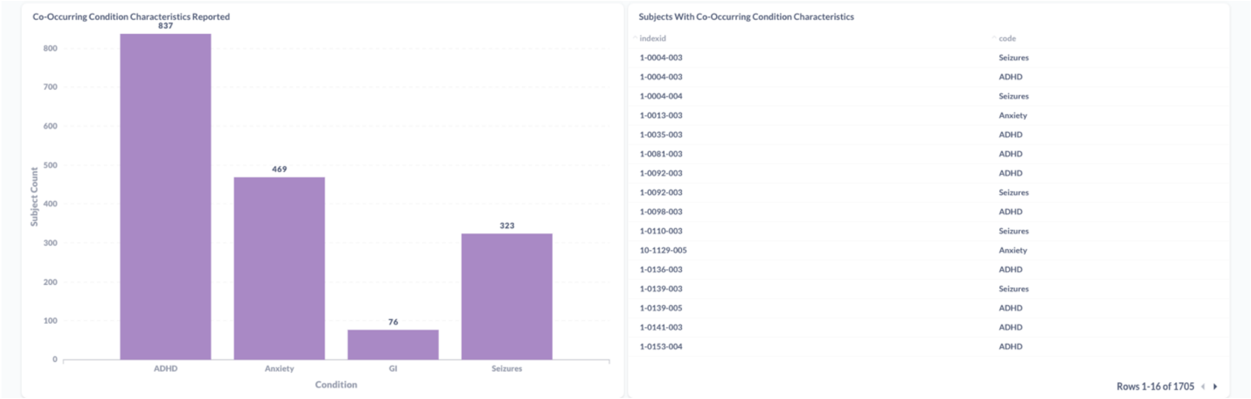
Co-Occurring Condition Characteristics Reported
837
469
76
323
Subject Count
Condition
ADHD
Anxiety
GI
Seizures
Subjects With Co-Occurring Condition Characteristics
indexid
code
1-0004-003 Seizures
1-0004-003 ADHD
1-0004-004 Seizures
1-0013-003 Anxiety
1-0035-003 ADHD
1-0081-003 ADHD
1-0092-003 ADHD
1-0092-003 Seizures
1-0098-003 ADHD
1-0110-003 Seizures
10-1129-005 Anxiety
1-0136-003 ADHD
1-0139-003 Seizures
1-0139-005 ADHD
1-0141-003 ADHD
1-0153-004 ADHD
Rows 1-16 of 1705

C

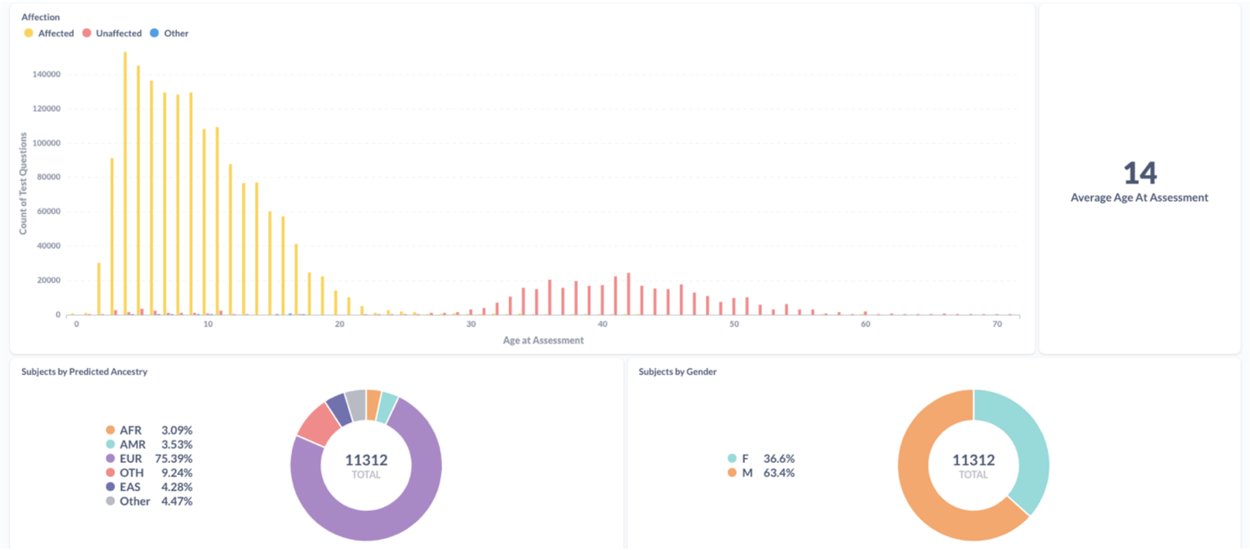


D

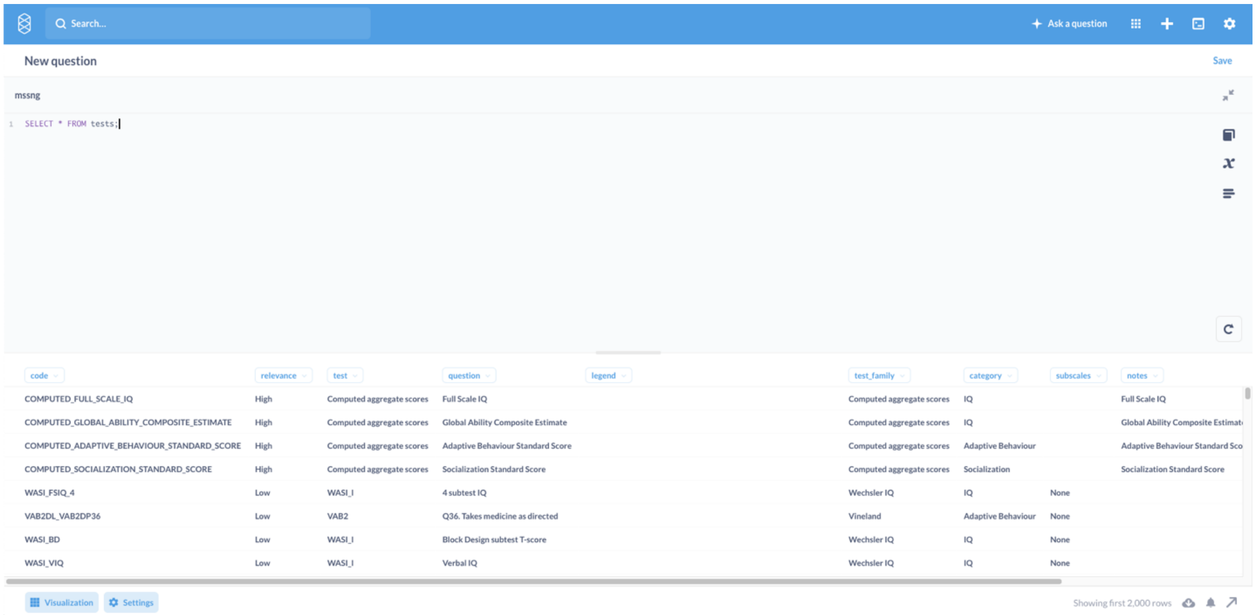


**Supplementary Figure 3:** Screenshots of the Phenotype Data Explorer, showing (A) categorization of tests, (B) co-occurring conditions, (C) summary information involving ancestry, sex, and age at assessment; and (D) results when querying individual test scores.

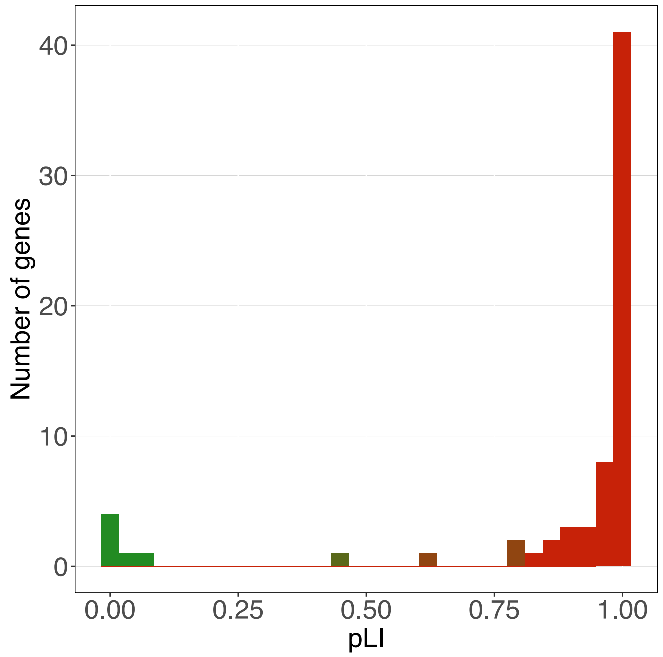


**Supplementary Figure 4:** Distribution of pLI values for the 68 ASD-associated genes not reported in the previous TADA+ analysis (Satterstrom et al., 2020).

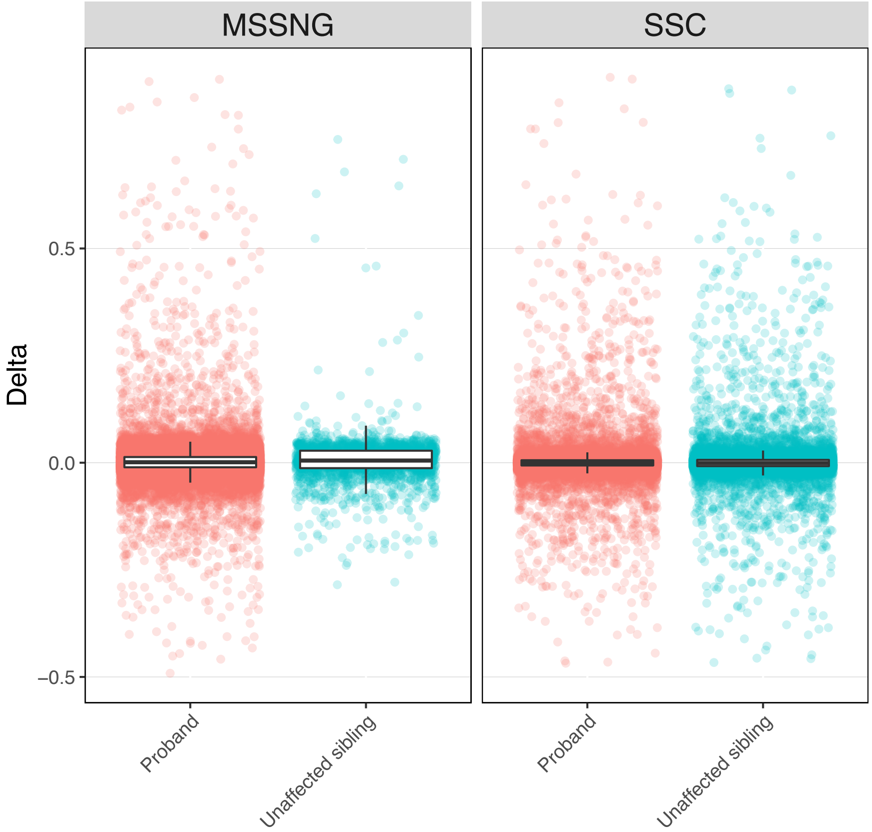


**Supplementary Figure 5:** Distributions of heteroplasmy deltas (child heteroplasmy minus mother heteroplasmy for a given position in the mitochondrial genome sequence) in individuals with ASD versus unaffected siblings in MSSNG (variants tested: ASD n=35,279; unaffected sibling n=2,171) and SSC (ASD n=29,005; unaffected sibling n=18,328).

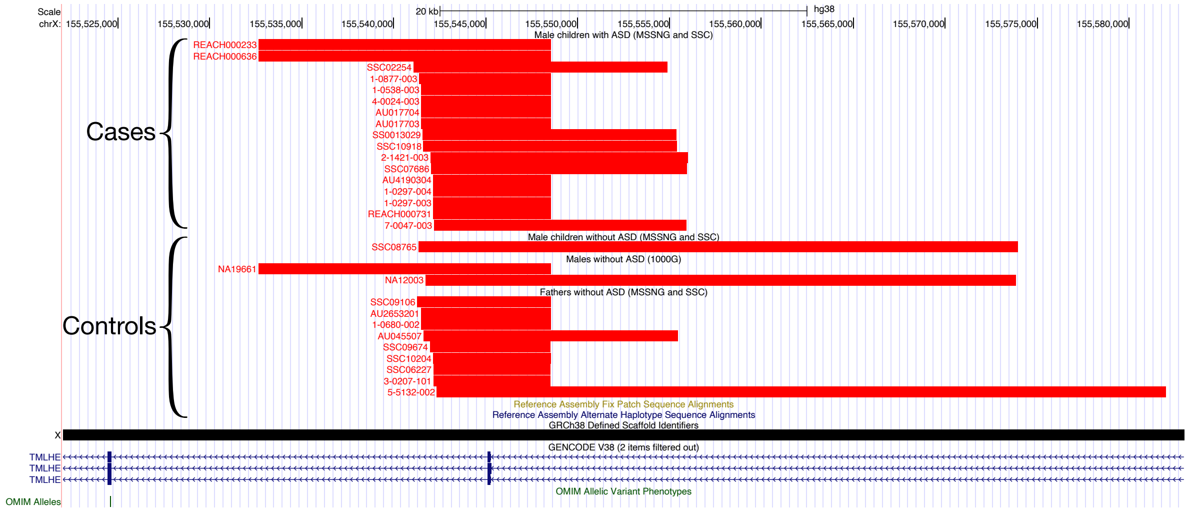


**Supplementary Figure 6**: Deletions in males overlapping exon 2 of *TMLHE*. Deletions are categorized as being in children with ASD in MSSNG or SSC, children without ASD in MSSNG or SSC (i.e., unaffected siblings), population controls from the 1000 Genomes Project, and unaffected fathers in MSSNG or SSC.

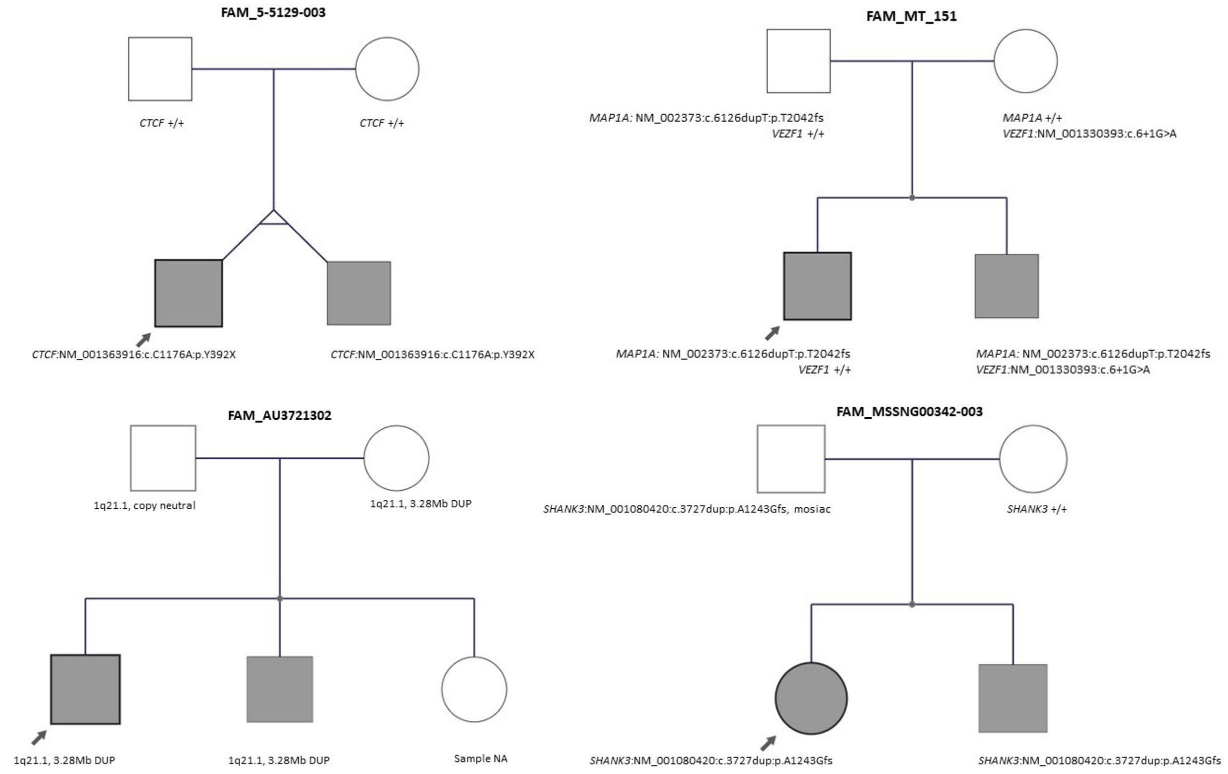


**Supplementary Figure 7:** Examples of multiplex families from MSSNG with ASD-associated rare variants.

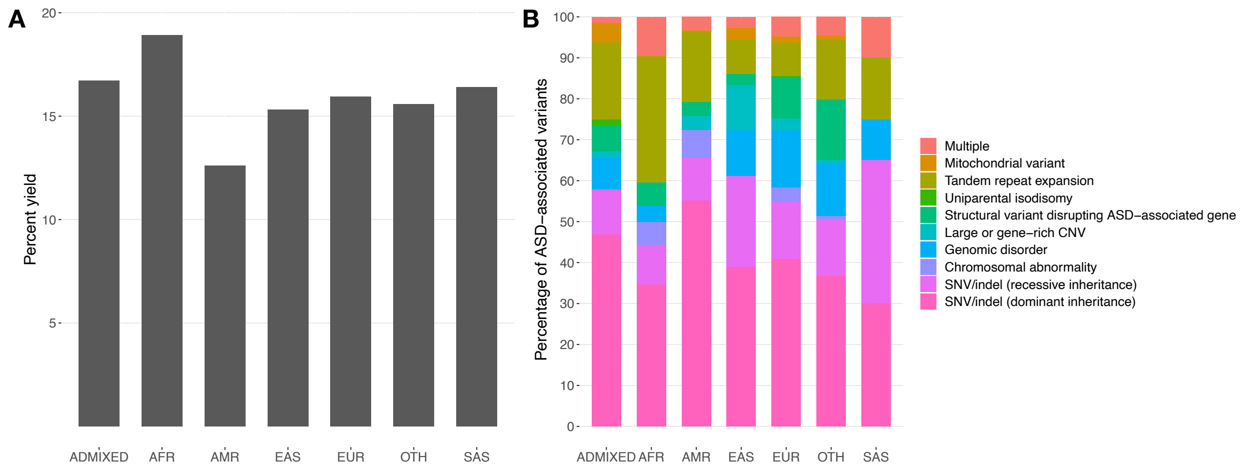


**Supplementary Figure 8:** ASD-associated rare variant yield and composition, stratified by ancestry group. (A) Percentage of individuals with ASD in each ancestry group having an ASD-associated rare variant. (B) Percentage of individuals with ASD with an ASD-associated rare variant having a rare variant in each category.

A

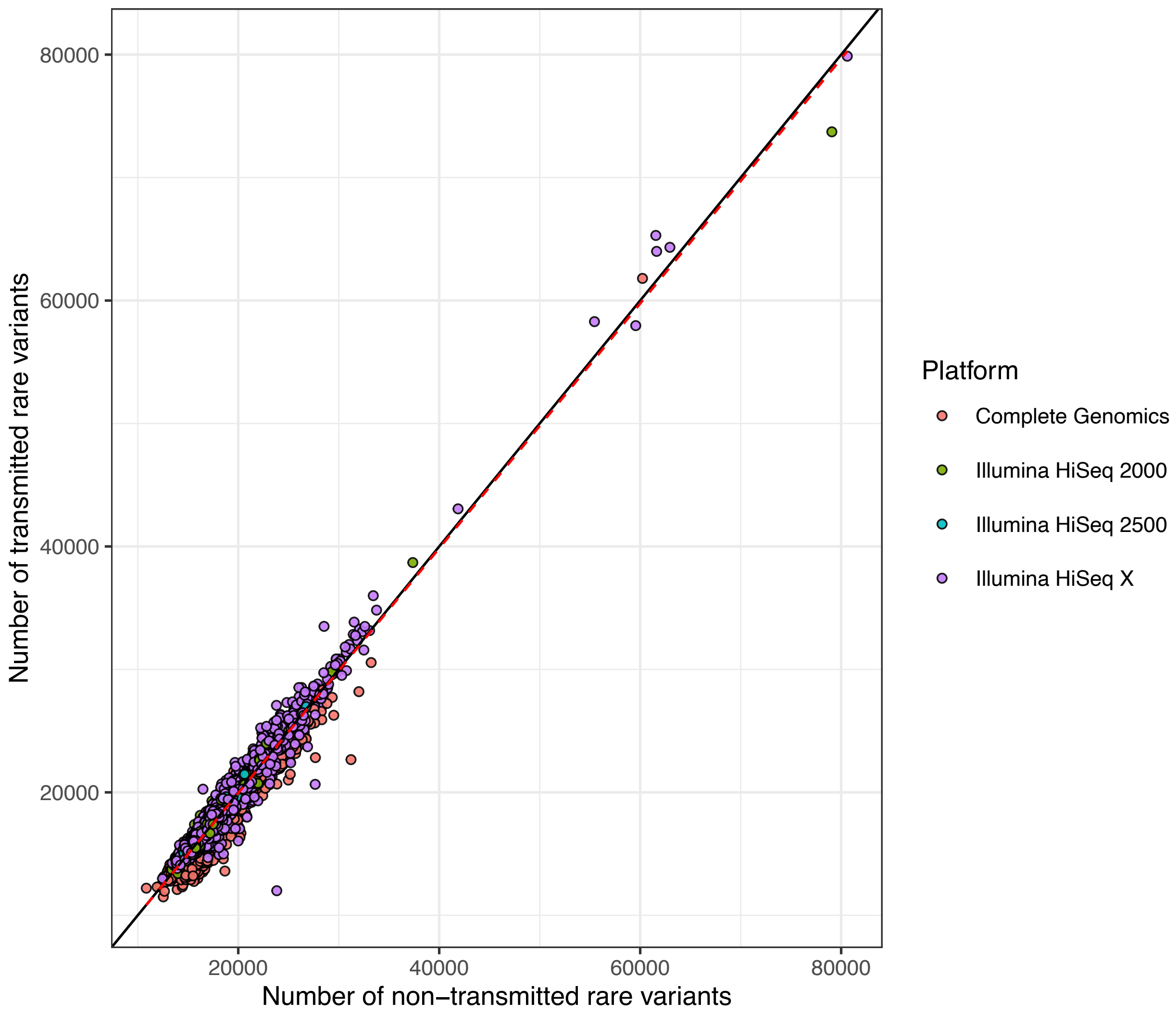

Number of transmitted rare variants
80000
60000
40000
20000
20000
40000
60000
80000
Number of non−transmitted rare variants
Platform
Complete Genomics
Illumina HiSeq 2000
Illumina HiSeq 2500
Illumina HiSeq X

**B**

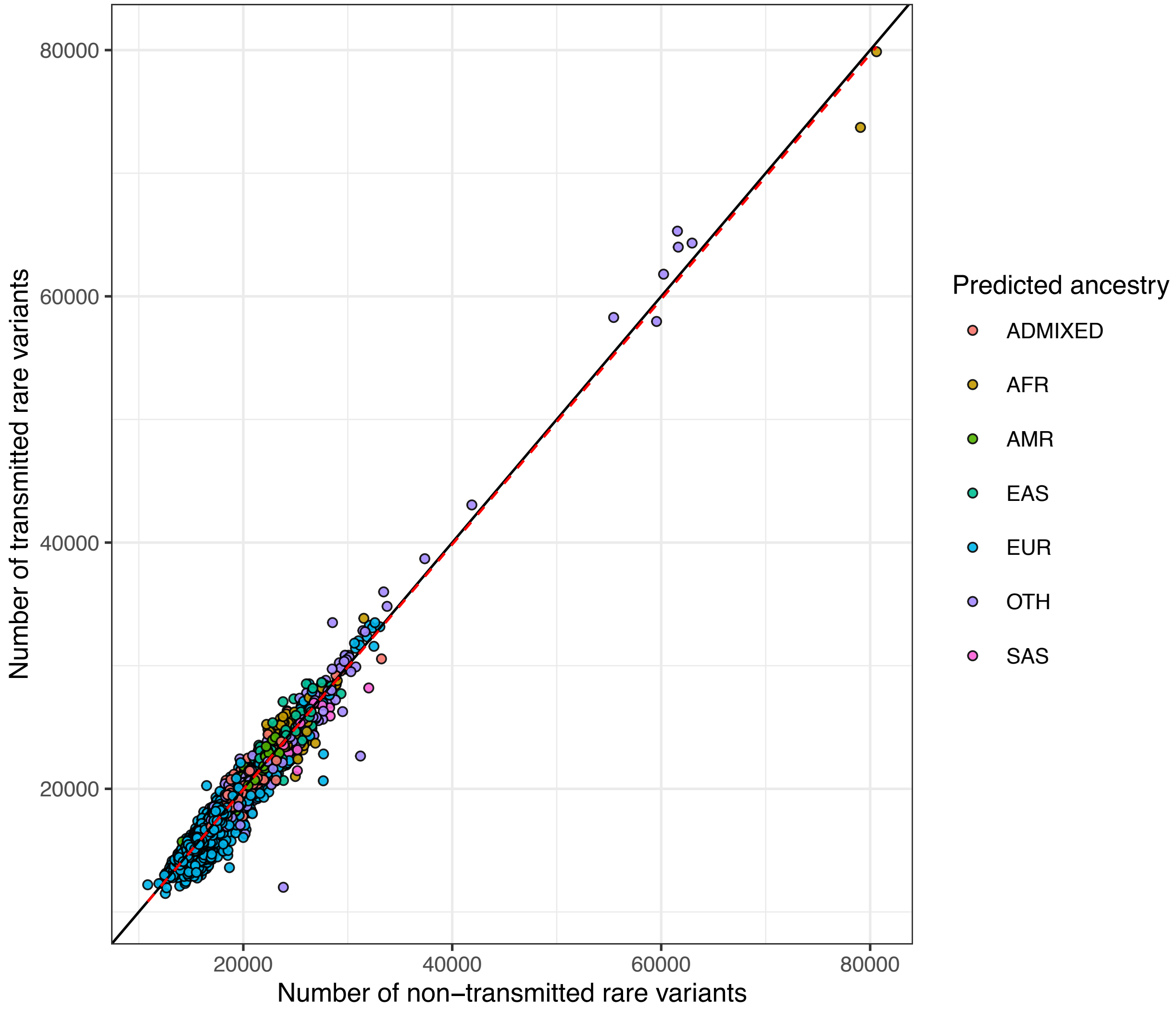

Number of transmitted rare variants
80000
60000
40000
20000
20000
40000
60000
80000
Number of non−transmitted rare variants
Predicted ancestry
ADMIXED
AFR
AMR
EAS
EUR
OTH
SAS

**C**

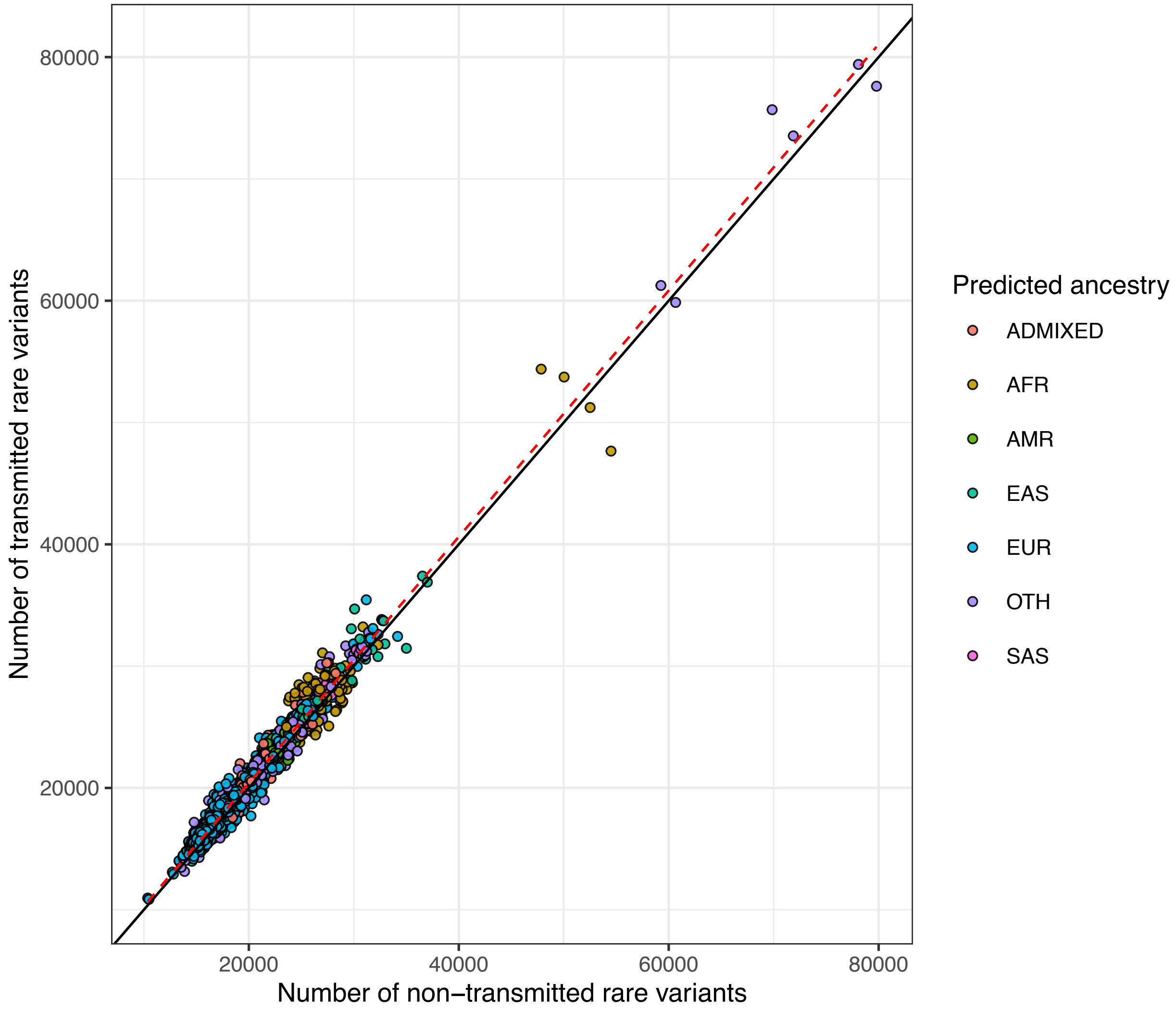


**Supplementary Figure 9:** Number of transmitted and non-transmitted rare variants in parents from MSSNG and SSC. (A) MSSNG parents, stratified by sequencing platform. (B) MSSNG parents, stratified by predicted ancestry. (C) SSC parents, stratified by predicted ancestry.

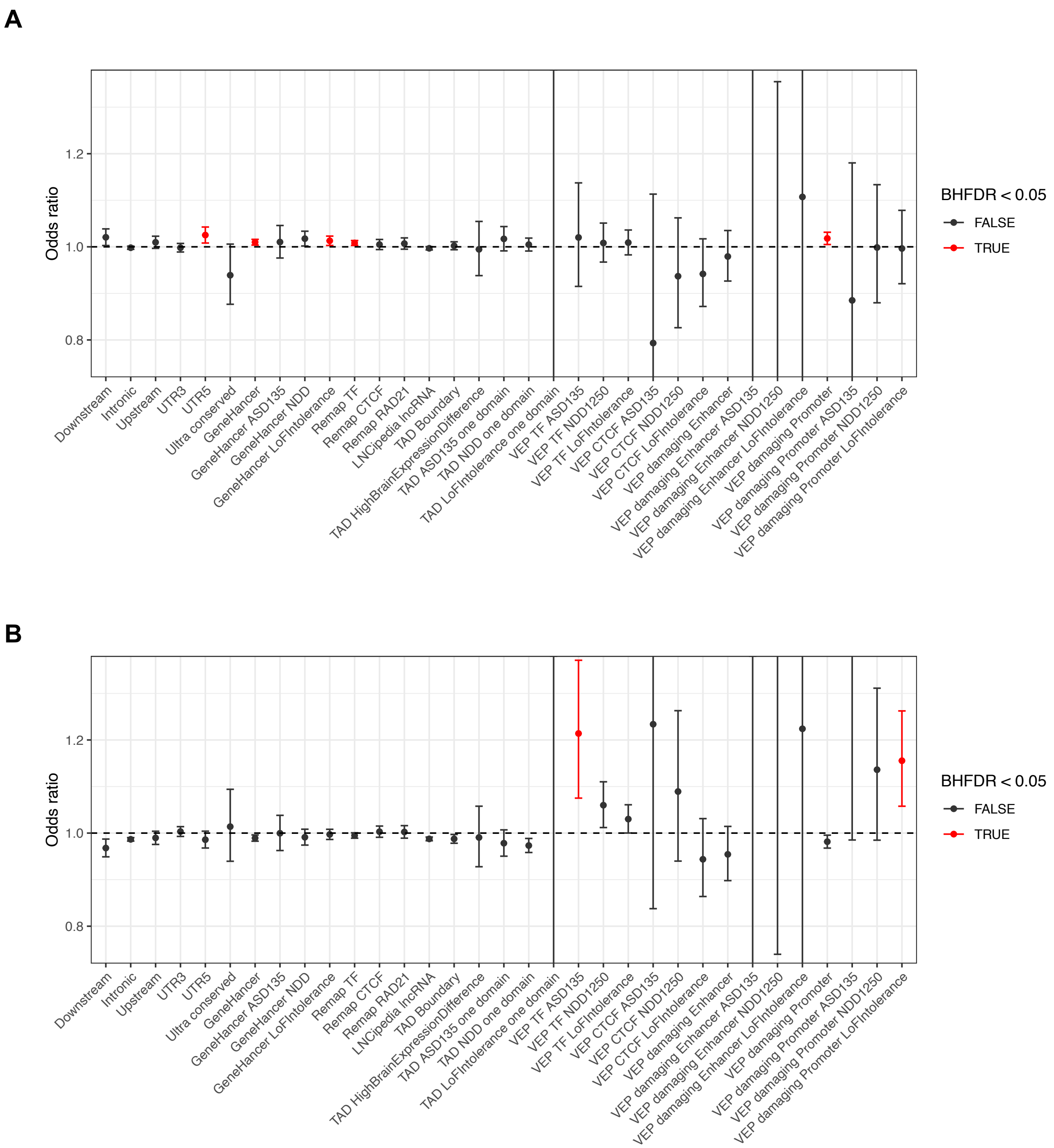
A
B
Odds ratio
1.2
1.0
0.8
BHFDR < 0.05
FALSE
TRUE
Downstream
Intronic
Upstream
UTR3
UTR5
Ultra conserved
GeneHancer
GeneHancer ASD135
GeneHancer NDD
GeneHancer LoFIntolerance
Remap TF
Remap CTCF
Remap RAD21
LNCipedia lncRNA
TAD Boundary
TAD HighBrainExpressionDifference
TAD ASD135 one domain
TAD NDD one domain
TAD LoFIntolerance one domain
VEP TF ASD135
VEP TF NDD1250
VEP TF LoFIntolerance
VEP CTCF ASD135
VEP CTCF NDD1250
VEP CTCF LoFIntolerance
VEP damaging Enhancer
VEP damaging Enhancer ASD135
VEP damaging Enhancer NDD1250
VEP damaging Enhancer LoFIntolerance
VEP damaging Promoter
VEP damaging Promoter ASD135
VEP damaging Promoter NDD1250
VEP damaging Promoter LoFIntolerance

**C**

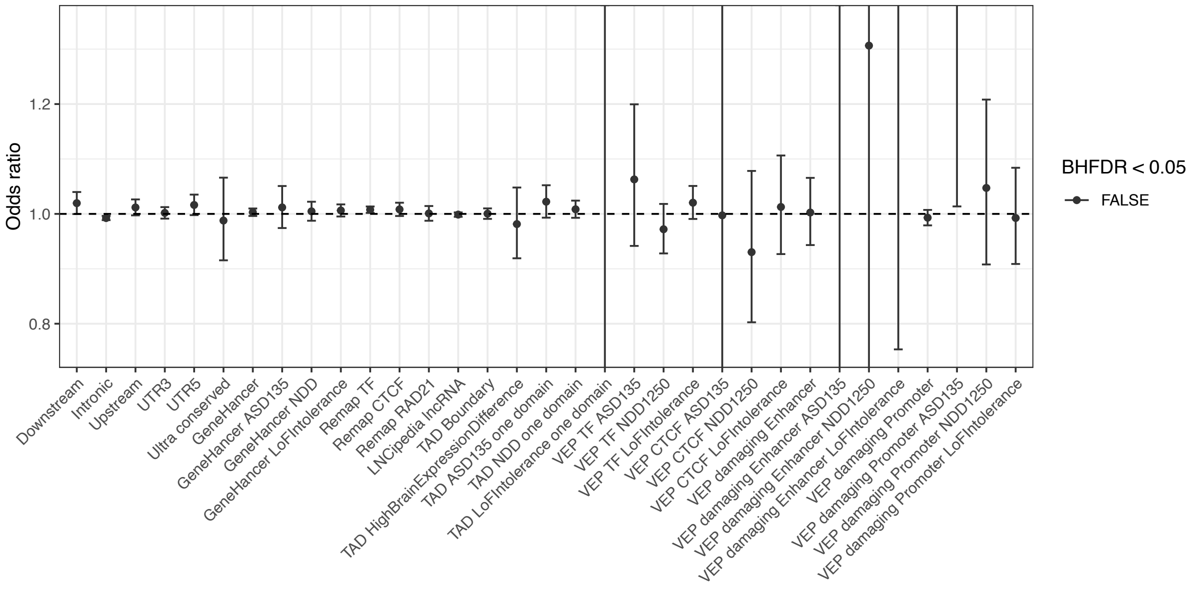


**Supplementary Figure 10:** Odds ratios for the number of rare variants (<0.1% frequency) in a given category that were transmitted versus non-transmitted in (A) Individuals with ASD in MSSNG, (B) individuals with ASD in SSC, and (C) unaffected siblings in SSC.

**A**

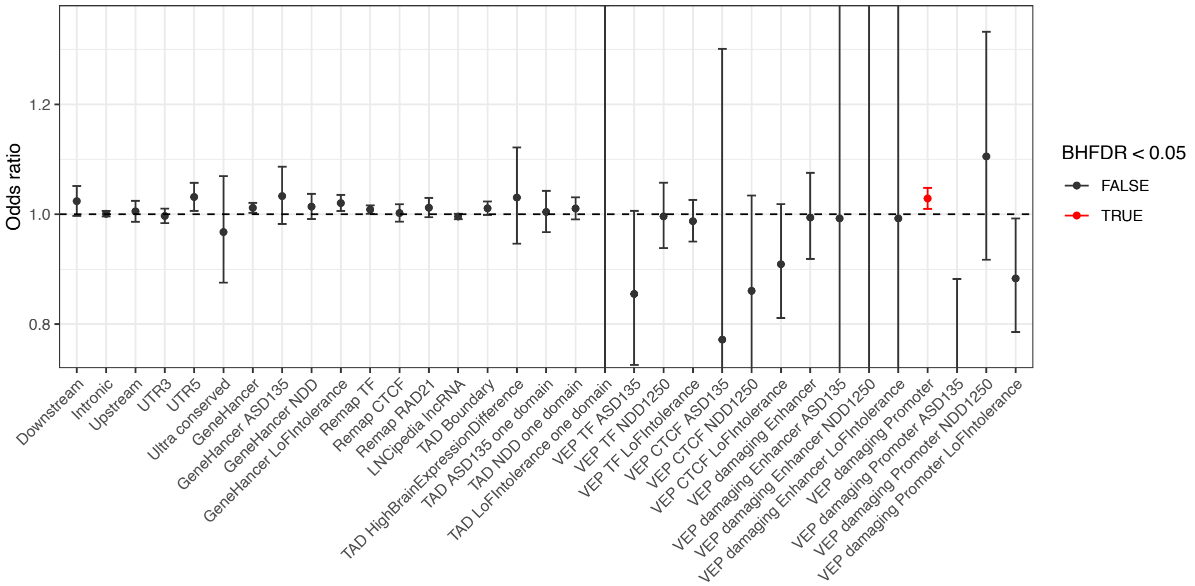
Odds ratio
1.2
1.0
0.8
Downstream
Intronic
Upstream
UTR3
UTR5
Ultra conserved
GeneHancer
GeneHancer ASD135
GeneHancer NDD
GeneHancer LoFIntolerance
Remap TF
Remap CTCF
Remap RAD21
LNCipedia lncRNA
TAD Boundary
TAD HighBrainExpressionDifference
TAD ASD135 one domain
TAD NDD one domain
TAD LoFIntolerance one domain
VEP TF ASD135
VEP TF NDD1250
VEP TF LoFIntolerance
VEP CTCF ASD135
VEP CTCF NDD1250
VEP CTCF LoFIntolerance
VEP damaging Enhancer
VEP damaging Enhancer ASD135
VEP damaging Enhancer NDD1250
VEP damaging Enhancer LoFIntolerance
VEP damaging Promoter
VEP damaging Promoter ASD135
VEP damaging Promoter NDD1250
VEP damaging Promoter LoFIntolerance
BHFDR < 0.05
FALSE
TRUE

**B**

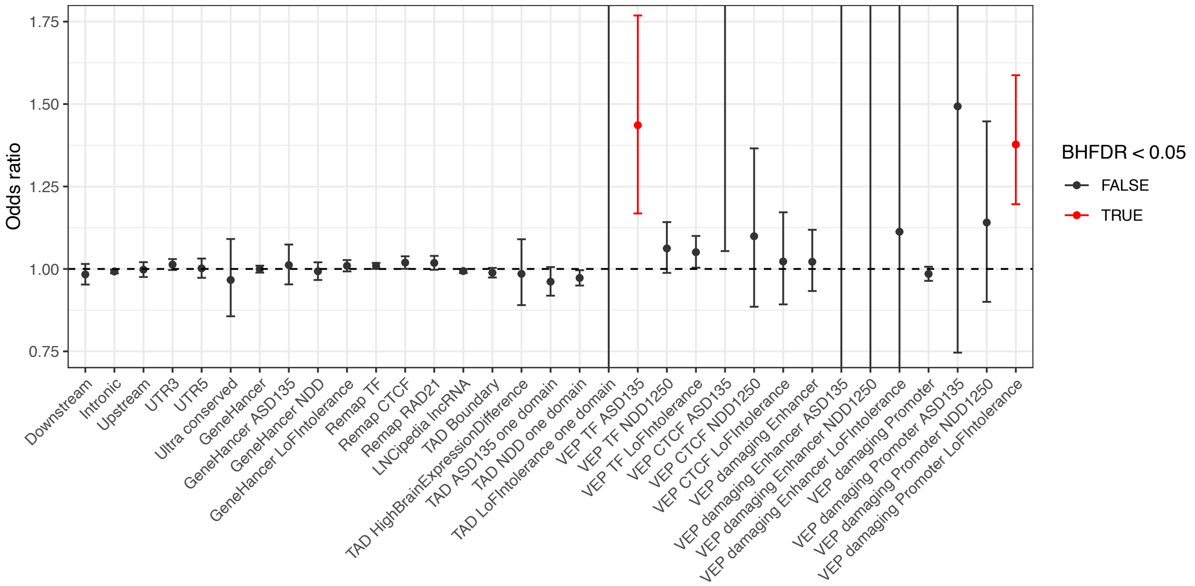
Odds ratio
1.75
1.50
1.25
1.00
0.75
Downstream
Intronic
Upstream
UTR3
UTR5
Ultra conserved
GeneHancer
GeneHancer ASD135
GeneHancer NDD
GeneHancer LoFIntolerance
Remap TF
Remap CTCF
Remap RAD21
LNCipedia lncRNA
TAD Boundary
TAD HighBrainExpressionDifference
TAD ASD135 one domain
TAD NDD one domain
TAD LoFIntolerance one domain
VEP TF ASD135
VEP TF NDD1250
VEP TF LoFIntolerance
VEP CTCF ASD135
VEP CTCF NDD1250
VEP CTCF LoFIntolerance
VEP damaging Enhancer
VEP damaging Enhancer ASD135
VEP damaging Enhancer NDD1250
VEP damaging Enhancer LoFIntolerance
VEP damaging Promoter
VEP damaging Promoter ASD135
VEP damaging Promoter NDD1250
VEP damaging Promoter LoFIntolerance
BHFDR < 0.05
FALSE
TRUE

**C**

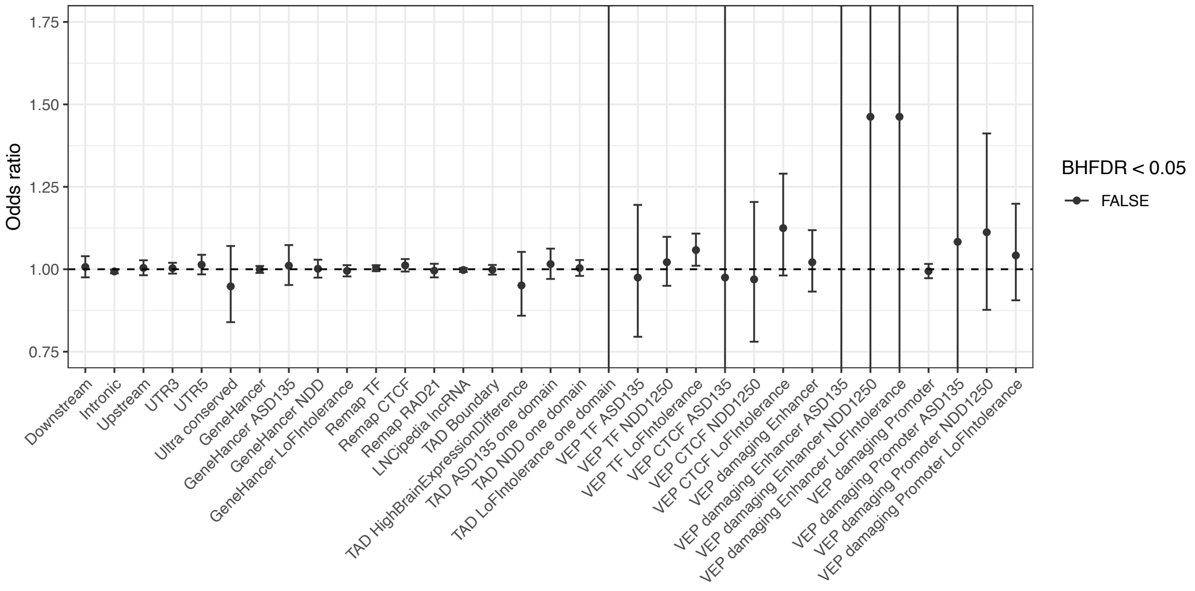


**Supplementary Figure 11:** Odds ratios for the number of singleton variants (private to a family) in a given category that were transmitted versus non-transmitted in (A) individuals with ASD in MSSNG, (B) individuals with ASD in SSC, and (C) unaffected siblings in SSC.

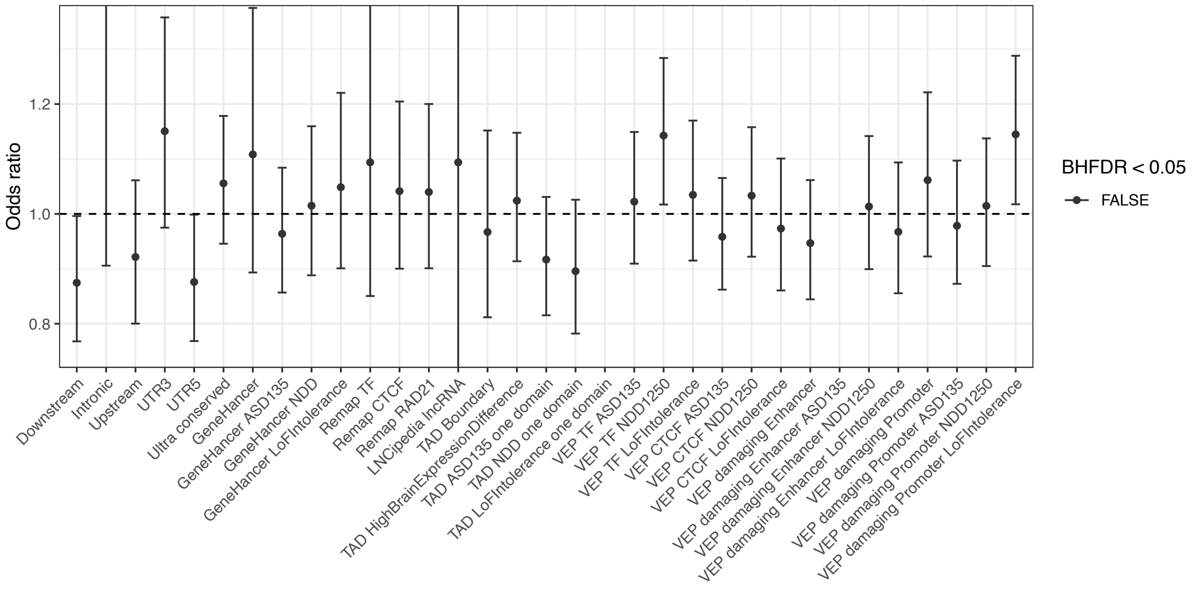


**Supplementary Figure 12:** Odds ratios for the number of rare variants in a given category in individuals with ASD from SSC versus their unaffected siblings.

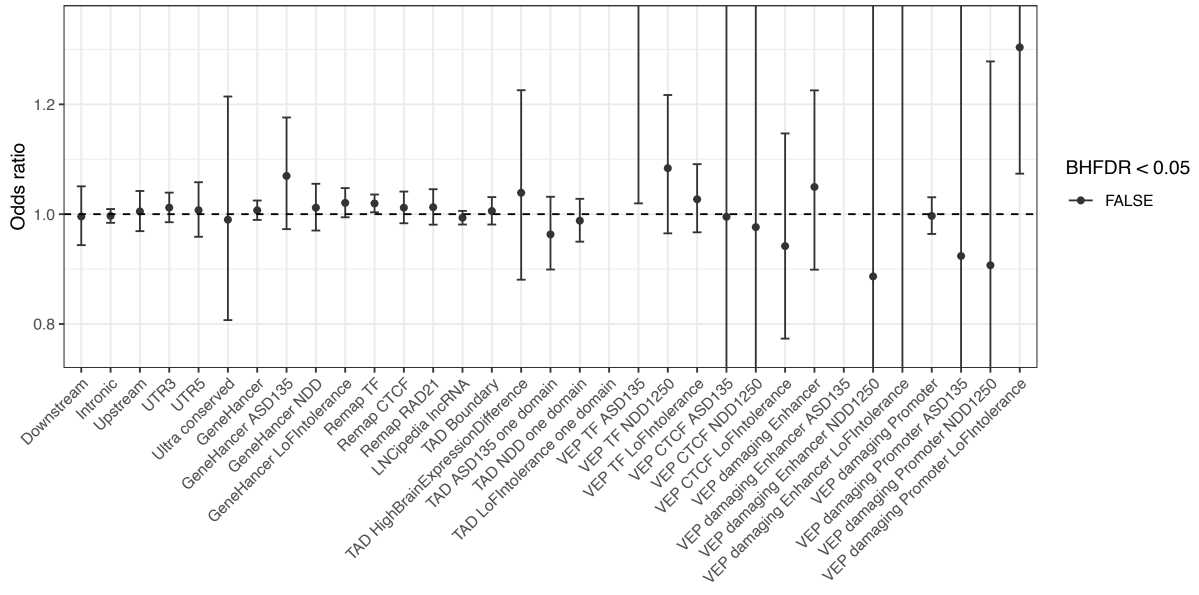


**Supplementary Figure 13:** Odds ratios for the number of singleton variants (private to a family) in a given category in individuals with ASD from SSC versus their unaffected siblings.

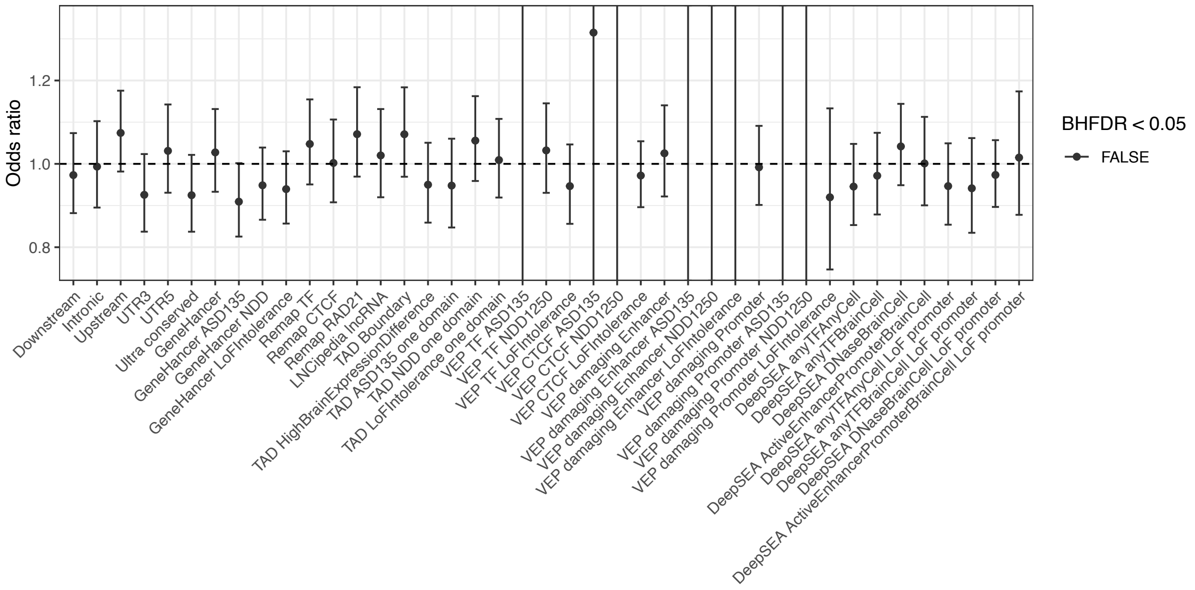


**Supplementary Figure 14:** Odds ratios for the number of *de novo* variants in a given category in individuals with ASD in SSC versus their unaffected siblings.

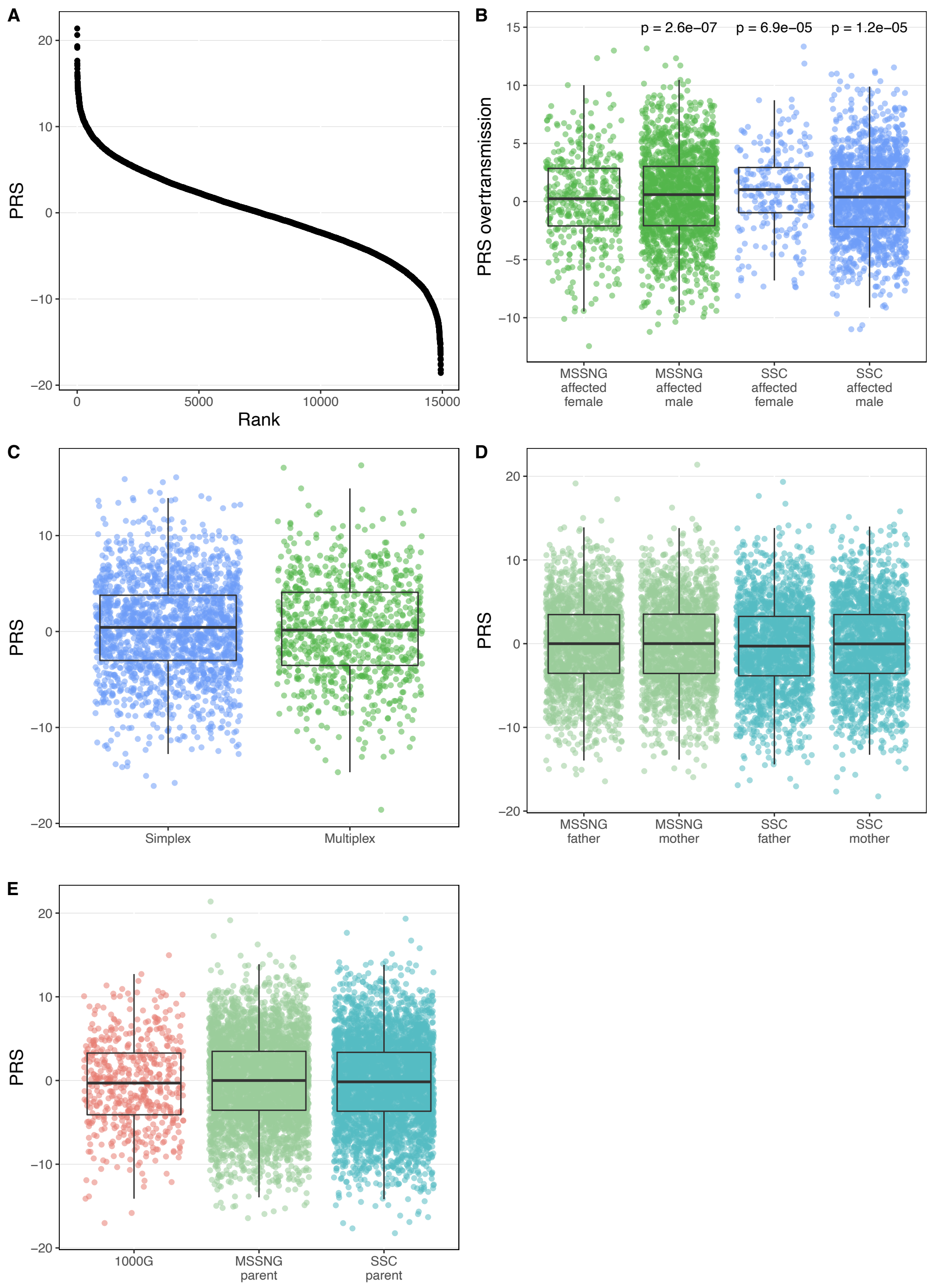


**Supplementary Figure 15:** Additional polygenic risk score (PRS) analysis. (A) PRS distribution when samples are ranked from highest to lowest score. (B) Sex-stratified polygenic transmission disequilibrium tests. (C) Comparison between individuals with ASD from multiplex families in MSSNG and individuals with ASD in SSC (which are all simplex families). (D) Comparison between unaffected fathers and unaffected mothers, stratified by dataset (MSSNG or SSC). (E) Comparison between unaffected parents and 1000 Genomes Project population controls.

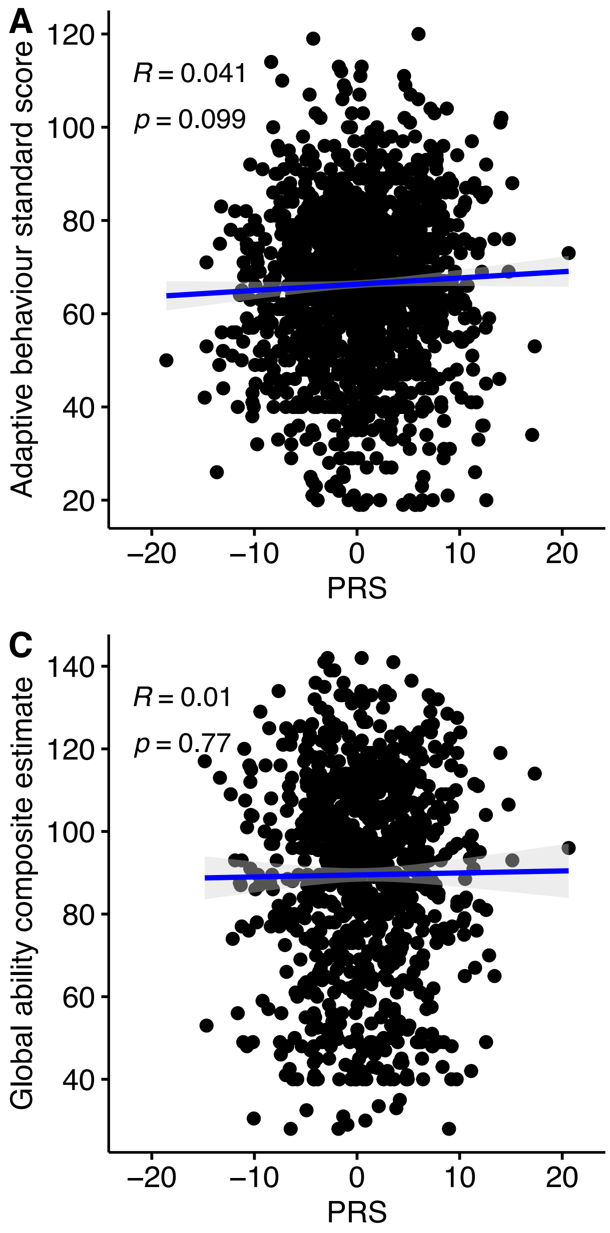


**Supplementary Figure 16:** Relationship between polygenic risk score (PRS) and composite phenotype measures. (A) Adaptive behavior standard score (n=2,787); (B) full-scale IQ (n=1,279); (C) global ability composite estimate (n=1,782); (D) socialization standard score (n=2,765).

**A**

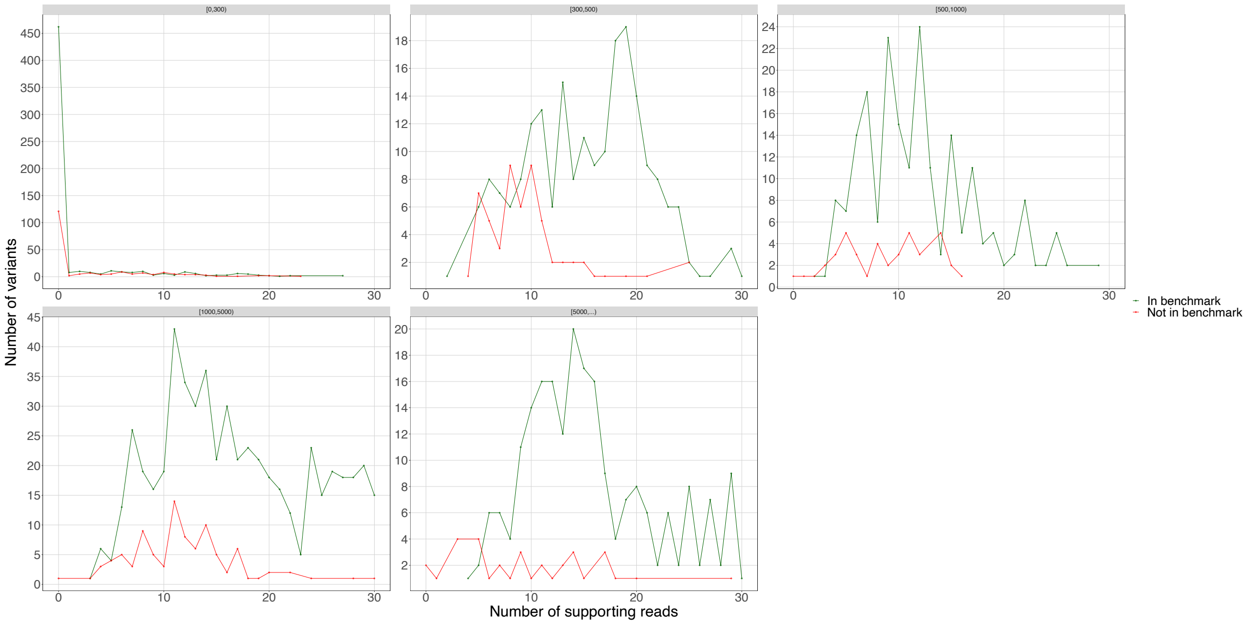


**B**

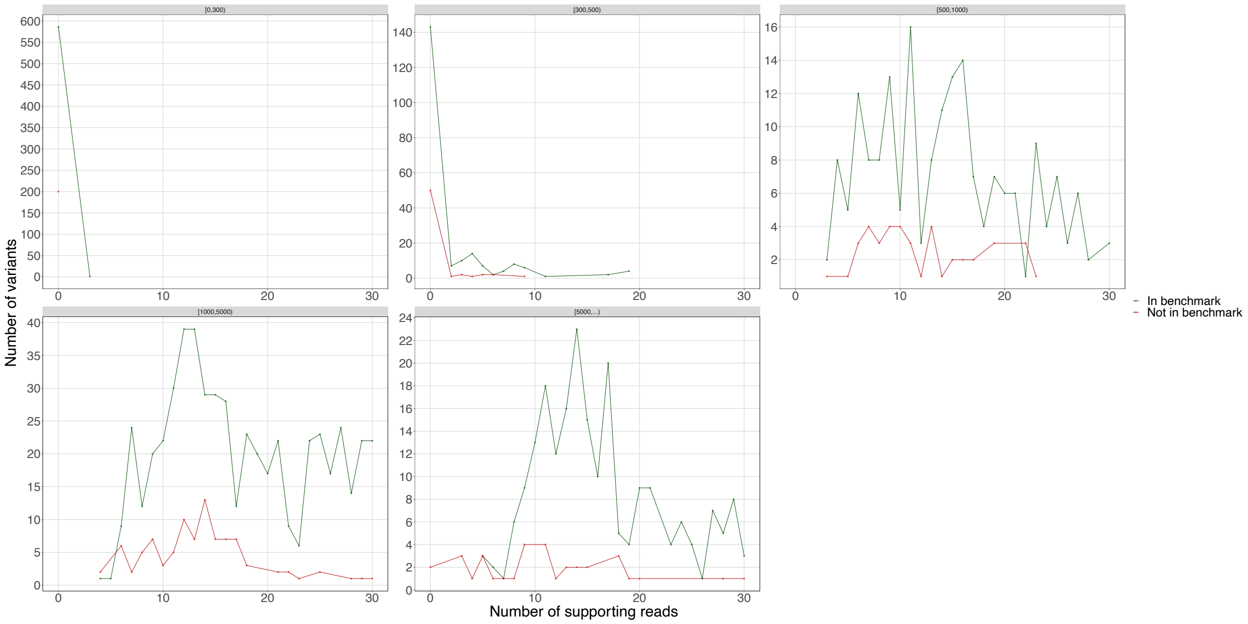


**Supplementary Figure 17:** Distributions of the number of deletions detected in the HuRef genome by (A) Manta or (B) DELLY that were or were not confirmed by the HuRef structural variant benchmark as a function of the number of supporting anomalously mapped paired-end reads. Similar trends were observed for the other two genomes/benchmarks tested (NA12878 and HG002) and for other variant types (duplications, insertions, and inversions). The facets indicate different deletion size bins; for example, the first facet represents deletions between 0 and 300 bp in size.

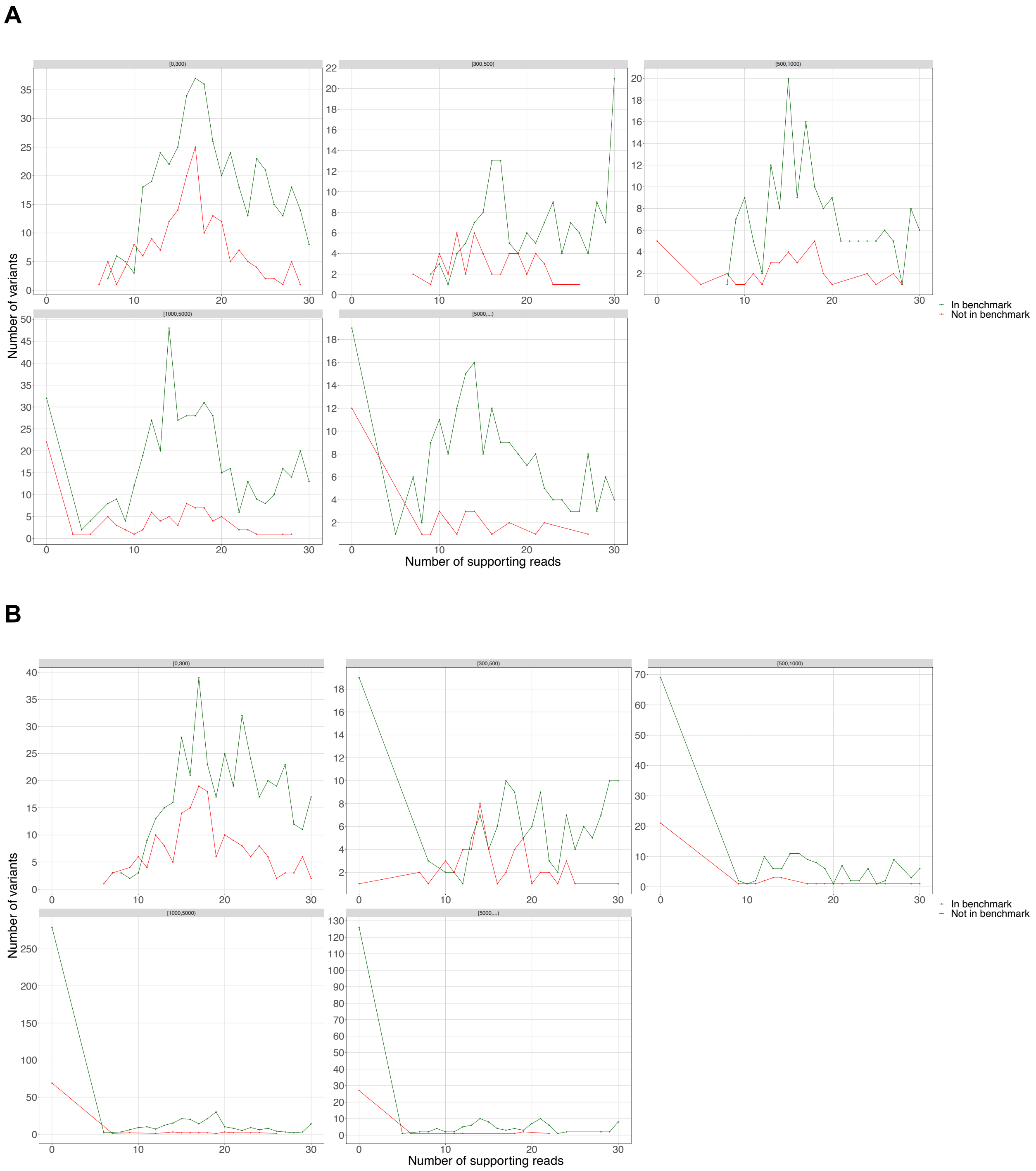


**Supplementary Figure 18:** Distributions of the number of deletions detected in the HuRef genome by (A) Manta or (B) DELLY that were or were not confirmed by the HuRef structural variant benchmark as a function of the number of supporting split reads. Similar trends were observed for the other two genomes/benchmarks tested (NA12878 and HG002) and for other variant types (duplications, insertions, and inversions). The facets indicate different deletion size bins; for example, the first facet represents deletions between 0 and 300 bp in size.

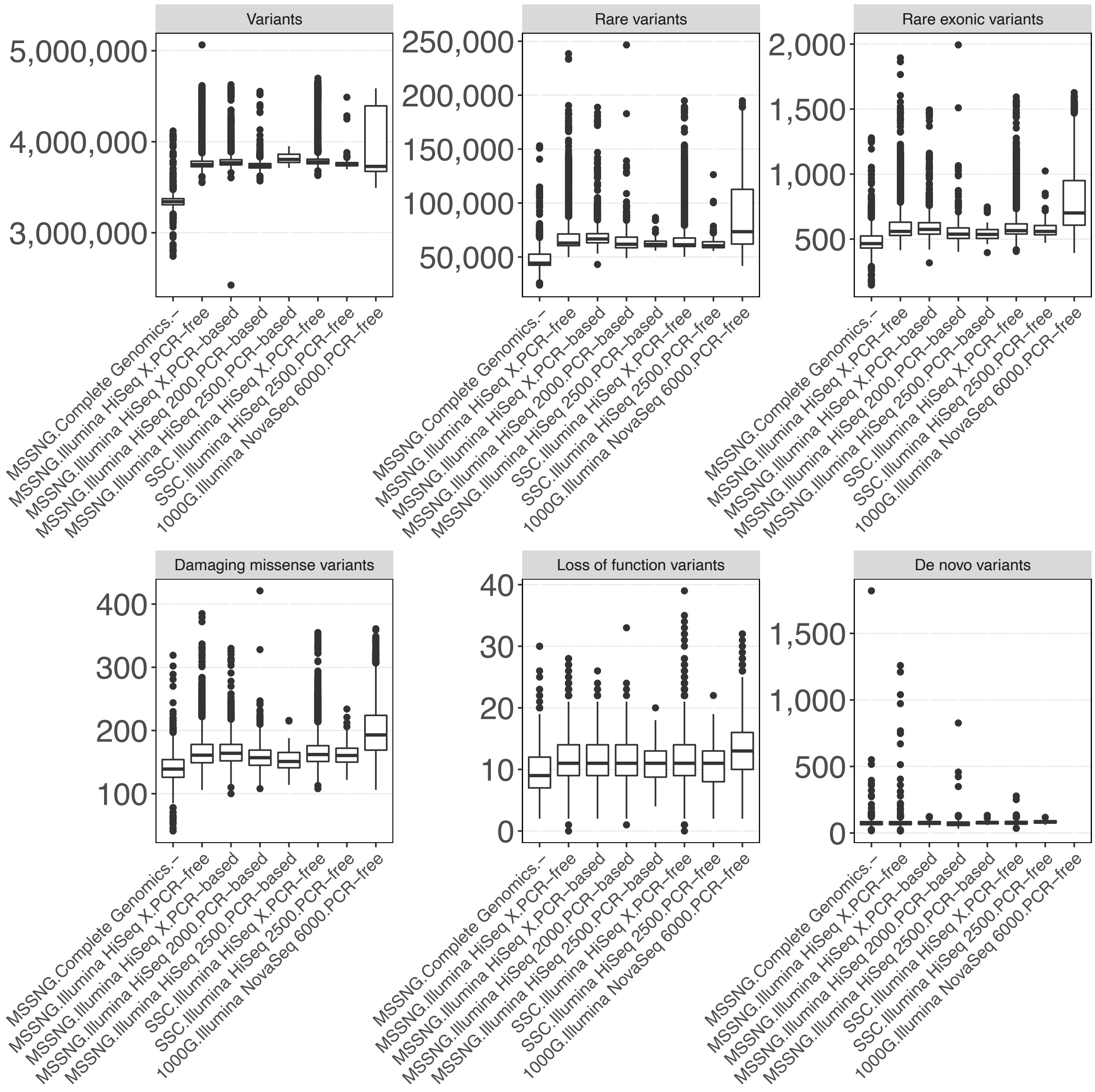


**Supplementary Figure 19:** Distributions of different categories of single nucleotide variants (SNVs), stratified by sequencing platform and DNA library preparation method.

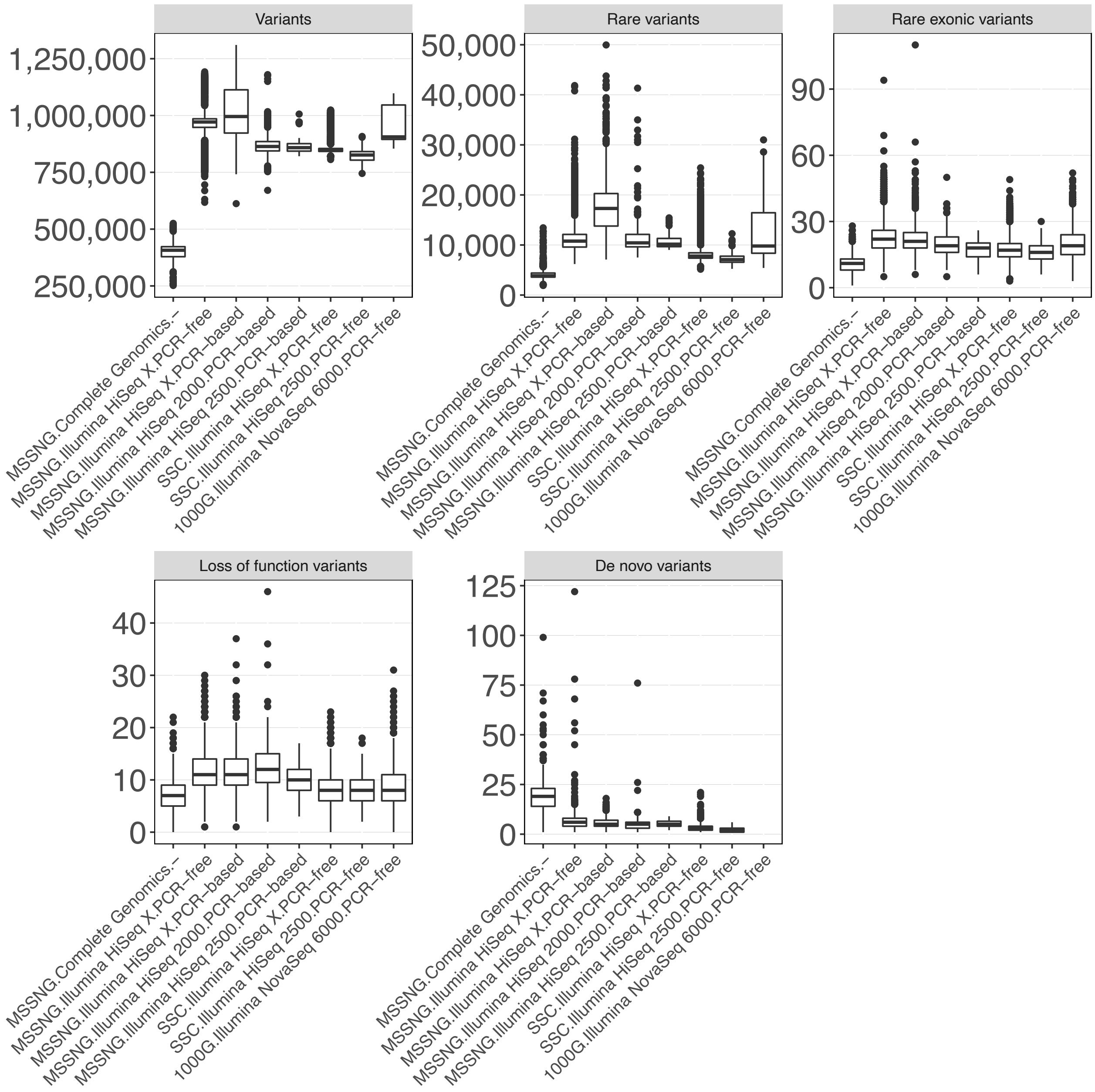


**Supplementary Figure 20:** Distributions of different categories of indels, stratified by sequencing platform and DNA library preparation method.

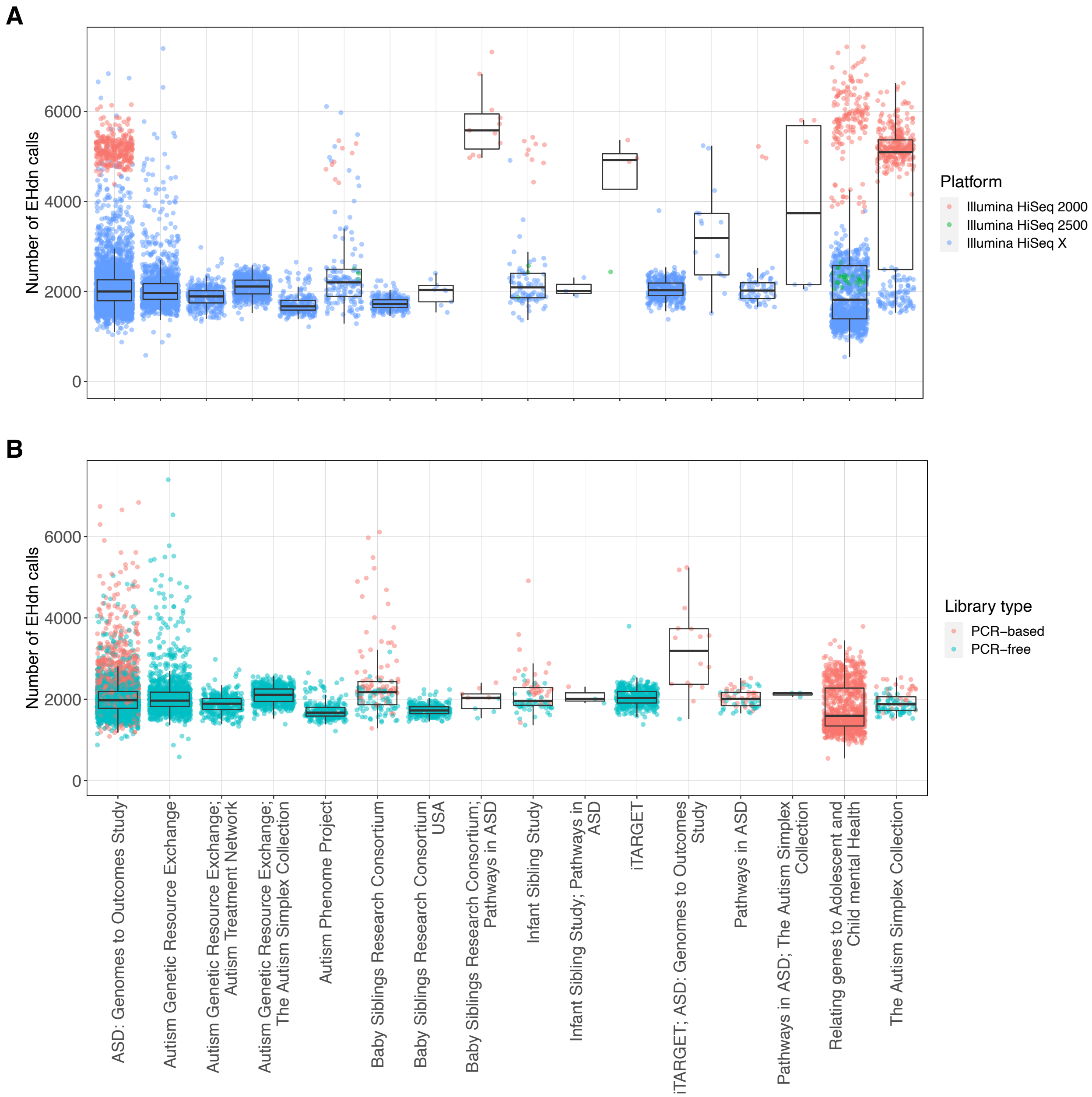


**Supplementary Figure 21:** Number of tandem repeats detected by ExpansionHunter Denovo in each MSSNG sample, stratified by cohort and colored by either (A) sequencing platform or (B) DNA library preparation method. In part (B), only samples sequences on the Illumina HiSeq X platform are included.

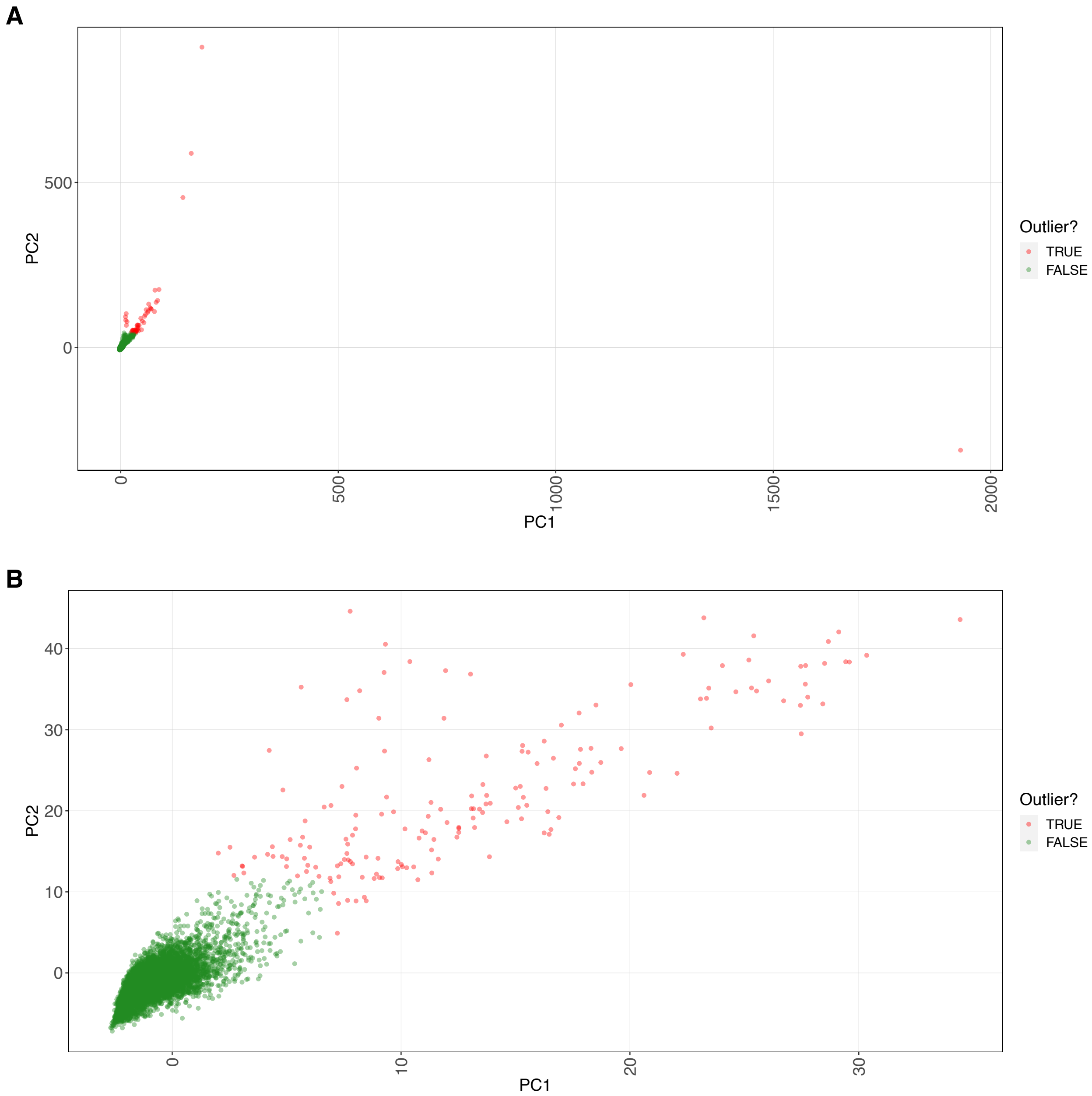


**Supplementary Figure 22:** Principal Component Analysis of tandem repeats detected by ExpansionHunter Denovo in MSSNG samples sequenced on the Illumina HiSeq 2500 or HiSeq X platforms. Outliers are indicated after (A) one round of outlier detection, and (B) two rounds of outlier detection (i.e., after removing the outliers in part A and recomputing the mean and standard deviation of each principal component).
